# Defining the causes of sporadic Parkinson’s disease in the global Parkinson’s genetics program (GP2)

**DOI:** 10.1101/2022.11.25.22282764

**Authors:** Clodagh Towns, Madeleine Richer, Simona Jasaityte, Eleanor J. Stafford, Julie Joubert, Tarek Antar, Alejandro Martinez-Carrasco, Mary B. Makarious, Bradford Casey, Dan Vitale, Kristin Levine, Hampton Leonard, Caroline B. Pantazis, Laurel A. Screven, Dena G. Hernandez, Claire E. Wegel, Justin Solle, Mike A. Nalls, Cornelis Blauwendraat, Andrew B. Singleton, Manuela M. X. Tan, Hirotaka Iwaki, Huw R. Morris, the Global Parkinson’s Genetics Program (GP2)

**Affiliations:** Department of Clinical and Movement Neurosciences, Queen Square Institute of Neurology, University College London, London, UK; Molecular Genetics Section, Laboratory of Neurogenetics, National Institute on Aging, National Institutes of Health, Bethesda, MD, USA; Department of Clinical Research, Michael J. Fox Foundation for Parkinson’s Research, New York City, NY, USA; Center for Alzheimer’s and Related Dementias (CARD), National Institute on Aging and National Institute of Neurological Disorders and Stroke, National Institutes of Health, Bethesda, MD, USA; Data Tecnica International, Washington, DC, USA; Department of Medical and Molecular Genetics, Indiana University School of Medicine, Indianapolis, IN, USA; Integrative Genomics Unit, Laboratory of Neurogenetics, National Institute on Aging, National Institutes of Health, Bethesda, MD, USA; Department of Neurology, Oslo University Hospital, Oslo, Norway

**Author notes:** Corresponding author: Professor Huw R Morris, Department of Clinical and Movement Neurosciences, UCL Queen Square Institute of Neurology, Royal Free Hospital, Rowland Hill Street, London NW3 2PF,.

## Abstract

The Global Parkinson’s Genetics Program (GP2) will genotype over 150,000 participants from around the world, and integrate genetic and clinical data for use in large-scale analyses to dramatically expand our understanding of the genetic architecture of PD. This report details the workflow for cohort integration into the complex arm of GP2, and together with our outline of the monogenic hub in a companion paper, provides a generalizable blueprint for establishing large scale collaborative research consortia.

## Main Text

Parkinson’s disease (PD) is a multifactorial disorder with complex etiology. The largest genome-wide association study (GWAS) to-date included 37,688 cases, 18,618 proxy cases (unaffected first-degree relatives), and 1.4 million controls from European ancestry, and identified 90 independent risk signals across 78 genomic regions; 38 of which were novel signals^1^. Despite these advances, PD GWAS are currently limited by scale, a focus on European populations, and limited integration with clinical phenotype data.

A power calculation based on the 2019 GWAS data indicates that inclusion of an additional ∼99,000 cases would enable variants of smaller effect size that contribute to polygenic risk (*p*-value cut off: 1.35 × 10^−3^) to reach genome-wide significance. Therefore, expanding PD GWAS to at least this size will result in identification of additional risk loci and improve genetic prediction of PD occurrence^1^. The heritability of PD can be estimated using twin studies or statistical genetic methods and is thought to lie between 22% and 40% in European populations. Known genome wide significant loci currently explain ∼ 16% of the heritability of PD^1^. The use of polygenic risk score analysis (including loci that do not reach genome wide significance) indicates that there are likely to be a substantial number of loci contributing to PD risk that have not yet been defined. Our power analysis indicates that 99,000 PD cases will be needed to define loci with 80% power, a minor allele frequency of 0.21 and similar effects to the current state of the art analysis. The variability of phenotypes observed in PD are likely to have a genetic basis.^2,3,4^ Knowledge of associations between genotype and clinical outcomes will enable clinicians to provide patients with a more accurate prognosis. Understanding the gene-to-phenotype pathways responsible for specific PD features would provide an opportunity to develop treatments targeting phenotype-specific disease pathways, resulting in more efficient and personalized treatment with fewer side effects. We aim to capture the diversity of PD outcomes, including Parkinson’s itself but also related conditions such as dementia with Lewy bodies and atypical parkinsonisms, and to perform large-scale analyses of clinical-genetic data with sufficient power for gene discovery. This will comprise regression and time to event analysis for the phenotype of interest (e.g. dementia, dyskinesias, motor progression). It is likely that this will be limited to around 25% of samples included in this study with in depth longitudinal data, and further large scale longitudinal cohorts will be needed to explore the biology of progression and diverse phenotypes. The focus of PD GWAS on individuals of European ancestry has left gaps in our knowledge of PD-associated genetic variants in underrepresented populations and limited our ability to resolve GWAS loci ^5^. To advance understanding of the genetic determinants of PD on a global scale, we need to ensure representation of diverse ancestries with sample sizes sufficient to detect ancestry-specific signals. We aim to include at least 15,000 participants of African, South Asian and East Asian ancestry respectively.

The Global Parkinson’s Genetics Program (GP2, http://gp2.org/), funded by the Aligning Science Across Parkinson’s initiative (ASAP, https://parkinsonsroadmap.org/) will recruit PD cohorts from across the world ^6^. We will genotype >150,000 participants and integrate genetic data with harmonized clinical data for use in case-control and genotype-phenotype association studies. This figure will include a minimum of 50,000 individuals from ancestries currently underrepresented in PD research: including Black American, African, Middle Eastern, Central/ East/South Asian, Indian, Caribbean and Central/South American. The remaining participants are expected to be of European ancestry.^6^ Cleaned genetic and clinical data will be harmonized across cohorts and made available to PD researchers via a controlled-access online repository with a unified user agreement. Contributing investigators are encouraged to play an active role in the project by suggesting and leading analyses, and will receive authorship on GP2 publications as well as support for their analyses. GP2 also provides comprehensive training opportunities for researchers from contributing institutions via an online learning management system. The outcome of GP2 will be an open access resource which will integrate clinical and genetic data from a very large, diverse sample of PD cases, and facilitate discovery of new genetic determinants of PD occurrence and phenotypic variation in multiple ancestries. A global network of PD researchers will be established, facilitating future collaboration and advancement of PD research. The project began in January 2020 and current funding extends to 2029.

Coordinating cohorts around the world to include 150,000 participants is a considerable logistical challenge. As of May 2022, we have established a workflow for cohort integration, a standardized set of clinical data elements, a recommended consent template, a protocol for evaluating cohorts joining the study, and a process for integrating and harmonizing the incoming clinical data. More broadly we have established the complex hub framework, which created a code of conduct and publication policy, created a DNA quality and shipping protocol, and created the NeuroBooster Array (NBA) genotyping workflow.

The size and depth of the dataset which is being generated by GP2 will provide a major opportunity to discover new genotype-phenotype associations. Core analyses, analyses that will be continuously updated by the analysis teams at GP2 with the inclusion of new samples, will include case-control, age at onset, and progression GWAS, as well as within-ancestry analyses of previously underrepresented populations. Beyond this, GP2 is supporting additional analytical projects proposed by contributing collaborators. The GP2 network will also facilitate collaboration on auxiliary studies between investigators sharing the same area of interest (e.g., biomarkers of different modalities) to address outstanding questions in PD research.

GP2 is conducted according to overarching principles of democratization of data, collaboration and cooperation, safe and responsible data sharing, commitment to diversity in research, transparency and reproducibility, and production of an actionable resource in accordance with the Findable, Accessible, Interoperable and Reusable (FAIR) scientific data management principles.^7^ Here, we report the specific steps undertaken by the Cohort Integration Working Group (CIWG) to identify, recruit, and harmonize clinical cohorts within the complex disease arm of GP2. GP2 also has a monogenic arm (https://gp2.org/working-groups/) which focuses on cases with potential monogenic causes of PD, i.e. those with early age at onset or a family history of PD (*monogenic protocol reference*).

## Methods

PD investigators and cohorts are identified through relevant publications, or through existing PD consortia. We welcome contact from interested investigators around the world. Prospective collaborators complete a <u>Site Interest Form (SIF)</u> (Supplementary Material) to provide a brief overview of their cohort. Data availability, type of cohort (e.g., brain bank, drug study etc.), ethnic diversity and sample/data sharing restrictions are all considered by the CIWG when reviewing SIF submissions. Cohorts’ consent documents are reviewed by the Operations and Compliance Working Group (OCWG) to ensure consent for sample/data sharing was obtained, and compliance with regional and cohort-specific data sharing restrictions. The OCWG assists the investigator if revisions are needed, and template consent language is provided online (https://gp2.org/resources/consent-guidelines/ [Supplementary Material]). We have an inclusive approach; to-date >95% of cohorts evaluated have been accepted. Following cohort approval, a concise Collaboration Agreement is signed by the collaborating PI and the Michael J. Fox Foundation (MJFF), and the necessary material and data transfer agreements are completed. The cohort’s samples are then transferred to one of GP2’s genotyping centers, and data are transferred through secure upload to a cloud server.

Once the cohort’s clinical data have been transferred, harmonization and quality control (QC) are performed by the CIWG, following a standard framework.^8^ Core clinical data were decided on the basis of clinical scales available in existing PD cohorts (primarily the Tracking Parkinson’s ^9^, PPMI^10^, PDBP^11^, NET PD-LS1^12^ and GEoPD^13^ cohorts) as well as on the basis of recent proposals for a modular set of assessment criteria to characterize longitudinal PD cohorts.^14,15^ The core clinical data elements are the clinical data categories of interest which will be harmonized across cohorts and used for analysis (Table 1). We have generated a common dataset for brain banks, and have added data elements for time to event analysis (Supplementary Table 1). We are also collecting information on availability of other data and samples, including, plasma, serum, RNA, fibroblasts, CSF, skin biopsy, PBL, iPS to expand awareness of the resources that are available at contributing sites (Supplementary Material).

**Table 1.**
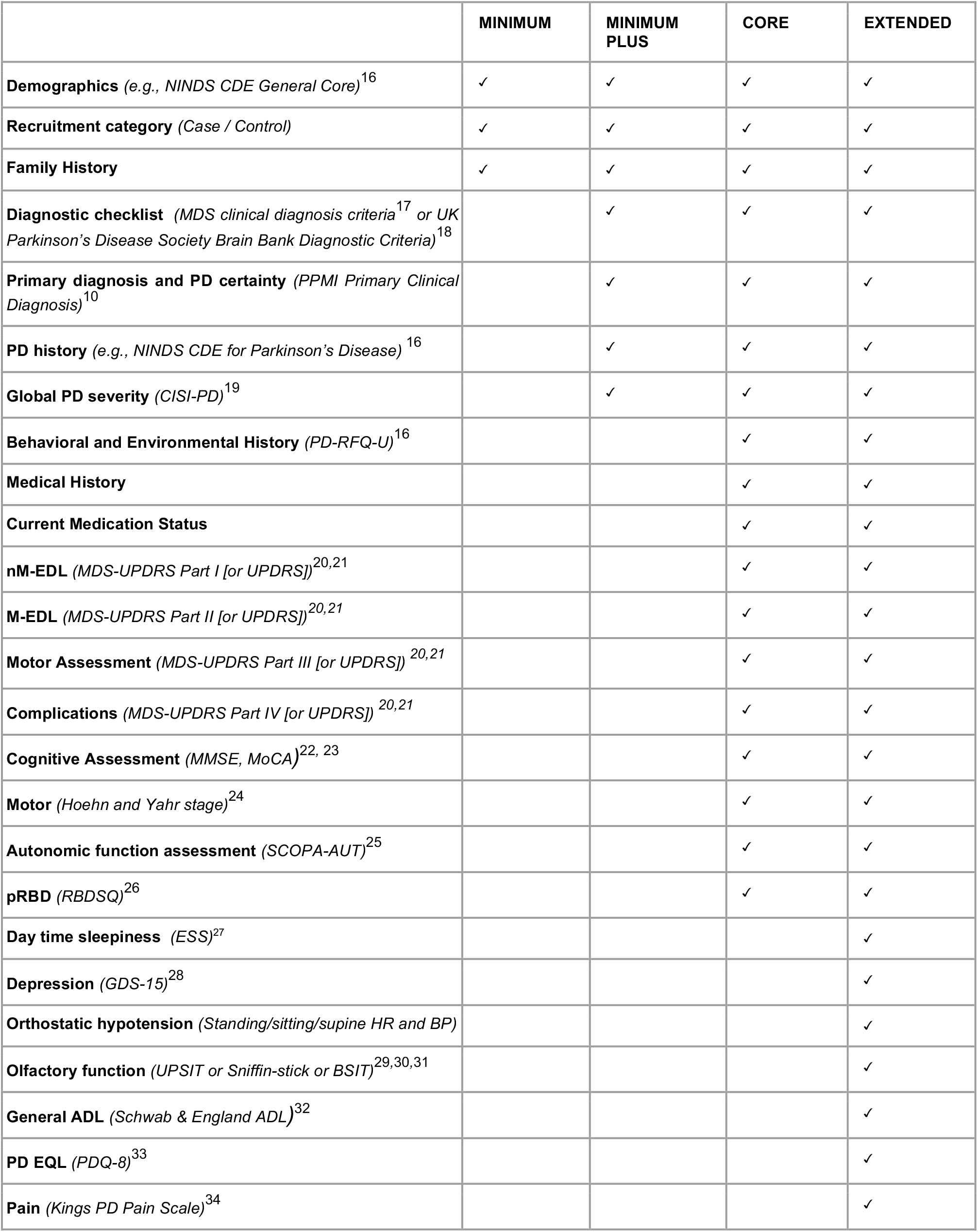
Recommended Core Clinical Data.

The common dataset is defined in a common data dictionary (Supplementary Table 2). The harmonization of raw data uses a set of coding rules for re-coding and handling of missing data, using a modified custom script for each cohort.

DNA samples from each cohort are genotyped on the Neuro Booster Array (NBA; https://github.com/GP2code/Neuro_Booster_Array), developed in collaboration by Data Tecnica International, NIA, NINDS, Illumina Inc and GP2. The NBA consists of the Illumina Infinium Global Diversity Array (GDA; https://www.illumina.com/products/by-type/microarray-kits/infinium-global-diversity.html), a high-density (1.9M total variants) global backbone optimized for cross-population imputation coverage of the human genome, and additional custom content (>95,000 variants) which includes known causal variants for various neurodegenerative diseases and imputation boosters for underrepresented populations. QC of genotype data by the Data Analysis Working Group (DAWG) follows a standard pipeline. A custom genotype clustering file is used to ensure representation of the diverse genetic ancestries within the GP2 data. The clustering file used for the latest data release is based on 2,793 samples across 6 ancestry groups, and includes 420 Gaucher disease cases to capture variants of interest in the *GBA* risk gene. This file is available for download via the <u>GP2 Github repository</u> (https://github.com/GP2code). Cleaned genetic data are returned to the contributing investigator to use as they wish, and are uploaded to the GP2 repository for use in combined analyses (Figure 1). GP2 covers the costs of genotyping and sample shipment for all contributing cohorts, and can assist investigators with the analysis of their cohort’s data.

**Figure 1.**
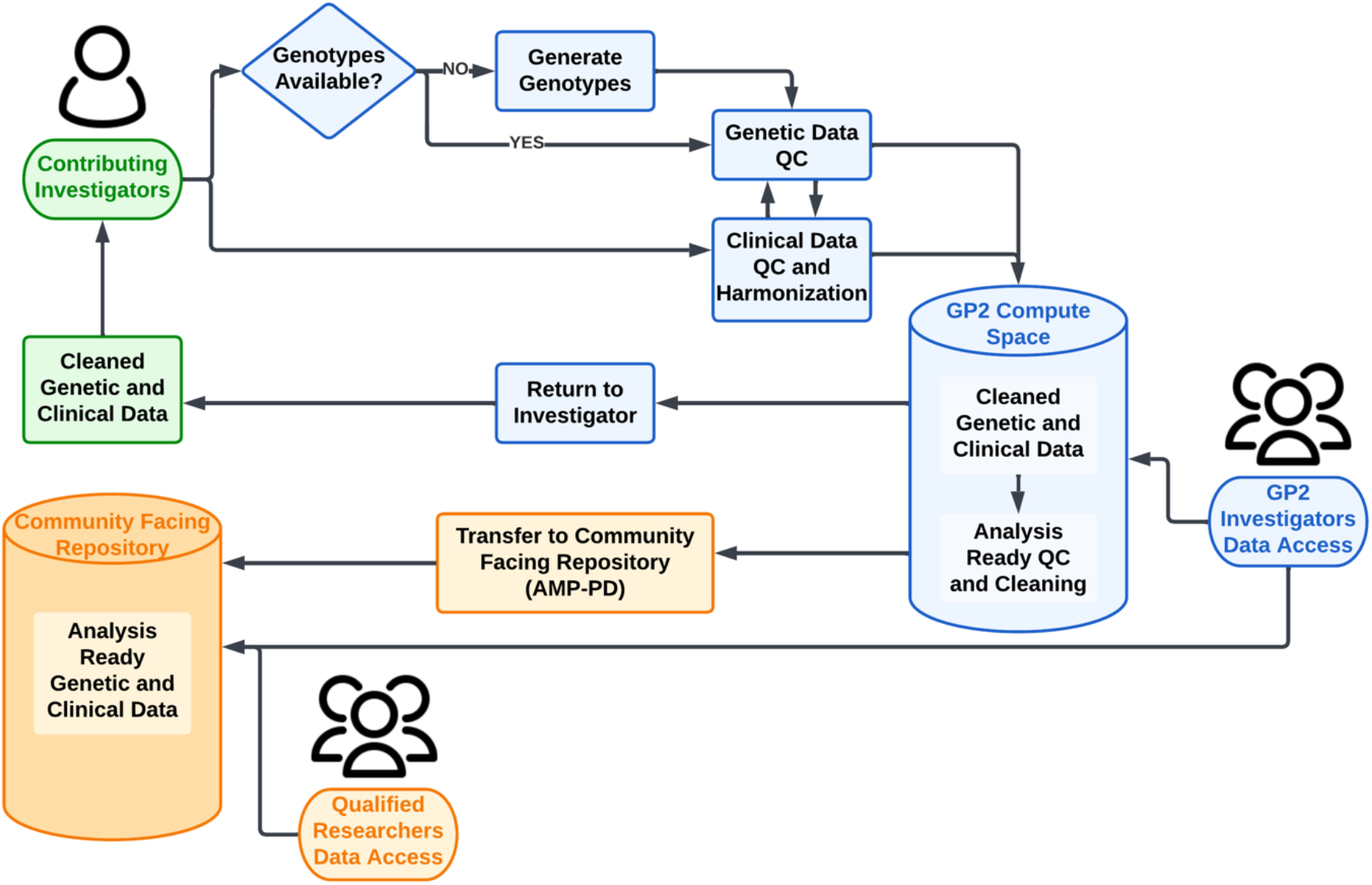
Cohort Integration Workflow. Data are contributed and returned to the local PI (green); new genetic data, data cleaning and harmonization is carried out by GP2 and made available for analysis within the GP2 consortium (blue); and data are released to qualified investigators via AMP-PD (orange). AMP-PD: Accelerating Medicines Partnership - Parkinson’s Disease (https://amp-pd.org)

The dataset which we are currently aggregating and harmonizing contains a wide variety of demographic and clinical factors, thanks to the diversity of contributing cohorts. As of March 2023, the CIWG has approved 145 cohorts for inclusion in GP2, of which 128 have been approved by the OCWG, and 74 have completed all necessary agreements and are in the process of transferring samples and clinical data. Samples that have been transferred are currently being genotyped and passed through the data QC pipeline. So far, the approved cohorts span over 50 different countries and territories. A map showing the geographic distribution of these cohorts, as well as current expected and completed sample numbers, can be found on the GP2 website (https://gp2.org/cohort-dashboard/ [Supplementary Material]).

## Supporting information

Global Parkinson's Genetics Program (GP2) - Banner Author List

## Data Availability

GP2 has partnered with the Accelerating Medicines Partnership - Parkinson's Disease (AMP-PD; https://amp-pd.org) to share data generated by GP2. Please see the "Data Availability" section within the manuscript for further details.

https://www.amp-pd.org/federated-cohorts/gp2#gp2-study-overview

## Data Availability

GP2 has partnered with the Accelerating Medicines Partnership - Parkinson’s Disease (AMP-PD; https://amp-pd.org) to share data generated by GP2, and in December 2021 the first GP2 genotyping data were released on the AMP-PD platform. As of 2023, the data consist of 14,902 samples (8,190 PD cases), representing a broad range of diverse ancestries defined directly from the genotyping data from these cohorts. Genotyping and data QC is ongoing, and there will be regular data releases (2-4 times per year) as the project progresses.

All contributing investigators have access to GP2 data, and external researchers can also gain access by following instructions on the GP2 website (https://gp2.org/applying-for-gp2-data-access-on-the-amp-pd-platform/). There are two tiers of data access. Tier 1 consists of summary statistics and any researcher can gain access by completing an online application. Tier 2 access includes deidentified, individual level genetic data, and to gain access researchers must sign a Data Usage Agreement co-signed by their institution.

## Code Availability

Code used in GP2 core analyses and the custom clustering files are deposited in the GP2 GitHub repository (https://github.com/GP2code). Code created and used by the CIWG can be found on the “GP2-WorkingGroups/CD-Cohort-Integration” subpage of the GP2 GitHub repository (https://github.com/GP2code/GP2-WorkingGroups/tree/main/CD-Cohort-Integration). The Data and Code Dissemination Working Group (DCD-WG) at GP2 is committed to making these data, resources, and training materials accessible to the scientific community. The DCD-WG actively supports efforts to consolidate analysis scripts with the necessary analytical tools and decisions via GitHub (https://github.com/GP2code), consolidate training materials with several translations in collaboration with the Training and Networking Working Group (https://gp2.org/training/), and consolidate additional analytical pipelines on Terra (https://amp-pd.org/tools).

## Inclusion and Ethics

GP2 is conducted according to overarching principles of democratization of data, collaboration and cooperation, safe and responsible data sharing, commitment to diversity in research, transparency and reproducibility, and production of an actionable resource in accordance with the Findable, Accessible, Interoperable and Reusable (FAIR) scientific data management principles. It is GP2 policy that local researchers are included in publications that use their data/samples.

## Acknowledgements

This research is supported by the Aligning Science Across Parkinson’s Initiative, the Intramural Research Program, National Institute on Aging, National Institutes of Health, Department of Health and Human Services, project ZO1 AG000949, and the Michael J. Fox Foundation for Parkinson’s Research.

Data used in the preparation of this article were obtained from Global Parkinson’s Genetics Program (GP2). GP2 is funded by the Aligning Science Across Parkinson’s (ASAP) initiative and implemented by The Michael J. Fox Foundation for Parkinson’s Research (https://gp2.org). For a complete list of GP2 members see https://gp2.org.

## Author Contributions

CT, MR, SJ, EJS, JJ, TA, ACM, MMXT, HI, and HRM are members of the GP2 Cohort Integration Working Group (CIWG), of which HRM is the lead, and MMXT and HI are co-leads. CT was the primary contributor in the drafting of this manuscript, assisted by MR, and JJ. The draft was reviewed by all CIWG members prior to circulation to other contributing authors for review. MBM is the co-lead of the GP2 Data and Code Dissemination Working Group and drafted the data availability section of this manuscript, together with BC who is senior associate director at the Michael J. Fox Foundation’s research division. DV, KL, HL and MAN are members of the GP2 Complex Disease Data Analysis Working Group (DAWG), of which MAN is the lead and HL is a co-lead. DAWG are responsible for the data cleaning and analysis described in this manuscript. DGH directs the Genomic Technologies Group within the Laboratory of Neurogenetics at NIH, which conducts the majority of sample genotyping for the Complex Network. CEW and JS are members of the GP2 Operations and Compliance Working Group of which JS is a co-lead. CBP and LAS are scientific program managers for the GP2 Complex Disease Network. ABS is the lead of the GP2 Complex Disease Network and CB is the co-lead. All listed authors reviewed the manuscript and provided comments and revisions prior to submission.

