## Supplementary Material for "Defining the causes of sporadic Parkinson’s disease in the global Parkinson’s genetics program (GP2)"

*Supplementary Material*

#### 1. GP2 Consent Guidelines and Recommended Language

The GP2 Consent Guidelines and Recommended Language can be viewed and downloaded at https://gp2.org/resources/consent-guidelines/. The current guidelines (Version October 2020) are below.

#### Consent Guidelines & Recommended Language

##### Summary

The Global Parkinson's Genetics Program (GP2) has developed the following template text to assist researchers who wish to submit samples and/or data to GP2. The template text is intended as a guide, and it can be adapted to reflect local requirements. However, the content should remain true to the goal of international data sharing. When developing your consent document, please be sure to include any additional content required by your local ethics review board which may not be listed below.

##### Introduction

If not consenting as part of another study: You are invited to take part in the Global Parkinson's Genetics Program (GP2). This is a research project. Researchers from around the world are collaborating through this project to study the genetics of Parkinson's Disease. GP2 includes people with and without Parkinson's Disease. You can find information about GP2 and the researchers leading the program at www.gp2.org.

At [name of institution], this project is being led by [name of investigator]. All research that uses peoples' personal data must be reviewed by an ethics board or committee. This project has been approved by [ethics board/committee information]. GP2 is funded by the Aligning Science Across Parkinson's (ASAP) initiative and The Michael J. Fox Foundation for Parkinson's Research.

If consenting as part of another study: You are invited to take part in [name of study]. This is a research project. [Explain the purpose of this study. Provide details regarding the institution(s), investigator(s), and ethics board review.]

[Name of study] is collaborating with researchers from around the world through the Global Parkinson's Genetics Program (GP2) to study the genetics of Parkinson's Disease. GP2 includes people with and without Parkinson's Disease. You can find more information about GP2 and the researchers leading the program at www.gp2.org.

##### Voluntariness

Your participation in this study is voluntary. The decision about whether or not to participate is completely up to you. If you choose not to participate in the study, your choice will not affect your medical care.

##### Research activities

If not consenting as part of another study: If you choose to participate, you will be asked to provide a small sample (less than a tablespoon) of blood [or saliva]. You will also be asked questions about your health and your family history of Parkinson's Disease.

If consenting as part of another study: [Explain the activities involved in this study. Include the collection of a blood or saliva sample for DNA.]

##### Payment for Participation

[GP2 will not provide payment for participation. If subjects will be reimbursed for time/inconvenience related to your research activities, please specify.]

[For US studies, specify cost/no-cost to participate.]

**Define the research data – genomic and personal data**

Your sample will be used to generate genomic data. Genes are the basic 'instruction book' for the cells that make up our bodies. Genes are made out of DNA, and all of the DNA in each cell is called the genome. Although our DNA is very similar to each other, your genomic data is entirely unique.

Other types of data about you will also be collected. This may include information about your sex, age, ethnicity, and diseases or syndromes you may have.

**Data coding (or pseudonymization)**

Information that directly identifies you, such as your name, will be replaced with a 'code' or 'ID number.' Your name and other identifying information will not be shared with other researchers.

**Describe future use**

Your samples and data will be used to learn about the genetic differences between people with and without disease. They may also be used in additional research studies involving PD, other neurological conditions, other types of disorders, or other biomedical research studies. These additional studies may involve development of cures, therapies and products and services for the benefit of PD and other patients.

**Explain international sharing**

This research may be conducted anywhere in the world; the researchers studying your samples and data may be located outside your country. These projects can also take place in universities, hospitals, nonprofit groups, for-profit companies, or government laboratories.

**Include commercial/non-academic use**

Your sample and data are a gift for research. The future research projects may take place in universities, hospitals, nonprofit groups, for-profit companies, or government laboratories.

Some of the research done with your information may one day lead to new software, tests, drugs, or other commercial products. If this happens, you will not receive any of the profits from these new products.

**Explain managed access (and unrestricted access for aggregate data)**

Your coded data, will be stored on a secured data platform. Information in this data platform can only be accessed and used by researchers who have been granted formal approval to access data and who have signed agreements to protect the confidentiality of the information. The access agreements also require researchers to respect the laws and ethical guidelines for scientific research.

Your coded data may also be combined with data from many thousands of other people in large-scale analyses. A summary of this data may be made public (openly accessible) to anyone without restriction.

**Include storage on cloud servers**

Your coded data will be stored on one or more data platforms coordinated by multiple institutions that can be used by researchers around the world. These researchers may be conducting their own projects or may be working on projects coordinated by the sponsors.

The data platform may be hosted on commercial cloud servers. The cloud refers to software and services that run on the Internet, instead of on a specific computer. These cloud servers meet international security and safety standards.

**Do not limit duration of storage**

Preferred: Your coded samples and/or data will be stored indefinitely or until they are withdrawn or no longer useful for current or future research.

If indefinite storage is not allowed: Your coded samples and/or data may be stored and used for research for 30 (thirty) years.

**Explain data withdrawal**

You may withdraw consent for research use of your samples and data at any time. If you choose to withdraw, your samples will be destroyed, and your data will be removed from the data platform. However, it may not be possible to retrieve data that has already been distributed for research use.

**Request consent to contact the participant for future research**

With your permission, we may re-contact you to invite you to provide additional data or to be involved in new research projects.

[Please verify requirements for consent to recontact with your ethics board].

**Discuss lack of benefit and risk of re-identification**

You will not benefit personally from sharing your data. Participating in the study may help researchers in many areas of scientific research, such as health and genetics.

Your information will be coded, which means it will not be connected to any information that directly identifies you, such as your name, address, and contact information. However, it is very difficult to make genetic information completely anonymous. There is a risk that people that have your information could try to connect it to your identity by combining it with other personal information about you, through a process called re-identification. Also, in the future, new technologies could be developed that make it easier to connect your genetic information to your identity. The risks related to re-identification are difficult to predict at this time.

Because genetic information is shared among people who are biologically related to you, it is possible that information about your family members could also be revealed.

There is always a risk that information from genetic studies might be used to make certain statements or conclusions about groups or communities. In some cases, this can lead to discrimination against individuals, families, groups or communities.

**Contacts**

[Provide contact information for both local study staff and the regulatory/ethics authority.]

#### 2. Clinical Cohort Meta-Data

GP2 is collecting information regarding other biosample and data types that are available for contributing cohorts. These samples/data will not be requested or shared by GP2. The information is intended to allow investigators to identify collaborators with similar interests and available data for auxiliary studies e.g., biomarker or imaging studies. The below table gives an indication of the availability of these data/sample types as of May 15th 2022.

| Data/sample modality | Number of cohorts with available data/samples |
|---|---|
| **MRI** | 43 |
| **DAT Scan** | 34 |
| **Plasma** | 57 |
| **Serum** | 55 |
| **RNA** | 28 |
| **PBL** | 21 |
| **LCL** | 3 |
| **CSF** | 24 |
| **Skin Biopsy** | 22 |
| **iPS** | 15 |
| **Post-mortem Brain Tissue** | 15 |

#### 3. Cohort Site Interest Form

Below is a blank copy of the Site Interest Form that cohort Investigators are asked to complete to register their interest in joining GP2. The submitted form is reviewed by the Cohort Integration Working Group to determine whether the cohort should be included. The numbers entered into the form can be approximate; it is intended to provide an overview of the cohort and samples/data availability. Investigators who are interested in joining GP2 can complete the live form here: https://forms.gle/tAEfZHu94jpu8g6i9

### GP2 complex trait group: Site Interest Form (Version 2)

Thank you for your interest in collaboration. We would like to ask some general questions about your cohort, number of available (will be available) data and samples, genotyping chips, and IRB approved information especially for data sharing policies. The form consists of 8 sections and will take 20 minutes to fill in. You can edit the input after you submit it. If you have any questions please contact.

*Required

1. Email *

2. NAME *

3. INSTITUTION *

4. SHORT NAME OF STUDY *

5. FULL NAME OF STUDY *

6. PI *

7. email *

https://docs.google.com/forms/d/1DDEO2z0D-FexwQCsi5vVuBnx34jPi_V3YGH59rF7Bq4/edit

8. Year Started *

______________________

9. Year Completed *

______________________

10. FOLLOW UP COMPLETED *

4-digit, 9999 if the end is not planned

______________________

11. Is this a multi-site study? *

*Mark only one oval.*

- ( ) Yes
- ( ) No

12. What is the main setting of the study (main location of recruitment)? *

______________________

13. Who are the participants? *

*Tick all that apply.*

- [ ] PD only
- [ ] PD cases and controls
- [ ] Defined PD group, e.g. early onset PD, familial PD, DBS
- [ ] Other: ______________________

14. What is the study context? *

*Mark only one oval.*

- ( ) Observational study
- ( ) Interventional study e.g. drug trial

15. What is the study timespan? *

*Mark only one oval.*

- ( ) Cross-sectional: one assessment only, with no further contact
- ( ) Cross-sectional plus: one assessment with further questionnaires, EPR, Mortality follow-up
- ( ) Longitudinal (prospective): multiple face-to-face assessments
- ( ) Longitudinal (retrospective)

16. Planned follow-up duration and visit intervals

e.g. 5 years and every 6 month. (Skip this question if not longitudinal)

______________________________

17. Main Study Site Region *

*Mark only one oval.*

- ( ) North America
- ( ) South America
- ( ) Europe
- ( ) Africa
- ( ) Asia/Oceania
- ( ) No main region but multi-region

18. Country of main site *

______________________________

19. Publication DOI or webpage describing study design

______________________________

Study Participants

20. Rough breakdown of ancestry - Europeans (%) *

Among those with blood/DNA samples

______________________________

21. Rough breakdown of ancestry - Africans (%) *

Among those with blood/DNA samples

______________________________

22. Rough breakdown of ancestry - Asians (%) *

Among those with blood/DNA samples

______________________________

23. Inclusion criteria: what criteria was used to confirm/diagnose PD cases?

*Mark only one oval.*

- ◯ MDS Diagnostic criteria
- ◯ UK Brain Bank criteria
- ◯ Other: ______________________________

24. Inclusion criteria: what are the disease stage criteria for recruiting PD cases?

E.g. diagnosis < 5 years, symptom onset < 3 years, Hoehn and Yahr stage < 3, no medication etc.

______________________________
______________________________
______________________________
______________________________
______________________________

25. Inclusion criteria: are there any other important criteria for PD cases?
E.g. MMSE<27, no family history etc.

26. What are the inclusion criteria for controls?

27. Are there any other specific recruitment features?

28. What is the percentage (%) with post-mortem confirmation of diagnosis? *
0 - 100

#### Genotyping information

Please answer the following questions if genotyping has done/planned. Depending on the coverage of the genotyping chip, GP2 may not need to re-genotype the samples. Please put NA if the genotyping has not been done/planned.

29. Name and version of genotyping chip if used *

"NA" if genotyping has not been done/planned.

30. Number of SNPs on the chip *

Rough number is fine (e.g. 230K, 510K, 1M). "NA" if genotype has not been done/planned.

#### Currently available data and samples

For the following questions, please answer based on the current availability of data/samples.

31. Number of participants with PD *

Those with the clinical data available and transferrable.

32. Number of PD patients whose DNA is currently available *

If DNA are not extracted from specimen, please DO NOT include them here.

33. Number of PD patients whose genotype is currently available *

If genotyping has been done and can be transferred to NIA. 0 otherwise

34. Number of participants without PD *

Those with the clinical data available and transferrable.

35. Number of non-PD participants whose DNA is currently available *

If DNA are not extracted from specimen, please DO NOT include them here but include then in the following question

36. Number of non-PD participants whose genotype is currently available *

_______________________

37. Shipping current DNA samples *

Samples are going to be genotyped at NIH with a custom array for neurodegenerative disorders. Data and samples will be returned to submitting sites. If it is impossible to send DNA samples to NIH, please check the box below. This will be subject to MTA and collaborative agreements but an initial indication of the situation for your study will be helpful

*Mark only one oval.*

- ( ) We anticipate we will be able to send samples to NIH for genotyping
- ( ) We anticipate we will not be able to send samples to NIH for genotyping
- ( ) We do not know the situation with transferring samples at this time
- ( ) NIH(NIA) should have had all the DNA samples already
- ( ) Other: _______________________

#### Additional data/sample expected to be available by the end of 2022

NIA needs to receive samples by the end of 2022 at the latest. For the ongoing study, please provide your best guess about the additional availability of data and samples by the end of 2022. For the completed study, please provide 0 for all questions in this section.

38. Number of ADDITIONAL PD patients *

_______________________

39. Number of ADDITIONAL PD patients whose DNA will be available by the end of 2022 *

Can Include participants with blood/specimen but DNA have not been extracted yet

_______________________

40. Number of ADDITIONAL PD patients whose genotype will be available by the end of 2022 *

_______________________

41. Number of ADDITIONAL non-PD patients *

42. Number of ADDITIONAL non-PD participants whose DNA will be available by the end of 2022 *

Can Include participants with blood/specimen but whose DNA has not been extracted yet

43. Number of ADDITIONAL non-PD participants whose genotype will be available by the end of 2022 *

44. Shipping ADDITIONAL DNA samples

*Mark only one oval.*

- ◯ We anticipate we will be able to send samples to NIH for genotyping
- ◯ We anticipate we will not be able to send samples to NIH for genotyping
- ◯ We do not know the situation with transferring samples at this time
- ◯ Other: ____________

#### Ethics Approval for Use of Data & Samples

45. Institutional Review/Ethics Review Board

Which ethics committee reviewed and approved this human subjects research?

46. Date of ethics board approval

*Example: 7 January 2019*

47. Data Sharing *

What are the terms of data sharing outlined in the consent documents?

*Mark only one oval.*

- Use limited to study for which they were originally collected
- Use limited to PI/Institution at which study was conducted
- Use limited to Parkinson's Disease reseach
- Consent silent on future use
- Consent explicitly allows sharing of data
- Other: ______________________

48. Sample Sharing *

What are the terms of sample sharing outlined in the consent documents?

*Mark only one oval.*

- Use limited to study for which they were originally collected
- Use limited to PI/Institution at which study was conducted
- Use limited to Parkinson's Disease reseach
- Consent silent on future use
- Consent explicitly allows sharing of samples
- Other: ______________________

49. International Sharing *

Does the consent allow sharing of data and/or samples outside of the country in which the study was performed?

*Mark only one oval.*

- No
- Consent silent on international sharing/storage
- Yes
- Other: ______________________

50. Commercial Use *

Does the consent allow for commercial use of the samples/data?

*Mark only one oval.*

- No
- Consent silent on commercial use
- Yes
- Other: ____________________

51. Data Storage Location *

Does the consent form specify where data will be stored?

*Mark only one oval.*

- Storage location specified in consent
- Consent silent on storage location
- Consent allows storage outside the institution and/or on cloud servers
- Other: ____________________

52. Duration of Storage *

Is the duration of data storage limited?

*Mark only one oval.*

- Limited
- Consent silent on duration of storage
- Unlimited
- Other: ____________________

53. If storage duration is limited, when does it expire?

____________________

*Example: 7 January 2019*

Collected Data

54. Basic clinical variables available for most participants - multiple responses are possible

*Tick all that apply.*

- [ ] Gender/sex
- [ ] Age at diagnosis
- [ ] Age at onset
- [ ] Censored vital status (Alive/Dead with Date)
- [ ] Family history of PD (Basic - present/absent)
- [ ] Family history (Detailed - structure, number of affected family members, c/w AD/AR/X-linked)
- [ ] Education level
- [ ] Self-reporting race
- [ ] Medical history (common/PD related diseases)
- [ ] Diagnostic certainty
- [ ] Time to major PD events (e.g., HY3, wearing-off, dyskinesia)
- [ ] Diagnosis checklist (MDS diagnostic criteria)

55. Other samples available for some participants

*Tick all that apply.*

- [ ] Plasma
- [ ] Serum
- [ ] RNA
- [ ] PBL (Peripheral blood lymphocytes)
- [ ] LCL (Lymphoblastoid cell lines)
- [ ] Skin biopsy
- [ ] iPS
- [ ] Post-mortem brain
- [ ] CSF

56. *Omics Data Availability

*Tick all that apply.*

| | Blood | Brain | Other Tissues |
|---|---|---|---|
| RNAseq | ☐ | ☐ | ☐ |
| Methylation | ☐ | ☐ | ☐ |
| Metabolomics | ☐ | ☐ | ☐ |
| Proteomics | ☐ | ☐ | ☐ |
| Exome/Genome sequence files | ☐ | ☐ | ☐ |

Global Parkinson's Genetics Program

57. Clinical Data Availability *

*Mark only one oval per row.*

| | Cross-sectional | Longitudinal | None |
|---|---|---|---|
| Medication information | ◯ | ◯ | ◯ |
| Global PD severity (e.g. CISI-PD) | ◯ | ◯ | ◯ |
| Lifestyle (smoking, alcohol, income) | ◯ | ◯ | ◯ |
| Environment (residence, work) | ◯ | ◯ | ◯ |
| Head trauma history | ◯ | ◯ | ◯ |
| Hoehn and Yahr | ◯ | ◯ | ◯ |
| UPDRS (original/MDS) | ◯ | ◯ | ◯ |
| Cognitive measurements | ◯ | ◯ | ◯ |
| REM sleep behavior disorder | ◯ | ◯ | ◯ |
| Autonomic function | ◯ | ◯ | ◯ |
| Depression/anxiety measurement | ◯ | ◯ | ◯ |
| Sleep assessment | ◯ | ◯ | ◯ |
| Vitals (Height, weight, BMI, BP) | ◯ | ◯ | ◯ |
| Orthostatic hypotension | ◯ | ◯ | ◯ |
| Smell assessment | ◯ | ◯ | ◯ |
| Activities of daily living (e.g. Schwab & England) | ◯ | ◯ | ◯ |
| Quality of life (e.g. PDQ39/PDQ8) | ◯ | ◯ | ◯ |
| Pain assessment | ◯ | ◯ | ◯ |
| Blood CBC, Chem | ◯ | ◯ | ◯ |

58. Imaging data availability *

*Mark only one oval per row.*

| | Cross-sectional | Longitudinal | None |
|---|---|---|---|
| MRI | ◯ | ◯ | ◯ |
| DATSCAN | ◯ | ◯ | ◯ |

59. Pathology availability *

Is pathology available for more than half (PD) participants?

*Mark only one oval.*

◯ Yes

◯ No

60. Free note (describe any additional data types you can provide)

______________________________

______________________________

______________________________

______________________________

______________________________

61. Are you able to recontact patients if available for further follow-up/research studies? *

*Mark only one oval.*

◯ Yes

◯ No

Additional questions

62. Are you also interested in joining the GP2 monogenic hub?

The GP2 monogenic hub will collate possible monogenic cases. If you are interested, please indicate here and you will be recontacted by the Monogenic team

*Mark only one oval.*

- ◯ Yes
- ◯ No

63. In publication for research using GP2 data, do you support:

Transparency, Collaboration, and Open Science is GP2's policy goal. We would like to know your general position on this. Multiple responses possible

*Tick all that apply.*

- ☐ Open data
- ☐ Open code (analytical scripts)
- ☐ Publishing with CC-BY license AND immediate free access through PubMed Central

64. Would you like to request analytical support from the GP2 data analysis team?

*Tick all that apply.*

- ☐ Yes
- ☐ Not sure
- ☐ No

65. Please send the consent form, protocol and case report form to

And any comments here.

_______________________________________________

_______________________________________________

_______________________________________________

_______________________________________________

_______________________________________________

### 4. Map of contributing cohorts and genotyping tracker

Below is a map of the geographic distribution of cohorts and the progress of genotyping (as of 15th May 2022). The live map is regularly updated and can be viewed on the GP2 website at https://gp2.org/cohort-dashboard/.

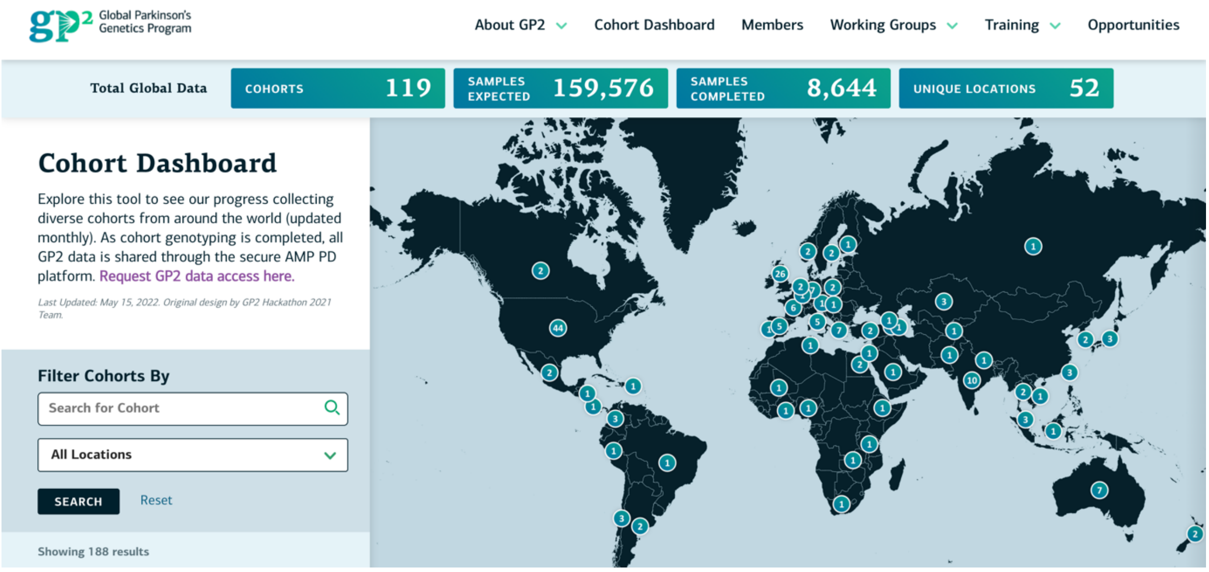

Details of institutions contributing PD cohorts to GP2 can be found on the GP2 webpage at https://gp2.org/members/

### 5. Supplementary Table 1. Pathology Data Elements

| Modality | Item | Description | ItemType | Required | Values | Comments |
|---|---|---|---|---|---|---|
| Base | participant_id | participant's ID | string | required | | |
| Base | asap_id | Unique participant ID asigned by ASAP | string | required | | |
| Pathology | path_brain_id | Brain Bank Subject ID (Unique Subject ID at Brain Bank) | string | nullable | | |
| Demographics | date_birth_unix | NINDS CDE for Parkinson's Disease: Date of Birth (Unix date) If not provided days or months, we regard this as the middle of the month (15) or the year (July/30) | numeric | nullable | (y>= -21914 ) & (y<=51535) | |
| Demographics | sex | NINDS CDE for Parkinson's Disease: Sex assigned at birth | string | nullable | ["Male", "Female", "Intersex", "Unknown", "Other", "Not Reported"] | |
| Demographics | ethnicity | NINDS CDE for Parkinson's Disease: Ethnicity | string | nullable | ["Hispanic or Latino", "Not Hispanic or Latino", "Unknown", "Not Reported"] | |
| Demographics | race | NINDS CDE for Parkinson's Disease: Race category + | string | nullable | ["American Indian or Alaska Native", "Asian", "White", "Black or African American", "Multi-racial", "Native Hawaiian or Other Pacific Islander", "Other", "Unknown", "Not Reported"] | |
| Family history | family_hx_1st_PD | Did your parents, full-siblings, or children have Parkinson's disease? | string | nullable | ["Yes", "No"] | |
| Diagnosis | primary_diagnosis | Most likely primary neurodegeneralive diagnosis (clinical) | string | required | ["Idiopathic PD", "Alzheimer's disease", "Frontotemporal dementia", "Corticobasal syndrome", "Dementia with Lewy bodies", "Dopa-responsive dystonia", "Essential tremor", "Hemiparkinson/hemiatrophy syndrome", "Juvenile autosomal recessive parkinsonism", "Motor neuron disease with parkinsonism", "Multiple system atrophy", "Neuroleptic-induced parkinsonism", "Normal pressure hydrocephalus", "Progressive supranuclear palsy", "Psychogenic parkinsonism", "Vascular parkinsonism", "No PD nor other neurological disorder", "Spinocerebellar Ataxia (SCA)", "Prodromal non-motor PD", "Prodromal motor PD", "Other neurological disorder"] | |
| Diagnosis | primary_diagnosis_text | If "97:Other neurological disorders(s) (specify)" please specify here (baseline) | string | nullable | | |
| PD History | age_at_onset | Age at onset (any symptom) | numeric | nullable | (y>=0) & (y<=120) | |
| PD History | age_at_first_motor_symptom | Age at first motor symptom | numeric | nullable | (y>=0) & (y<=120) | |
| PD History | first_motor_symptom | First motor symptom. pick all that apply and separate with "\|". (e.g, 1, 1\|3 or 2\|3\|5) | string | nullable | {1:"tremor", 2:"micrographia", 3:"stiffness/frozen shoulder", 4:"Impaired manual dexterity", 5:"Gait disorder", 6:"General slowing up", 7:"Other"} | |
| PD History | age_at_diagnosis | Age at diagnosis | numeric | nullable | (y>=0) & (y<=120) | |
| Medical History | hx_dementia_mci | Have you ever been diagnosed with Dementia or MCI? | string | nullable | ["Yes", "No"] | |
| Medical History | hx_melanoma | Have you ever been diagnosed with Melanoma | string | nullable | ["Yes", "No"] | |
| PD History | living_status_at_censoring | Event is death. 0=right censored (not yet) 1= event at the "age" | numeric | nullable | ["0","1","2"] | Event will be 1 for all brain bank cases |
| Pathology | path_year_death | year of death | numeric | nullable | (y>1920)&(y<2050) | |
| Pathology | age_at_death | expired_age (Age at Death) | string | nullable | | |
| Pathology | cause_death | Cause of death | string | nullable | | |
| Pathology | path_PMI_hours | Interval between death and autopsy start (hours) | numeric | nullable | (y>0)&(y<1200) | |
| Pathology | pm_RIN | Post mortem RIN | | nullable | (0-10) | |
| Pathology | pm_PH | Post mortem Ph | | nullable | (0-14) | |
| Pathology | brain_weight | Brain weight | numeric | nullable | grams | |
| Pathology | path_autopsy_dx_main | Pathological diagnosis | string | required | ["Lewy body disease nos", "Parkinson's disease", "Parkinson's disease with dementia", "Dementia with Lewy bodies", "Multiple system atrophy (SND>OPCA)", "Multiple system atrophy (OPCA<SND)", "Multiple system atrophy (SND=OPCA)", "Progressive supranuclear palsy", "Corticobasal degeneration", "Globular glial tauoapathy (GGT)", "Chronic traumatic encephalopathy (CTE)", "FTLD-Tau (Pick's)", "FTLD-Tau (MAPT)", "FTLD-Tau (AGD)", "FTLD-TDP43, Type A", "FTLD-TDP43, Type B", "FTLD-TDP43, Type C", "FTLD-TDP43, Type D", "FTLD-TDP43, Type E", "Motor neurone disease-TDP43 (MND or ALS)", "FTLD-MND-TDP43", "Huntington's disease", "Spinocerebellar ataxia, nos", "Prion disease, nos", "Alzheimer's disease (high level neuropathological change)", "Alzheimer's disease (intermediate level neuropathological change)", "Control, Low level AD neuropathological change", "Control, Limbic predominant age-related TDP43 proteinopathy (LATE)", "Control, Argyrophilic grain disease", "Control, Primary age-related tauopathy (PART)", "Control, Ageing-related tau astrogliopathy (ARTAG)", "Control, Cerebrovascular disease (atherosclerosis)", "Control, Cerebrovascular disease (hyaline arteriolosclerosis)", "Control, Cerebrovascular disease (cerebral amyloid angiopathy)", "Control, no misfolded protein or significant vascular pathology", "Other neurological disorder"] | |
| Pathology | path_autopsy_dx_brain_bank | Pathological diagnosis | string | required | | Recorded as it is from data given by brain bank |
| Pathology | path_braak_nft | Braak Tangle stage | string | nullable | ["0", "I", "II", "III", "IV", "V", "VI", "I/II", "III/IV", "IV/V", "V/VI"] | |
| Pathology | path_braak_asyn | Braak LB stage | string | nullable | ["0", "1", "2", "3", "4", "5", "6", "1/2", "3/4", "4/5", "5/6"] | |
| Pathology | path_cerad | Semiquantitative assessment of neuritic plaques | string | nullable | ["None", "Sparse", "Moderate", "Frequent"] | |
| Pathology | path_thal | thal stage: Amyloid beta | string | nullable | ["0", "1", "2", "3", "4", "5", "1/2", "3", "4/5"] | |
| Pathology | path_mckeith | McKeith and USSLBD Stage | string | nullable | ["Neocortical", "Limbic (transitional)" ,"Brainstem", "Amygdala Predominant", "Olfactory bulb only"] | |
| Pathology | path_sn_neuronal_loss | sn_neuronal_loss (none, mild, moderate, severe, 0-3) | numeric | nullable | ["None", "Mild", "Moderate", "Severe", "Not assessed", "Unknown"] | |
| Pathology | path_infarcts | Cerebral infarcts (indicator for a large infarct or in study region) | string | nullable | ['Yes", "No"] | |
| Pathology | path_nia_ri | NIA_RI criteria | string | nullable | ['low', 'intermediate', 'high', 'none'] | |
| Pathology | path_nia_aa | NIA_AA criteria (pathological. not biomarker criteria) | string | nullable | ["A1","A2","A3"] | |
| Pathology | path_nia_aa | NIA_AA criteria (pathological. not biomarker criteria) | string | nullable | ["B1","B2","B3"] | |
| Pathology | path_nia_aa | NIA_AA criteria (pathological. not biomarker criteria) | string | nullable | ["C1","C2","C3"] | |
| Pathology | known_pathogenic_mutation | MutationNOS (any known relevant mutation or polymorphisms, name?) | string | nullable | | |
| Pathology | arteriolosclerosis_severity_scale | Arteriolosclerosis severity scale | string | nullable | ["None", "Mild", "Moderate", "Severe", "Not assessed", "Unknown"] | |
| Pathology | atherosclerosis | Atherosclerosis severity scale | string | nullable | ["None", "Mild", "Moderate", "Severe", "Not assessed", "Unknown"] | |
| Pathology | amyloid_angiopathy_severity_sca | Amyloid angiopathy severity scale | string | nullable | ["None", "Mild", "Moderate", "Severe", "Not assessed", "Unknown"] | |

### 6. Supplementary Table 2. GP2 Core Data Dictionary

| Single measure | Modality | no. | Item | Description | ItemType | Required | Values | AMP_PD_item | Conversion | Comment |
|---|---|---|---|---|---|---|---|---|---|---|
| 1 | Base | 1 | **participant_id** | participant's ID | string | required | | participant_id | identical | |
| 0 | Base | 2 | **visit_name** | visit name: screening, baseline, V01, V02 etc. baseline is when the participants were enrolled | string | required | | visit_name | identical | |
| 0 | Base | 3 | **visit_month** | month from baseline visit (longitudinal study. Negative value is possible for screening visits), rounded integer | integer | nullable | (y>=-1200) & (y<=1200) | visit_month | identical | planned month |
| 0 | Base | | **date_visit_unix** | date of visit in UNIX DATE (days from 1970-01-01) If not provided days or months, we regard this as the middle of the month (15) or the year (July/30) | numeric | nullable | (y>= -21914 ) & (y<=51535) | | | |
| 1 | Base | | **date_baseline_unix** | date of baseline in UNIX DATE (days from 1970-01-01) If not provided days or months, we regard this as the middle of the month (15) or the year (July/30) | numeric | nullable | (y>= -21914 ) & (y<=51535) | | | |
| 1 | Base | 4 | age_at_baseline | age at baseline visit | numeric | nullable | (y>=0) & (y<=125) | age_at_baseline | identical | |
| 1 | Demographics | 1 | date_enrollment | date of enrollment (YYYY/MM) | string | nullable | | | | |
| 1 | Demographics | 2 | date_birth_unix | NINDS CDE for Parkinson's Disease: Date of Birth (Unix date) If not provided days or months, we regard this as the middle of the month (15) or the year (July/30) | numeric | nullable | (y>= -21914 ) & (y<=51535) | | | |
| 1 | Demographics | 3 | sex | NINDS CDE for Parkinson's Disease: Sex assigned at birth | string | required | ["Male", "Female", "Intersex", "Unknown", "Other", "Not Reported"] | sex | convertible | |
| 1 | Demographics | 4 | ethnicity | NINDS CDE for Parkinson's Disease: Ethnicity | string | required | ["Hispanic or Latino", "Not Hispanic or Latino", "Unknown", "Not Reported"] | ethnicity | convertible | |
| 1 | Demographics | 5 | race | NINDS CDE for Parkinson's Disease: Race category + | string | required | ["American Indian or Alaska Native", "Asian", "White", "Black or African American", "Multi-racial", "Native Hawaiian or Other Pacific Islander", "Other", "Unknown", "Not Reported"] | race | convertible | |
| 1 | Demographics | 6 | education_years | NINDS CDE for Parkinson's Disease: Number of years of education | numeric | nullable | (y>=0) & (y<=40) | education_level_years | convertible | |
| 1 | Demographics | 7 | education_level | Derived value for level of education | string | nullable | ["<High School", "High School/GED", "Some college without degree", "Associate degree college", "Bachelor's degree", "Master's degree", "Professional or doctoral degree", "Refuse", "Other"] | | | |
| 1 | Family history | 1 | family_hx_1st_PD | Did your parents, full-siblings, or children have Parkinson's disease? | string | nullable | ["Yes", "No"] | | | biological_mot |
| 1 | Family history | 2 | family_hx_2nd_PD | Did your parents, grand parents, full/half-siblings, uncles, aunts have Parkinson's disease? | string | nullable | ["Yes", "No"] | "Refused", "Don't | | |
| 1 | Family history | 3 | family_hx_generations_PD | Are there any people in your relatives with Parkinson's disease more than one generation? | string | nullable | ["Yes", "No"] | | | |
| 1 | Family history | 4 | family_hx_n_of_PD | How many people in the family including the proband are affected with PD | numeric | nullable | (y>=0) & (y<=100) | | | |
| 1 | Enrollment Category | 1 | study_arm | Study Arm: case/contro/positive control/affected/unaffected, placebo/control. etc should be defined at enrollment | string | required | | study_arm | identical | |
| 1 | Enrollment Category | 1 | Phenotype | Phenotype for sample sheet (basically derived from study_arm) | string | required | ["PD", "Control", "Prodromal", "Other", "Not Reported"] | | | |
| 1 | Diagnosis | 1 | primary_diagnosis | Most likely primary neurodegeneralive diagnosis (baseline) | string | nullable | ["Idiopathic PD", "Alzheimer's disease", "Frontotemporal dementia", "Corticobasal syndrome", "Dementia with Lewy bodies", "Dopa-responsive dystonia", "Essential tremor", "Hemiparkinson/hemiatrophy syndrome", "Juvenile autosomal recessive parkinsonism", "Motor neuron disease with parkinsonism", "Multiple system atrophy", "Neuroleptic-induced parkinsonism", "Normal pressure hydrocephalus", "Progressive supranuclear palsy", "Psychogenic parkinsonism", "Vascular parkinsonism", "No PD nor other neurological disorder", "Spinocerebellar Ataxia (SCA)", "Prodromal non-motor PD", "Prodromal motor PD", "Other neurological disorder"] | | | |
| 1 | Diagnosis | 2 | primary_diagnosis_text | If "97:Other neurological disorders(s) (specify)" please specify here (baseline) | string | nullable | | | | |
| 1 | Diagnosis | 3 | code_primary_diagnosis_confide | What is your percent confidence in your most likely primary diagnosis? (baseline) | string | nullable | ["0-25%", "26-50%", "51-75%", "76-100%"] | | | |
| 1 | Diagnosis | 4 | last_diagnosis | Lastest diagnosis or autopsy diagnosis | string | nullable | ["Idiopathic PD", "Alzheimer's disease", "Frontotemporal dementia", "Corticobasal syndrome", "Dementia with Lewy bodies", "Dopa-responsive dystonia", "Essential tremor", "Hemiparkinson/hemiatrophy syndrome", "Juvenile autosomal recessive parkinsonism", "Motor neuron disease with parkinsonism", "Multiple system atrophy", "Neuroleptic-induced parkinsonism", "Normal pressure hydrocephalus", "Progressive supranuclear palsy", "Psychogenic parkinsonism", "Vascular parkinsonism", "No PD nor other neurological disorder", "Spinocerebellar Ataxia (SCA)", "Prodromal non-motor PD", "Prodromal motor PD", "Other neurological disorder"] | | | |
| 1 | Diagnosis | 5 | visit_month_of_last_diagnosis | Months from the baseline to the visit confirming the latest diagnosis (or the date of death when path confirmed) | numeric | nullable | (y>=-1200) & (y<=1200) | | | |
| 1 | Diagnosis | 6 | diagnosis_change | Had the diagnosis changed during the follow-up? (Mostly important for prodromal cohort) | string | nullable | ["Yes", "No"] | | | |
| 1 | Diagnosis | 7 | visit_month_diagnosis_change | Months from the baseline to the change of diagnosis (to the latest diagnosis) | numeric | nullable | (y>=-1200) & (y<=1200) | | | |
| 1 | Diagnosis | 3 | code_last_diagnosis_confidence | What is your percent confidence in your most likely last diagnosis? | string | nullable | ["0-25%", "26-50%", "51-75%", "76-100%"] | | | |
| 1 | Diagnosis | 8 | handedness | Right handedness or left handedness? | string | nullable | ["Right", "Left", "Mixed"] | | | |
| 1 | Diagnosis | 9 | dominant_side | Side predominantly affected at onset | string | nullable | ["Right", "Left", "Symmetric"] | | | |
| 1 | Known_relatedness | 1 | related_individuals_reported | {id: relationship). e.g. {"id-011": "Mother", "id-012":"Father", "id-019":Full-sibling} | string | nullabe | | | | |
| 0 | Vital | 1 | weight_kg | in kg | numeric | nullable | (y>=20) & (y<=225) | | | |
| 0 | Vital | 1 | height_cm | in cm | numeric | nullable | (y>=100) & (y<=220) | | | |
| 0 | Vital | 1 | BMI | | numeric | nullable | (y>=10) & (y<=75) | | | |
| 0 | Vital | 1 | heart_rate | per minute | numeric | nullable | (y>=35) & (y<=200) | | | |
| 0 | Vital | 1 | systolic_blood_pressure | in mmHg (use only if no information is given on sitting, standing, or supine positioning) | numeric | nullable | (y>=50) & (y<=250) | | | |
| 0 | Vital | 1 | diastolic_blood_pressure | in mmHg (use only if no information is given on sitting, standing, or supine positioning) | numeric | nullable | (y>=30) & (y<=250) | | | |
| 1 | MDS_Diagnostic_criteria | 1 | dx_essential_bradykinesia | essential criterion: required | string | nullable | ["Yes", "No"] | | | |
| 1 | MDS_Diagnostic_criteria | 2 | dx_essential_rest_tremor | essential criterion: either rest_tremor or rigidity required | string | nullable | ["Yes", "No"] | | | |
| 1 | MDS_Diagnostic_criteria | 3 | dx_essential_rigidity | essential criterion: either rest_tremor or rigidity required | string | nullable | ["Yes", "No"] | | | |
| 1 | MDS_Diagnostic_criteria | 4 | dx_supportive_1_positive_drug_response | 1. Clear and dramatic beneficial response to dopaminergic therapy. During initial treatment, patient returned to normal or near-normal level of function. In the absence of clear documentation of initial response a dramatic response can be classified as: a) Marked improvement with dose increases or marked worsening with dose decreases. Mild changes do not qualify. Document this either objectively (>30% in UPDRS III with change in treatment), or subjectively (clearly-documented history of marked changes from a reliable patient or caregiver). b) Unequivocal and marked on/off fluctuations, which must have at some point included predictable end-of-dose wearing off. | string | nullable | ["Yes", "No"] | | | |

#### Supplementary Table 2. GP2 Core Data Dictionary (continued)

| Single measure | Modality | no. | Item | Description | ItemType | Required | Values | AMP_PD_item | Conversion | Comment |
|---|---|---|---|---|---|---|---|---|---|---|
| 1 | MDS_Diagnostic_criteria | 5 | dx_supportive_2_levodopa_induced_dyskinesia | 2. Presence of levodopa-induced dyskinesia | string | nullable | ["Yes", "No"] | | | |
| 1 | MDS_Diagnostic_criteria | 6 | dx_supportive_3_rest_tremor | 3. Rest tremor of a limb, documented on clinical examination (in past, or on current examination) | string | nullable | ["Yes", "No"] | | | |
| 1 | MDS_Diagnostic_criteria | 7 | dx_supportive_4_olfactory_loss_or_MIBG | 4. The presence of either olfactory loss or cardiac sympathetic denervation on MIBG scintigraphy | string | nullable | ["Yes", "No"] | | | |
| 1 | MDS_Diagnostic_criteria | 8 | dx_exclusion_1_cerebellar_signs | 1. Unequivocal cerebellar abnormalities, such as cerebellar gait, limb ataxia, or cerebellar oculomotor abnormalities (eg, sustained gaze evoked nystagmus, macro square wave jerks, hypermetric saccades) | string | nullable | ["Yes", "No"] | | | |
| 1 | MDS_Diagnostic_criteria | 9 | dx_exclusion_2_abnormal_eye_movement | 2. Downward vertical supranuclear gaze palsy, or selective slowing of downward vertical saccades | string | nullable | ["Yes", "No"] | | | |
| 1 | MDS_Diagnostic_criteria | 10 | dx_exclusion_3_FTD_5y | 3. Diagnosis of probable behavioral variant frontotemporal dementia or primary progressive aphasia, defined according to consensus criteria31 within the first 5 y of disease | string | nullable | ["Yes", "No"] | | | |
| 1 | MDS_Diagnostic_criteria | 11 | dx_exclusion_4_lower_limbs_only_3y | 4. Parkinsonian features restricted to the lower limbs for more than 3 y | string | nullable | ["Yes", "No"] | | | |
| 1 | MDS_Diagnostic_criteria | 12 | dx_exclusion_5_drug_induced_parkinsonism | 5. Treatment with a dopamine receptor blocker or a dopamine-depleting agent in a dose and time-course consistent with drug-induced parkinsonism | string | nullable | ["Yes", "No"] | | | |
| 1 | MDS_Diagnostic_criteria | 13 | dx_exclusion_6_no_drug_response | 6. Absence of observable response to high-dose levodopa despite at least moderate severity of disease | string | nullable | ["Yes", "No"] | | | |
| 1 | MDS_Diagnostic_criteria | 14 | dx_exclusion_7_cortical_signs | 7. Unequivocal cortical sensory loss (ie, graphesthesia, stereognosis with intact primary sensory modalities), clear limb ideomotor apraxia, or progressive aphasia | string | nullable | ["Yes", "No"] | | | |
| 1 | MDS_Diagnostic_criteria | 15 | dx_exclusion_8_negative_neuroimaging | 8. Normal functional neuroimaging of the presynaptic dopaminergic system | string | nullable | ["Yes", "No"] | | | |
| 1 | MDS_Diagnostic_criteria | 16 | dx_exclusion_9_other_parkinsonism | 9. Documentation of an alternative condition known to produce parkinsonism and plausibly connected to the patient's symptoms, or, the expert evaluating physician, based on the full diagnostic assessment feels that an alternative syndrome is more likely than PD | string | nullable | ["Yes", "No"] | | | |
| 1 | MDS_Diagnostic_criteria | 17 | dx_redflag_01_gait_impairment_5y | 1. Rapid progression of gait impairment requiring regular use of wheelchair within 5 y of onset | string | nullable | ["Yes", "No"] | | | |
| 1 | MDS_Diagnostic_criteria | 18 | dx_redflag_02_progression_absent | 2. A complete absence of progression of motor symptoms or signs over 5 or more y unless stability is related to treatment | string | nullable | ["Yes", "No"] | | | |
| 1 | MDS_Diagnostic_criteria | 19 | dx_redflag_03_severe_bulbar_dysfunction_5y | 3. Early bulbar dysfunction: severe dysphonia or dysarthria (speech unintelligible most of the time) or severe dysphagia (requiring soft food, NG tube, or gastrostomy feeding) within first 5 y | string | nullable | ["Yes", "No"] | | | |
| 1 | MDS_Diagnostic_criteria | 20 | dx_redflag_04_inspiratory_respiratory_dysfunction | 4. Inspiratory respiratory dysfunction: either diurnal or nocturnal inspiratory stridor or frequent inspiratory sighs | string | nullable | ["Yes", "No"] | | | |
| 1 | MDS_Diagnostic_criteria | 21 | dx_redflag_05_severe_autonomic_failure_5y | 5. Severe autonomic failure in the first 5 y of disease. This can include:<br>a) Orthostatic hypotension32—orthostatic decrease of blood pressure within 3 min of standing by at least 30 mm Hg systolic or 15 mm Hg diastolic, in the absence of dehydration, medication, or other diseases that could plausibly explain autonomic dysfunction, or<br>b) Severe urinary retention or urinary incontinence in the first 5 y of disease (excluding long-standing or small amount stress incontinence in women), that is not simply functional incontinence. In men, urinary retention must not be attributable to | string | nullable | ["Yes", "No"] | | | |
| 1 | MDS_Diagnostic_criteria | 22 | dx_redflag_06_recurrent_falls_3y | 6. Recurrent (>1/y) falls because of impaired balance within 3 y of onset | string | nullable | ["Yes", "No"] | | | |
| 1 | MDS_Diagnostic_criteria | 23 | dx_redflag_07_dysproportionate_dystonia | 7. Disproportionate anterocollis (dystonic) or contractures of hand or feet within the first 10 y | string | nullable | ["Yes", "No"] | | | |
| 1 | MDS_Diagnostic_criteria | 24 | dx_redflag_08_no_nonmotor_symptoms_5y | 8. Absence of any of the common nonmotor features of disease despite 5 y disease duration. These include sleep dysfunction (sleep-maintenance insomnia, excessive daytime somnolence, symptoms of REM sleep behavior disorder), autonomic dysfunction (constipation, daytime urinary urgency, symptomatic orthostasis), hyposmia, or psychiatric dysfunction (depression, anxiety, or hallucinations) | string | nullable | ["Yes", "No"] | | | |
| 1 | MDS_Diagnostic_criteria | 25 | dx_redflag_09_unexplained_pyramidal_tract_signs | 9. Otherwise-unexplained pyramidal tract signs, defined as pyramidal weakness or clear pathologic hyperreflexia (excluding mild reflex asymmetry and isolated extensor plantar response) | string | nullable | ["Yes", "No"] | | | |
| 1 | MDS_Diagnostic_criteria | 26 | dx_redflag_10_symmetric_parkinsonism | 10. Bilateral symmetric parkinsonism. The patient or caregiver reports bilateral symptom onset with no side predominance, and no side predominance is observed on objective examination | string | nullable | ["Yes", "No"] | | | |
| 1 | MDS_Diagnostic_criteria | 27 | dx_criteria_application | Apply the criteria | string | nullable | ["Clinically established PD", "Probable PD", "Other", 'Unknown"] | | | |
| 1 | PD History | | age_at_onset | Age at onset (any symptom) | numeric | nullable | (y>=0) & (y<=120) | | | |
| 1 | PD History | 1 | age_at_first_motor_symptom | Age at first motor symptom | numeric | nullable | (y>=0) & (y<=120) | | | |
| 1 | PD History | 2 | first_motor_symptom | First motor symptom. pick all that apply and separate with "\|". (e.g, 1, 1\|3 or 2\|3\|5) | string | nullable | {1:"tremor", 2:"micrographia", 3:"stiffness/frozen shoulder", 4:"Impaired manual dexterity", 5:"Gait disorder", 6:"General slowing up", 7:"Other"} | | | |
| 1 | PD History | 3 | age_at_diagnosis | Age at diagnosis | numeric | nullable | (y>=0) & (y<=120) | | | |
| 1 | PD History | 19 | living_status_at_censoring | Event is death. 0=right censored (not yet) 1= event at the "age" | numeric | nullable | y.isin([0,1,2]) | | | |
| 1 | PD History | 20 | age_at_living_status_censored | age at event (status=1), age at last obs (status=0), age at first obs (status=2) | numeric | nullable | (y>=0) & (y<=120) | | | |
| 1 | PD History | 21 | hy3_status_at_censoring | Event is reaching hy 3 or higher. 0=right censored (not yet) 1= event at the "age", 2=left censored (already) | numeric | nullable | y.isin([0,1,2]) | | | |
| 1 | PD History | 22 | age_at_hy3_status_censored | age at event (status=1), age at last obs (status=0), age at first obs (status=2) | numeric | nullable | (y>=0) & (y<=120) | | | |
| 1 | PD History | 23 | dyskinesia_status_at_censoring | Event is dyskinesia occurance. 0=right censored (not yet) 1= event at the "age", 2=left censored (already) | numeric | nullable | y.isin([0,1,2]) | | | |
| 1 | PD History | 24 | age_at_dyskinesia_status_censored | age at event (status=1), age at last obs (status=0), age at first obs (status=2) | numeric | nullable | (y>=0) & (y<=120) | | | |
| 1 | PD History | 25 | motor_fluctuation_status_at_censoring | Event is motor fluctuation occurance. 0=right censored (not yet) 1= event at the "age", 2=left censored (already) | numeric | nullable | y.isin([0,1,2]) | | | |
| 1 | PD History | 26 | age_at_motor_fluctuation_status_censored | age at event (status=1), age at last obs (status=0), age at first obs (status=2) | numeric | nullable | (y>=0) & (y<=120) | | | |
| 1 | PD History | 27 | levodopa_status_at_censoring | Event is initiation of levodopa. 0=right censored (not yet) 1= event at the "age", 2=left censored (already) | numeric | nullable | y.isin([0,1,2]) | | | |
| 1 | PD History | 28 | age_at_levodopa_status_censored | age at event (status=1), age at last obs (status=0), age at first obs (status=2) | numeric | nullable | (y>=0) & (y<=120) | | | |
| 1 | PD History | 27 | agonist_status_at_censoring | Event is initiation of agonist. 0=right censored (not yet) 1= event at the "age", 2=left censored (already) | numeric | nullable | y.isin([0,1,2]) | | | |
| 1 | PD History | 28 | age_at_agonist_status_censored | age at event (status=1), age at last obs (status=0), age at first obs (status=2) | numeric | nullable | (y>=0) & (y<=120) | | | |
| 1 | PD History | 29 | pdmed_status_at_censoring | Event is initiation of any PD medication. 0=right censored (not yet) 1= event at the "age", 2=left censored (already) | numeric | nullable | y.isin([0,1,2]) | | | |
| 1 | PD History | 30 | age_at_pdmed_status_censored | age at event (status=1), age at last obs (status=0), age at first obs (status=2) | numeric | nullable | (y>=0) & (y<=120) | | | |
| 1 | PD History | 27 | dementia_status_at_censoring | Event is the first dimentia diagnosis (not symptom). 0=right censored (not yet) 1= event at the "age", 2=left censored (already) | | | | | | |
| 1 | PD History | 28 | age_at_dementia_status_censored | age at event (status=1), age at last obs (status=0), age at first obs (status=2) | | | | | | |
| 1 | PD History | 19 | age_brain_surgery | age at brain surgery | numeric | nullable | (y>=0) & (y<=120) | | | |
| 1 | PD History | 20 | brain surgery type | type of brain surgery | string | nullable | ["DBS-STN", "DBS-GP", "DBS-VIM", "DBS-Other", "DBS-Unknown", "Pallidotomy (RF/RS)", "UltraSound VIM", "UltraSound STN", "UltraSound GP", "Other"] | | | |

Supplementary Table 2. GP2 Core Data Dictionary (continued)

| Single measure | Modality | no. | Item | Description | ItemType | Required | Values | AMP_PD_item | Conversion |
|---|---|---|---|---|---|---|---|---|---|
| 0 | CISI-PD | 1 | code_cisi_pd_motor | CISI-PD motor signs = {0:Normal, 1:Very Mild, 2:Mile, 3:Mild to moderate, 4:Moderate, 5:Severe, 6:Very severe} | integer | nullable | y.isin([0, 1, 2, 3, 4, 5, 6]) | | |
| 0 | CISI-PD | 2 | code_cisi_pd_disability | CISI-PD disability = {0:Normal, 1:Minimal slowness and/or clumsiness, 2:Slowness and/or clumsiness; no limitations, 3: Limitation for demanding activities; does not need help for basic ADL, 4: Limitation to perform basic ADL; help is required for some basic ADL, 5:Great limitation to perform basic ADL; help is requred most or all basic ADL, 6:Severly disabled; helpless; complete assistance needed} | integer | nullable | y.isin([0, 1, 2, 3, 4, 5, 6]) | | |
| 0 | CISI-PD | 3 | code_cisi_pd_motor_complications | CISI-PD motor complications (dyskinesia and fluctuations) = {0:Normal, 1:Very Mild, 2:Mile, 3:Mild to moderate, 4:Moderate, 5:Severe, 6:Very severe} | integer | nullable | y.isin([0, 1, 2, 3, 4, 5, 6]) | | |
| 0 | CISI-PD | 4 | code_cisi_pd_cognitive | CISI-PD cognitive status = {0:Normal, 1:Slowness and/or minimal cognitive problems, 2:Mild cognitie problems; no limitations, 3:Mild to moderate cognitive problems; does not need help for basic ADL, 4:Moderate cognitive problems; help is require for some basic ADL, 5:Severe cognitive problems; help is required for most or all basic ADL, 6:Severely disabled; helpless; complete assistance needed} | integer | nullable | y.isin([0, 1, 2, 3, 4, 5, 6]) | | |
| 1 | Medical | 1 | hx_hypertension | Have you ever been diagnosed with Hypertension/High blood pressure? | string | nullable | ["Yes", "No"] | | |
| 1 | Medical | 2 | hx_hyperlipidemia | Have you ever been diagnosed with Hyperlipidemia/High cholesterol? | string | nullable | ["Yes", "No"] | | |
| 1 | Medical | 3 | hx_diabetes | Have you ever been diagnosed with Diabetes? | string | nullable | ["Yes", "No"] | | |
| 1 | Medical | 4 | hx_myocardinal_infarction | Have you ever been diagnosed with Myocardial infarction/coronary thrombosis/heart attack? | string | nullable | ["Yes", "No"] | | |
| 1 | Medical | 5 | hx_congestive_heart_failure | Have you ever been diagnosed with Congestive heart failure | string | nullable | ["Yes", "No"] | | |
| 1 | Medical | 6 | hx_heart_disease | Have you ever been diagnosed with Heart disease? (MI, Angina, Af, AF) | string | nullable | ["Yes", "No"] | | |
| 1 | Medical | 7 | hx_dementia_mci | Have you ever been diagnosed with Dementia or MCI? | string | nullable | ["Yes", "No"] | | |
| 1 | Medical | 8 | hx_stroke | Have you ever been diagnosed with Stroke/TIA | string | nullable | ["Yes", "No"] | | |
| 1 | Medical | 9 | hx_melanoma | Have you ever been diagnosed with Melanoma | string | nullable | ["Yes", "No"] | | |
| 1 | Medical | 10 | hx_cancer | Have you ever been diagnosed with Cancer (Other than melanoma) -> any cancer | string | nullable | ["Yes", "No"] | | |
| 1 | Medical | | hx_crohn | Have you ever been diagnosed with Crohn's disease | string | nullable | ["Yes", "No"] | | |
| 1 | Medical | 11 | hx_autoimmune_disease | Have you ever been diagnosed with autoimmune disease? (Includes rheumatoid arthritis, lupus) | string | nullable | ["Yes", "No"] | | |
| 1 | Medical | 12 | hx_depression | Have you ever been diagnosed with depression | string | nullable | ["Yes", "No"] | | |
| 1 | Medical | 13 | hx_schizophrenia | Have you ever been diagnosed with schizophrenia | string | nullable | ["Yes", "No"] | | |
| 1 | Medical | 14 | hx_bipolar | Have you ever been diagnosed with bipolar disorder | string | nullable | ["Yes", "No"] | | |
| 1 | Medical | 15 | hx_restless_legs_syndrome | Have you ever been diagnosed with restless legs syndrome | string | nullable | ["Yes", "No"] | | |
| 1 | Medical | 16 | hx_head_trauma | Have you ever had a head injury or concussion? | string | nullable | ["Yes", "No"] | | |
| 1 | Medical | 17 | hx_anxiety | Have you ever been diagnosed with anxiety disorder | string | nullable | ["Yes", "No"] | | |
| 1 | Medical | 18 | hx_constipation | Have you ever diagnosed with constipation | string | nullable | ["Yes", "No"] | | |
| 1 | Medical | 19 | hx_sleep | Have you ever diagnosed with sleeping problems? | string | nullable | ["Yes", "No"] | | |
| 1 | Lifestyle | 1 | smoking_status | Smoking status (Can ignore non-substantial smoking) | string | nullable | "Unknown"] | | |
| 1 | Lifestyle | 2 | smoking_ever_smoker_yes | Smoking status 2 (Can ignore non-substantial smoking) Ever/Never distinction (Ever=1) | integer | nullable | y.isin([0,1]) | | |
| 1 | Lifestyle | 3 | smoking_years | Smoking years (Can ignore non-substantial smoking) | numeric | nullable | (y>=0) & (y<=120) | | |
| 1 | Lifestyle | 4 | smoking_pack_years | Smoking pack (20) x years | numeric | nullable | (y>=0) & (y<=10000) | | |
| 0 | Clinical | 1 | clinical_dx_constipation | Does the participants have constipation? | string | nullable | ["Yes", "No"] | | |
| 0 | Clinical | 2 | clinical_dx_hyposmia | Does the participants have hyposmia? | string | nullable | ["Yes", "No"] | | |
| 0 | Clinical | 3 | clinical_dx_depression | Does the participants have depression? | string | nullable | ["Yes", "No"] | | |
| 0 | Clinical | 4 | clinical_dx_mild_cognitive_impairment | Does the participants have mild cognitive impairment? | string | nullable | ["Yes", "No"] | | |
| 0 | Clinical | 5 | clinical_dx_dementia | Does the participants have dementia? | string | nullable | ["Yes", "No"] | | |
| 0 | Clinical | 6 | clinical_dx_insomnia | Does the participants have insomnia? | string | nullable | ["Yes", "No"] | | |
| 0 | Clinical | 7 | clinical_dx_daytime_sleep | Does the participants have daytime sleepiness? | string | nullable | ["Yes", "No"] | | |
| 0 | Clinical | 8 | clinical_dx_dyskinesia | Does the participants have dyskinesia? | string | nullable | ["Yes", "No"] | | |
| 0 | Clinical | 8 | clinical_dx_waring_off | Does the participants have waring off? | string | nullable | ["Yes", "No"] | | |
| 0 | Medication | 1 | Age at current medication review | Age at evaluation of "Current Medication Review" | numeric | nullable | (y>=20) & (y<=120) | | |
| 0 | Medication | | medication_data_availability | Is the medication data available? Yes or no for the dataset | integer | nullable | ["Yes", "No"] | | |
| 0 | Medication | 2 | levodopa_mg_daily | Levodopa Dosage per day | numeric | nullable | (y>=0) & (y<=10000) | | |
| 0 | Medication | 3 | ledd_daily | Levodopa Equivalent Dosage (LEDD) per day (including all PD meds) | numeric | nullable | (y>=0) & (y<=10000) | | |
| 0 | Medication | 4 | levodopa_best_response_pct | Best ever patient reported response to levodopa (%: 0-100) | numeric | nullable | | | |
| 0 | Medication | 5 | levodopa_use | Usage of levodopa | string | nullable | ["Yes", "No"] | | |
| 0 | Medication | 6 | dopamine_agonist_use | Usage of dopamine agonist | string | nullable | ["Yes", "No"] | | |
| 0 | Medication | 7 | anticholinergics_use | Usage of anticholinergics | string | nullable | ["Yes", "No"] | | |
| 0 | Medication | 8 | brain_surgery | Usage of DBS/Brain surgery | string | nullable | ["Yes", "No"] | | |
| 0 | Medication | 9 | cholin_esterase_inhibitor_use | Usage of cholinesterase Inhibitors | string | nullable | ["Yes", "No"] | | |
| 0 | Medication | 10 | anti_depressant_use | Usage of anti-depressant drugs | string | nullable | ["Yes", "No"] | | |
| 0 | Medication | 11 | sleeping_pills_use | Usage of sleeping pills | string | nullable | ["Yes", "No"] | | |
| 0 | Medication | 12 | stomach_medicines_use | Usage of stomach medicines (including for nausea, GERD) | string | nullable | ["Yes", "No"] | | |
| 0 | Medication | 13 | laxatives_use | Usage of laxatives (not fibers) | string | nullable | ["Yes", "No"] | | |
| 0 | Medication | 14 | NSAIDs_use | Usage of NSAIDs | string | nullable | ["Yes", "No"] | | |
| 0 | Medication | 15 | morphines_use | Usage of morphines | string | nullable | ["Yes", "No"] | | |
| 0 | Medication | 16 | anti_psychotics_use | Usage of anti-psychotics | string | nullable | ["Yes", "No"] | | |
| 0 | Medication | 16 | insulin_use | Usage of insulin | string | nullable | ["Yes", "No"] | | |
| 0 | Medication | 16 | hormone_replacement_therapy | Usage of sex hormones replacement therapy | string | nullable | ["Yes", "No"] | | |
| 0 | Hoehn and | 36 | hoehn_and_yahr_stage | modified or original HY | numeric | nullable | (y>=0) & (y<=5) | code_upd2hy_hoehn_and_yahr_stage | convertible |
| 0 | Rankin | 1 | rankin_scale | modified or original Rankin Scale | numeric | nullable | (y>=0) & (y<=6) | | |
| 0 | MDS-UPDRS | 1 | mds_updrs_part_i_primary_info_source | MDS-UPDRS Part I Questions 1-6 Primary Source Of Information | string | nullable | | mds_updrs_part_i_primary_info_source | identical |
| 0 | MDS-UPDRS | 2 | code_upd2101_cognitive_impairment | 1.01 MDS-UPDRS - Cognitive Impairment (UPD2101) | integer | nullable | y.isin([0,1,2,3,4]) | code_upd2101_cognitive_impairment | identical |
| 0 | MDS-UPDRS | 3 | code_upd2102_hallucinations_and_psychosis | 1.02 MDS-UPDRS - Hallucinations and Psychosis (UPD2102) | integer | nullable | y.isin([0,1,2,3,4]) | code_upd2102_hallucinations_and_psychosis | identical |
| 0 | MDS-UPDRS | 4 | code_upd2103_depressed_mood | 1.03 MDS-UPDRS - Depressed Mood (UPD2103) | integer | nullable | y.isin([0,1,2,3,4]) | code_upd2103_depressed_mood | identical |
| 0 | MDS-UPDRS | 5 | code_upd2104_anxious_mood | 1.04 MDS-UPDRS - Anxious Mood (UPD2104) | integer | nullable | y.isin([0,1,2,3,4]) | code_upd2104_anxious_mood | identical |
| 0 | MDS-UPDRS | 6 | code_upd2105_apathy | 1.05 MDS-UPDRS - Apathy (UPD2105) | integer | nullable | y.isin([0,1,2,3,4]) | code_upd2105_apathy | identical |
| 0 | MDS-UPDRS | 7 | code_upd2106_dopamine_dysregulation_syndrome_features | 1.06 MDS-UPDRS - Features of Dopamine Dysregulation Syndrome (UPD2106) | integer | nullable | y.isin([0,1,2,3,4]) | code_upd2106_dopamine_dysregulation_syndrome_features | identical |
| 0 | MDS-UPDRS | 8 | mds_updrs_part_i_sub_score | MDS-UPDRS Part I Questions 1-6 Summary Sub-Score | integer | nullable | (y>=0) & (y<=24) | mds_updrs_part_i_sub_score | identical |
| 0 | MDS-UPDRS | 9 | mds_updrs_part_i_pat_quest_primary_info_source | MDS-UPDRS Part I Patient Questionnaire Primary Source Of Information | string | nullable | | mds_updrs_part_i_pat_quest_primary_info_source | identical |
| 0 | MDS-UPDRS | 10 | code_upd2107_pat_quest_sleep_problems | 1.07 MDS-UPDRS - Sleep Problems (UPD2107) | integer | nullable | y.isin([0,1,2,3,4]) | code_upd2107_pat_quest_sleep_problems | identical |
| 0 | MDS-UPDRS | 11 | code_upd2108_pat_quest_daytime_sleepiness | 1.08 MDS-UPDRS - Daytime Sleepiness (UPD2108) | integer | nullable | y.isin([0,1,2,3,4]) | code_upd2108_pat_quest_daytime_sleepiness | identical |
| 0 | MDS-UPDRS | 12 | code_upd2109_pat_quest_pain_and_other_sensations | 1.09 MDS-UPDRS - Pain And Other Sensations (UPD2109) | integer | nullable | y.isin([0,1,2,3,4]) | code_upd2109_pat_quest_pain_and_other_sensations | identical |
| 0 | MDS-UPDRS | 13 | code_upd2110_pat_quest_urinary_problems | 1.10 MDS-UPDRS - Urinary Problems (UPD2110) | integer | nullable | y.isin([0,1,2,3,4]) | code_upd2110_pat_quest_urinary_problems | identical |
| 0 | MDS-UPDRS | 14 | code_upd2111_pat_quest_constipation_problems | 1.11 MDS-UPDRS - Constipation Problems (UPD2111) | integer | nullable | y.isin([0,1,2,3,4]) | code_upd2111_pat_quest_constipation_problems | identical |
| 0 | MDS-UPDRS | 15 | code_upd2112_pat_quest_lightheadedness_on_standing | 1.12 MDS-UPDRS - Lightheadedness on Standing (UPD2112) | integer | nullable | y.isin([0,1,2,3,4]) | code_upd2112_pat_quest_lightheadedness_on_standing | identical |
| 0 | MDS-UPDRS | 16 | code_upd2113_pat_quest_fatigue | 1.13 MDS-UPDRS - Fatigue (UPD2113) | integer | nullable | y.isin([0,1,2,3,4]) | code_upd2113_pat_quest_fatigue | identical |

Global Parkinson's Genetics Program

### Supplementary Table 2. GP2 Core Data Dictionary (continued)

| Single measure | Modality | no. | Item | Description | ItemType | Required | Values | AMP_PD_item | Conversion | Comment |
|---|---|---|---|---|---|---|---|---|---|---|
| 0 | MDS-UPDRS | 17 | mds_updrs_part_i_pat_quest_sub_score | MDS-UPDRS Part I Patient Questionnaire Questions 7-13 Summary Sub-Score | integer | nullable | (y>=0) & (y<=28) | mds_updrs_part_i_pat_quest_sub_score | identical | |
| 0 | MDS-UPDRS | 18 | mds_updrs_part_i_summary_score | MDS-UPDRS Part I Summary Score | integer | nullable | (y>=0) & (y<=52) | mds_updrs_part_i_summary_score | identical | |
| 0 | MDS-UPDRS | 1 | mds_updrs_part_ii_primary_info_source | MDS-UPDRS Part II Primary Source Of Information | string | nullable | | mds_updrs_part_ii_primary_info_source | identical | |
| 0 | MDS-UPDRS | 2 | code_upd2201_speech | 2.01 MDS-UPDRS - Speech (UPD2201) | integer | nullable | y.isin([0,1,2,3,4]) | code_upd2201_speech | identical | |
| 0 | MDS-UPDRS | 3 | code_upd2202_saliva_and_drooling | 2.02 MDS-UPDRS - Saliva And Drooling (UPD2202) | integer | nullable | y.isin([0,1,2,3,4]) | code_upd2202_saliva_and_drooling | identical | |
| 0 | MDS-UPDRS | 4 | code_upd2203_chewing_and_swallowing | 2.03 MDS-UPDRS - Chewing And Swallowing (UPD2203) | integer | nullable | y.isin([0,1,2,3,4]) | code_upd2203_chewing_and_swallowing | identical | |
| 0 | MDS-UPDRS | 5 | code_upd2204_eating_tasks | 2.04 MDS-UPDRS - Eating Tasks (UPD2204) | integer | nullable | y.isin([0,1,2,3,4]) | code_upd2204_eating_tasks | identical | |
| 0 | MDS-UPDRS | 6 | code_upd2205_dressing | 2.05 MDS-UPDRS - Dressing (UPD2205) | integer | nullable | y.isin([0,1,2,3,4]) | code_upd2205_dressing | identical | |
| 0 | MDS-UPDRS | 7 | code_upd2206_hygiene | 2.06 MDS-UPDRS - Hygiene (UPD2206) | integer | nullable | y.isin([0,1,2,3,4]) | code_upd2206_hygiene | identical | |
| 0 | MDS-UPDRS | 8 | code_upd2207_handwriting | 2.07 MDS-UPDRS - Handwriting (UPD2207) | integer | nullable | y.isin([0,1,2,3,4]) | code_upd2207_handwriting | identical | |
| 0 | MDS-UPDRS | 9 | code_upd2208_doing_hobbies_and_other_activities | 2.08 MDS-UPDRS - Doing Hobbies And Other Activities (UPD2208) | integer | nullable | y.isin([0,1,2,3,4]) | code_upd2208_doing_hobbies_and_other_activities | identical | |
| 0 | MDS-UPDRS | 10 | code_upd2209_turning_in_bed | 2.09 MDS-UPDRS - Turning In Bed (UPD2209) | integer | nullable | y.isin([0,1,2,3,4]) | code_upd2209_turning_in_bed | identical | |
| 0 | MDS-UPDRS | 11 | code_upd2210_tremor | 2.10 MDS-UPDRS - Tremor (UPD2210) | integer | nullable | y.isin([0,1,2,3,4]) | code_upd2210_tremor | identical | |
| 0 | MDS-UPDRS | 12 | code_upd2211_get_out_of_bed_car_or_deep_chair | 2.11 MDS-UPDRS - Get Out Of Bed, Car, Or Deep Chair (UPD2211) | integer | nullable | y.isin([0,1,2,3,4]) | code_upd2211_get_out_of_bed_car_or_deep_chair | identical | |
| 0 | MDS-UPDRS | 13 | code_upd2212_walking_and_balance | 2.12 MDS-UPDRS - Walking And Balance (UPD2212) | integer | nullable | y.isin([0,1,2,3,4]) | code_upd2212_walking_and_balance | identical | |
| 0 | MDS-UPDRS | 14 | code_upd2213_freezing | 2.13 MDS-UPDRS - Freezing (UPD2213) | integer | nullable | y.isin([0,1,2,3,4]) | code_upd2213_freezing | identical | |
| 0 | MDS-UPDRS | 15 | mds_updrs_part_ii_summary_score | MDS-UPDRS Part II Summary Score | integer | nullable | (y>=0) & (y<=52) | mds_updrs_part_ii_summary_score | identical | |
| 0 | MDS-UPDRS | 37 | upd23a_medication_for_pd | 3a MDS-UPDRS - Medication For Parkinson's Disease (UPD23A) | string | nullable | ["No", "Yes"] | upd23a_medication_for_pd | identical | |
| 0 | MDS-UPDRS | 38 | upd23b_clinical_state_on_medication | 3b MDS-UPDRS - Clinical State On Medication (UPD23B) | string | nullable | ["ON", "OFF", "Unknown"] | upd23b_clinical_state_on_medication | identical | |
| 0 | MDS-UPDRS | 39 | upd23c_on_levodopa | 3c MDS-UPDRS - Is the patient on levodopa? | string | nullable | ["No", "Yes"] | | | |
| 0 | MDS-UPDRS | 40 | upd23c1_min_since_last_levodopa | 3c1 MDS-UPDRS - If patients on levodopa, minutes since last levodopa dose | numeric | nullable | (y>=0) | | | |
| 0 | MDS-UPDRS | 1 | code_upd2301_speech_problems | 3.01 MDS-UPDRS - Speech Problems (UPD2301) | integer | nullable | y.isin([0,1,2,3,4]) | code_upd2301_speech_problems | identical | |
| 0 | MDS-UPDRS | 2 | code_upd2302_facial_expression | 3.02 MDS-UPDRS - Facial Expression (UPD2302) | integer | nullable | y.isin([0,1,2,3,4]) | code_upd2302_facial_expression | identical | |
| 0 | MDS-UPDRS | 3 | code_upd2303a_rigidity_neck | 3.03a MDS-UPDRS - Rigidity Neck (UPD2303A) | integer | nullable | y.isin([0,1,2,3,4]) | code_upd2303a_rigidity_neck | identical | |
| 0 | MDS-UPDRS | 4 | code_upd2303b_rigidity_rt_upper_extremity | 3.03b MDS-UPDRS - Rigidity Right Upper Extremity (UPD2303B) | integer | nullable | y.isin([0,1,2,3,4]) | code_upd2303b_rigidity_rt_upper_extremity | identical | |
| 0 | MDS-UPDRS | 5 | code_upd2303c_rigidity_left_upper_extremity | 3.03c MDS-UPDRS - Rigidity Left Upper Extremity (UPD2303C) | integer | nullable | y.isin([0,1,2,3,4]) | code_upd2303c_rigidity_left_upper_extremity | identical | |
| 0 | MDS-UPDRS | 6 | code_upd2303d_rigidity_rt_lower_extremity | 3.03d MDS-UPDRS - Rigidity Right Lower Extremity (UPD2303D) | integer | nullable | y.isin([0,1,2,3,4]) | code_upd2303d_rigidity_rt_lower_extremity | identical | |
| 0 | MDS-UPDRS | 7 | code_upd2303e_rigidity_left_lower_extremity | 3.03e MDS-UPDRS - Rigidity Left Lower Extremity (UPD2303E) | integer | nullable | y.isin([0,1,2,3,4]) | code_upd2303e_rigidity_left_lower_extremity | identical | |
| 0 | MDS-UPDRS | 8 | code_upd2304a_right_finger_tapping | 3.04a MDS-UPDRS - Right Finger Tapping (UPD2304A) | integer | nullable | y.isin([0,1,2,3,4]) | code_upd2304a_right_finger_tapping | identical | |
| 0 | MDS-UPDRS | 9 | code_upd2304b_left_finger_tapping | 3.04b MDS-UPDRS - Left Finger Tapping (UPD2304B) | integer | nullable | y.isin([0,1,2,3,4]) | code_upd2304b_left_finger_tapping | identical | |
| 0 | MDS-UPDRS | 10 | code_upd2305a_right_hand_movements | 3.05a MDS-UPDRS - Right Hand Movements (UPD2305A) | integer | nullable | y.isin([0,1,2,3,4]) | code_upd2305a_right_hand_movements | identical | |
| 0 | MDS-UPDRS | 11 | code_upd2305b_left_hand_movements | 3.05b MDS-UPDRS - Left Hand Movements (UPD2305B) | integer | nullable | y.isin([0,1,2,3,4]) | code_upd2305b_left_hand_movements | identical | |
| 0 | MDS-UPDRS | 12 | code_upd2306a_pron_sup_movement_right_hand | 3.06a MDS-UPDRS - Pronanation-Supination Movement Of Right Hand (UPD2306A) | integer | nullable | y.isin([0,1,2,3,4]) | code_upd2306a_pron_sup_movement_right_hand | identical | |
| 0 | MDS-UPDRS | 13 | code_upd2306b_pron_sup_movement_left_hand | 3.06b MDS-UPDRS - Pronanation-Supination Movement Of Left Hand (UPD2306B) | integer | nullable | y.isin([0,1,2,3,4]) | code_upd2306b_pron_sup_movement_left_hand | identical | |
| 0 | MDS-UPDRS | 14 | code_upd2307a_right_toe_tapping | 3.07a MDS-UPDRS - Right Toe Tapping (UPD2307A) | integer | nullable | y.isin([0,1,2,3,4]) | code_upd2307a_right_toe_tapping | identical | |
| 0 | MDS-UPDRS | 15 | code_upd2307b_left_toe_tapping | 3.07b MDS-UPDRS - Left Toe Tapping (UPD2307B) | integer | nullable | y.isin([0,1,2,3,4]) | code_upd2307b_left_toe_tapping | identical | |
| 0 | MDS-UPDRS | 16 | code_upd2308a_right_leg_agility | 3.08a MDS-UPDRS - Right Leg Agility (UPD2308A) | integer | nullable | y.isin([0,1,2,3,4]) | code_upd2308a_right_leg_agility | identical | |
| 0 | MDS-UPDRS | 17 | code_upd2308b_left_leg_agility | 3.08b MDS-UPDRS - Left Leg Agility (UPD2308B) | integer | nullable | y.isin([0,1,2,3,4]) | code_upd2308b_left_leg_agility | identical | |
| 0 | MDS-UPDRS | 18 | code_upd2309_arising_from_chair | 3.09 MDS-UPDRS - Arising from Chair (UPD2309) | integer | nullable | y.isin([0,1,2,3,4]) | code_upd2309_arising_from_chair | identical | |
| 0 | MDS-UPDRS | 19 | code_upd2310_gait | 3.10 MDS-UPDRS - Gait (UPD2310) | integer | nullable | y.isin([0,1,2,3,4]) | code_upd2310_gait | identical | |
| 0 | MDS-UPDRS | 20 | code_upd2311_freezing_of_gait | 3.11 MDS-UPDRS - Freezing Of Gait (UPD2311) | integer | nullable | y.isin([0,1,2,3,4]) | code_upd2311_freezing_of_gait | identical | |
| 0 | MDS-UPDRS | 21 | code_upd2312_postural_stability | 3.12 MDS-UPDRS - Postural Stability (UPD2312) | integer | nullable | y.isin([0,1,2,3,4]) | code_upd2312_postural_stability | identical | |
| 0 | MDS-UPDRS | 22 | code_upd2313_posture | 3.13 MDS-UPDRS - Posture (UPD2313) | integer | nullable | y.isin([0,1,2,3,4]) | code_upd2313_posture | identical | |
| 0 | MDS-UPDRS | 23 | code_upd2314_body_bradykinesia | 3.14 MDS-UPDRS - Body Bradykinesia (UPD2314) | integer | nullable | y.isin([0,1,2,3,4]) | code_upd2314_body_bradykinesia | identical | |
| 0 | MDS-UPDRS | 24 | code_upd2315a_postural_tremor_of_right_hand | 3.15a MDS-UPDRS - Postural Tremor Of Right Hand (UPD2315A) | integer | nullable | y.isin([0,1,2,3,4]) | code_upd2315a_postural_tremor_of_right_hand | identical | |
| 0 | MDS-UPDRS | 25 | code_upd2315b_postural_tremor_of_left_hand | 3.15b MDS-UPDRS - Postural Tremor Of Left Hand (UPD2315B) | integer | nullable | y.isin([0,1,2,3,4]) | code_upd2315b_postural_tremor_of_left_hand | identical | |
| 0 | MDS-UPDRS | 26 | code_upd2316a_kinetic_tremor_of_right_hand | 3.16a MDS-UPDRS - Kinetic Tremor Of Right Hand (UPD2316A) | integer | nullable | y.isin([0,1,2,3,4]) | code_upd2316a_kinetic_tremor_of_right_hand | identical | |
| 0 | MDS-UPDRS | 27 | code_upd2316b_kinetic_tremor_of_left_hand | 3.16b MDS-UPDRS - Kinetic Tremor Of Left Hand (UPD2316B) | integer | nullable | y.isin([0,1,2,3,4]) | code_upd2316b_kinetic_tremor_of_left_hand | identical | |
| 0 | MDS-UPDRS | 28 | code_upd2317a_rest_tremor_amplitude_right_upper_extremity | 3.17a MDS-UPDRS - Rest Tremor Amplitude Right Upper Extremity (UPD2317A) | integer | nullable | y.isin([0,1,2,3,4]) | code_upd2317a_rest_tremor_amplitude_right_upper_extremity | identical | |
| 0 | MDS-UPDRS | 29 | code_upd2317b_rest_tremor_amplitude_left_upper_extremity | 3.17b MDS-UPDRS - Rest Tremor Amplitude Left Upper Extremity (UPD2317B) | integer | nullable | y.isin([0,1,2,3,4]) | code_upd2317b_rest_tremor_amplitude_left_upper_extremity | identical | |
| 0 | MDS-UPDRS | 30 | code_upd2317c_rest_tremor_amplitude_right_lower_extremity | 3.17c MDS-UPDRS - Rest Tremor Amplitude Right Lower Extremity (UPD2317C) | integer | nullable | y.isin([0,1,2,3,4]) | code_upd2317c_rest_tremor_amplitude_right_lower_extremity | identical | |
| 0 | MDS-UPDRS | 31 | code_upd2317d_rest_tremor_amplitude_left_lower_extremity | 3.17d MDS-UPDRS - Rest Tremor Amplitude Left Lower Extremity (UPD2317D) | integer | nullable | y.isin([0,1,2,3,4]) | code_upd2317d_rest_tremor_amplitude_left_lower_extremity | identical | |
| 0 | MDS-UPDRS | 32 | code_upd2317e_rest_tremor_amplitude_lip_or_jaw | 3.17e MDS-UPDRS - Rest Tremor Amplitude Lip Or Jaw (UPD2317E) | integer | nullable | y.isin([0,1,2,3,4]) | code_upd2317e_rest_tremor_amplitude_lip_or_jaw | identical | |
| 0 | MDS-UPDRS | 33 | code_upd2318_consistency_of_rest_tremor | 3.18 MDS-UPDRS - Consistency Of Rest Tremor (UPD2318) | integer | nullable | y.isin([0,1,2,3,4]) | code_upd2318_consistency_of_rest_tremor | identical | |
| 0 | MDS-UPDRS | 34 | upd2da_dyskinesias_during_exam | 3.19 MDS-UPDRS - Dyskinesias During Exam (UPD2DA) | string | nullable | ["No", "Yes"] | upd2da_dyskinesias_during_exam | identical | |
| 0 | MDS-UPDRS | 35 | upd2db_movements_interfere_with_ratings | 3.20 MDS-UPDRS - Movements Interfere with Ratings (UPD2DB) | string | nullable | ["No", "Yes"] | upd2db_movements_interfere_with_ratings | identical | |
| 0 | MDS-UPDRS | 41 | mds_updrs_part_iii_summary_score | MDS-UPDRS Part III Summary Score | integer | nullable | (y>=0) & (y<=132) | mds_updrs_part_iii_summary_score | identical | |
| 0 | MDS-UPDRS | 1 | code_upd2401_time_spent_with_dyskinesias | 4.01 MDS-UPDRS - Time Spent With Dyskinesias (UPD2401) | integer | nullable | y.isin([0,1,2,3,4]) | code_upd2401_time_spent_with_dyskinesias | identical | |
| 0 | MDS-UPDRS | 2 | code_upd2402_functional_impact_of_dyskinesias | 4.02 MDS-UPDRS - Functional Impact Of Dyskinesias (UPD2402) | integer | nullable | y.isin([0,1,2,3,4]) | code_upd2402_functional_impact_of_dyskinesias | identical | |
| 0 | MDS-UPDRS | 3 | code_upd2403_time_spent_in_the_off_state | 4.03 MDS-UPDRS - Time Spent In The Off State (UPD2403) | integer | nullable | y.isin([0,1,2,3,4]) | code_upd2403_time_spent_in_the_off_state | identical | |
| 0 | MDS-UPDRS | 4 | code_upd2404_functional_impact_of_fluctuations | 4.04 MDS-UPDRS - Functional Impact Of Fluctuations (UPD2404) | integer | nullable | y.isin([0,1,2,3,4]) | code_upd2404_functional_impact_of_fluctuations | identical | |
| 0 | MDS-UPDRS | 5 | code_upd2405_complexity_of_motor_fluctuations | 4.05 MDS-UPDRS - Complexity Of Motor Fluctuations (UPD2405) | integer | nullable | y.isin([0,1,2,3,4]) | code_upd2405_complexity_of_motor_fluctuations | identical | |
| 0 | MDS-UPDRS | 6 | code_upd2406_painful_off_state_dystonia | 4.06 MDS-UPDRS - Painful Off-State Dystonia (UPD2406) | integer | nullable | y.isin([0,1,2,3,4]) | code_upd2406_painful_off_state_dystonia | identical | |
| 0 | MDS-UPDRS | 7 | mds_updrs_part_iv_summary_score | MDS-UPDRS Part IV Summary Score | integer | nullable | (y>=0) & (y<=24) | mds_updrs_part_iv_summary_score | identical | |
| 0 | MDS-UPDRS | 8 | upd2406_1_total_hours_off | 4.06.1 MDS-UPDRS - Total Hours OFF | numeric | nullable | (y>=0) & (y<=1440) | | | |
| 0 | MDS-UPDRS | 9 | upd2406_2_off_hours_wo_dystonia | 4.06.2 MDS-UPDRS - Total OFF Hours with Dystonia | numeric | nullable | (y>=0) & (y<=1440) | | | |
| 0 | MDS-UPDRS | 10 | upd2406_3_pct_off_dystonia | 4.06.3 MDS-UPDRS - % OFF Dystonia = ((2/1)*100) | numeric | nullable | (y>=0) & (y<=100) | | | |
| 0 | MoCA | 1 | moca01_alternating_trail_making | MOCA: 01. Alternating Trail Making | integer | nullable | y.isin([0,1]) | moca01_alternating_trail_making | identical | |
| 0 | MoCA | 2 | moca02_visuoconstr_skills_cube | MOCA: 02. Visuoconstructional Skills - Copy Cube | integer | nullable | y.isin([0,1]) | moca02_visuoconstr_skills_cube | identical | |
| 0 | MoCA | 3 | moca03_visuoconstr_skills_clock_cont | MOCA: 03. Visuoconstructional Skills - Draw Clock Contour | integer | nullable | y.isin([0,1]) | moca03_visuoconstr_skills_clock_cont | identical | |
| 0 | MoCA | 4 | moca04_visuoconstr_skills_clock_num | MOCA: 04. Visuoconstructional Skills - Draw Clock Numbers | integer | nullable | y.isin([0,1]) | moca04_visuoconstr_skills_clock_num | identical | |
| 0 | MoCA | 5 | moca05_visuoconstr_skills_clock_hands | MOCA: 05. Visuoconstructional Skills - Draw Clock Hands | integer | nullable | y.isin([0,1]) | moca05_visuoconstr_skills_clock_hands | identical | |
| 0 | MoCA | 5 | moca05_visuoconstr_skills_clock | MOCA: Draw Clock Total | integer | nullable | y.isin([0,1,2,3]) | | | |
| 0 | MoCA | 6 | moca_visuospatial_executive_subscore | MOCA: Visuospatial And Executive Subscore | integer | nullable | y.isin([0,1,2,3,4,5]) | moca_visuospatial_executive_subscore | identical | |

### Supplementary Table 2. GP2 Core Data Dictionary (continued)

| Single measure | Modality | no. | Item | Description | ItemType | Required | Values | AMP_PD_item | Conversion | Comment |
|---|---|---|---|---|---|---|---|---|---|---|
| 0 | MoCA | 7 | moca06_naming_lion | MOCA: 06. Naming - Lion | integer | nullable | y.isin([0,1]) | moca06_naming_lion | identical | |
| 0 | MoCA | 8 | moca07_naming_rhino | MOCA: 07. Naming - Rhino | integer | nullable | y.isin([0,1]) | moca07_naming_rhino | identical | |
| 0 | MoCA | 9 | moca08_naming_camel | MOCA: 08. Naming - Camel | integer | nullable | y.isin([0,1]) | moca08_naming_camel | identical | |
| 0 | MoCA | 10 | moca_naming_subscore | MOCA: Naming Subscore | integer | nullable | y.isin([0,1,2,3]) | moca_naming_subscore | identical | |
| 0 | MoCA | 11 | moca09_attention_forward_digit_span | MOCA: 09. Attention - Forward Digit Span | integer | nullable | y.isin([0,1]) | moca09_attention_forward_digit_span | identical | |
| 0 | MoCA | 12 | moca10_attention_backward_digit_span | MOCA: 10. Attention - Backward Digit Span | integer | nullable | y.isin([0,1]) | moca10_attention_backward_digit_span | identical | |
| 0 | MoCA | 13 | moca_attention_digits_subscore | MOCA: Attention Forward-Backward Repeat Lists Of Digits Subscore | integer | nullable | y.isin([0,1,2]) | moca_attention_digits_subscore | identical | |
| 0 | MoCA | 14 | moca11_attention_vigilance | MOCA: 11. Attention - Vigilance | integer | nullable | y.isin([0,1]) | moca11_attention_vigilance | identical | |
| 0 | MoCA | 15 | moca12_attention_serial_7s | MOCA: 12. Attention - Serial 7s | integer | nullable | y.isin([0,1,2,3]) | moca12_attention_serial_7s | identical | |
| 0 | MoCA | 16 | moca_attention_subscore | MOCA: Attention domain subscore | integer | nullable | y.isin([0,1,2,3,4,5,6]) | | | |
| 0 | MoCA | 17 | moca13_sentence_repetition | MOCA: 13. Sentence Repetition | integer | nullable | y.isin([0,1,2]) | | | |
| 0 | MoCA | 18 | moca13_sentence_repetition_1 | MOCA: 13. Sentence Repetition 1 | integer | nullable | y.isin([0,1]) | moca13_sentence_repetition | partial | |
| 0 | MoCA | 19 | moca13_sentence_repetition_2 | MOCA: 13. Sentence Repetition 2 | integer | nullable | y.isin([0,1]) | moca13_sentence_repetition | partial | |
| 0 | MoCA | 20 | moca14_verbal_fluency_number_of_words | MOCA: 14. Verbal Fluency - Number Of Words | integer | nullable | (y>=0) & (y<=100) | moca14_verbal_fluency_number_of_words | identical | |
| 0 | MoCA | 21 | moca15_verbal_fluency | MOCA: 15. Verbal Fluency | integer | nullable | y.isin([0,1]) | moca15_verbal_fluency | identical | |
| 0 | MoCA | 22 | moca_language_subscore | MOCA: Language Subscore | integer | nullable | y.isin([0,1,2,3]) | moca_language_subscore | identical | |
| 0 | MoCA | 23 | moca16_abstraction_1 | MOCA: 16. Abstraction 1 | integer | nullable | y.isin([0,1]) | moca16_abstraction | partial | |
| 0 | MoCA | 24 | moca16_abstraction_2 | MOCA: 16. Abstraction 2 | integer | nullable | y.isin([0,1]) | moca16_abstraction | partial | |
| 0 | MoCA | 25 | moca_abstraction_subscore | moca16_abstraction_1 + moca16_abstraction_2 | integer | nullable | y.isin([0,1,2]) | moca_abstraction_subscore | identical | |
| 0 | MoCA | 26 | moca17_delayed_recall_face | MOCA: 17. Delayed Recall - Face | integer | nullable | y.isin([0,1]) | moca17_delayed_recall_face | identical | |
| 0 | MoCA | 27 | moca18_delayed_recall_velvet | MOCA: 18. Delayed Recall - Velvet | integer | nullable | y.isin([0,1]) | moca18_delayed_recall_velvet | identical | |
| 0 | MoCA | 28 | moca19_delayed_recall_church | MOCA: 19. Delayed Recall - Church | integer | nullable | y.isin([0,1]) | moca19_delayed_recall_church | identical | |
| 0 | MoCA | 29 | moca20_delayed_recall_daisy | MOCA: 20. Delayed Recall - Daisy | integer | nullable | y.isin([0,1]) | moca20_delayed_recall_daisy | identical | |
| 0 | MoCA | 30 | moca21_delayed_recall_red | MOCA: 21. Delayed Recall - Red | integer | nullable | y.isin([0,1]) | moca21_delayed_recall_red | identical | |
| 0 | MoCA | 31 | moca_delayed_recall_subscore_optnl_cat_cue | MOCA: Delayed Recall Subscore Optional Category Cue | integer | nullable | y.isin([0,1,2,3,4,5]) | moca_delayed_recall_subscore_optnl_cat_cue | identical | |
| 0 | MoCA | 32 | moca_delayed_recall_subscore_optnl_mult_choice | MOCA: Delayed Recall Subscore Optional Multiple Choice Cue | integer | nullable | y.isin([0,1,2,3,4,5]) | moca_delayed_recall_subscore_optnl_mult_choice | identical | |
| 0 | MoCA | 33 | moca_delayed_recall_subscore | MOCA: Delayed Recall Subscore Uncued | integer | nullable | y.isin([0,1,2,3,4,5]) | moca_delayed_recall_subscore | identical | |
| 0 | MoCA | 34 | moca22_orientation_date_score | MOCA: 22. Orientation - Date Score | integer | nullable | y.isin([0,1]) | moca22_orientation_date_score | identical | |
| 0 | MoCA | 35 | moca23_orientation_month_score | MOCA: 23. Orientation - Month Score | integer | nullable | y.isin([0,1]) | moca23_orientation_month_score | identical | |
| 0 | MoCA | 36 | moca24_orientation_year_score | MOCA: 24. Orientation - Year Score | integer | nullable | y.isin([0,1]) | moca24_orientation_year_score | identical | |
| 0 | MoCA | 37 | moca25_orientation_day_score | MOCA: 25. Orientation - Day Score | integer | nullable | y.isin([0,1]) | moca25_orientation_day_score | identical | |
| 0 | MoCA | 38 | moca26_orientation_place_score | MOCA: 26. Orientation - Place Score | integer | nullable | y.isin([0,1]) | moca26_orientation_place_score | identical | |
| 0 | MoCA | 39 | moca27_orientation_city_score | MOCA: 27. Orientation - City Score | integer | nullable | y.isin([0,1]) | moca27_orientation_city_score | identical | |
| 0 | MoCA | 40 | moca_orientation_subscore | MOCA: Orientation Subscore | integer | nullable | y.isin([0,1,2,3,4,5,6]) | moca_orientation_subscore | identical | |
| 0 | MoCA | 41 | moca_total_score | MOCA Total Score | integer | nullable | (y>=0) & (y<=30) | moca_total_score | identical | |
| 0 | MoCA | 42 | moca_total_score_adjusted | MoCA Total Score adjusted for educationo | integer | nullable | (y>=0) & (y<=30) | | | |
| 0 | SDM | 1 | sdm_score | Symbol Digit Modalities Test | integer | nullable | (y>=0) & (y<=110) | | | |
| 0 | SCOPA-AUT | 1 | scopa_aut01_swallowing | SCOPA-AUT item 1 | string | nullable | ['never', 'sometimes', 'regularly', 'often'] | | | |
| 0 | SCOPA-AUT | 2 | scopa_aut02_dribbling | SCOPA-AUT item 2 | string | nullable | ['never', 'sometimes', 'regularly', 'often'] | | | |
| 0 | SCOPA-AUT | 3 | scopa_aut03_food_stuck | SCOPA-AUT item 3 | string | nullable | ['never', 'sometimes', 'regularly', 'often'] | | | |
| 0 | SCOPA-AUT | 4 | scopa_aut04_stomach_full_quickly | SCOPA-AUT item 4 | string | nullable | ['never', 'sometimes', 'regularly', 'often'] | | | |
| 0 | SCOPA-AUT | 5 | scopa_aut05_bowel_constipation | SCOPA-AUT item 5 | string | nullable | ['never', 'sometimes', 'regularly', 'often'] | | | |
| 0 | SCOPA-AUT | 6 | scopa_aut06_bowel_strain_hard | SCOPA-AUT item 6 | string | nullable | ['never', 'sometimes', 'regularly', 'often'] | | | |
| 0 | SCOPA-AUT | 7 | scopa_aut07_bowel_involuntary_loss | SCOPA-AUT item 7 | string | nullable | ['never', 'sometimes', 'regularly', 'often'] | | | |
| 0 | SCOPA-AUT | 8 | scopa_aut08_urine_hard_to_retain | SCOPA-AUT item 8 | string | nullable | ['never', 'sometimes', 'regularly', 'often', 'use | | | |
| 0 | SCOPA-AUT | 9 | scopa_aut09_urine_involuntary_loss | SCOPA-AUT item 9 | string | nullable | ['never', 'sometimes', 'regularly', 'often', 'use | | | |
| 0 | SCOPA-AUT | 10 | scopa_aut10_urine_residual | SCOPA-AUT item 10 | string | nullable | ['never', 'sometimes', 'regularly', 'often', 'use | | | |
| 0 | SCOPA-AUT | 11 | scopa_aut11_urine_stream_weak | SCOPA-AUT item 11 | string | nullable | ['never', 'sometimes', 'regularly', 'often', 'use | | | |
| 0 | SCOPA-AUT | 12 | scopa_aut12_urine_pass_less_2hr | SCOPA-AUT item 12 | string | nullable | ['never', 'sometimes', 'regularly', 'often', 'use | | | |
| 0 | SCOPA-AUT | 13 | scopa_aut13_urine_night | SCOPA-AUT item 13 | string | nullable | ['never', 'sometimes', 'regularly', 'often', 'use | | | |
| 0 | SCOPA-AUT | 14 | scopa_aut14_light_head_stand_up | SCOPA-AUT item 14 | string | nullable | ['never', 'sometimes', 'regularly', 'often'] | | | |
| 0 | SCOPA-AUT | 15 | scopa_aut15_light_head_standing | SCOPA-AUT item 15 | string | nullable | ['never', 'sometimes', 'regularly', 'often'] | | | |
| 0 | SCOPA-AUT | 16 | scopa_aut16_faint | SCOPA-AUT item 16 | string | nullable | ['never', 'sometimes', 'regularly', 'often'] | | | |
| 0 | SCOPA-AUT | 17 | scopa_aut17_perspiration_day | SCOPA-AUT item 17 | string | nullable | ['never', 'sometimes', 'regularly', 'often'] | | | |
| 0 | SCOPA-AUT | 18 | scopa_aut18_perspiration_night | SCOPA-AUT item 18 | string | nullable | ['never', 'sometimes', 'regularly', 'often'] | | | |
| 0 | SCOPA-AUT | 19 | scopa_aut19_too_bright | SCOPA-AUT item 19 | string | nullable | ['never', 'sometimes', 'regularly', 'often'] | | | |
| 0 | SCOPA-AUT | 20 | scopa_aut20_too_cold | SCOPA-AUT item 20 | string | nullable | ['never', 'sometimes', 'regularly', 'often'] | | | |
| 0 | SCOPA-AUT | 21 | scopa_aut21_too_hot | SCOPA-AUT item 21 | string | nullable | ['never', 'sometimes', 'regularly', 'often'] | | | |
| 0 | SCOPA-AUT | 22 | scopa_aut22_sex_impotent | SCOPA-AUT item 22 | string | nullable | ['never', 'sometimes', 'regularly', 'often', 'not | | | |
| 0 | SCOPA-AUT | 23 | scopa_aut23_sex_unable_ejaculate | SCOPA-AUT item 23 | string | nullable | ['never', 'sometimes', 'regularly', 'often', 'not | | | |
| 0 | SCOPA-AUT | 24 | scopa_aut23a_sex_medication | SCOPA-AUT item 23a | string | nullable | ['no', 'yes', 'unknown'] | | | |
| 0 | SCOPA-AUT | 25 | scopa_aut23at_sex_medication_text | SCOPA-AUT item 23a ED Med | string | nullable | | | | |
| 0 | SCOPA-AUT | 26 | scopa_aut24_sex_dry | SCOPA-AUT item 24 | string | nullable | ['never', 'sometimes', 'regularly', 'often', 'not | | | |
| 0 | SCOPA-AUT | 27 | scopa_aut25_sex_difficulty_orgasm | SCOPA-AUT item 25 | string | nullable | ['never', 'sometimes', 'regularly', 'often', 'not | | | |
| 0 | SCOPA-AUT | 28 | scopa_aut26a_constipation_med | SCOPA-AUT item 26a | string | nullable | ['no', 'yes', 'unknown'] | | | |
| 0 | SCOPA-AUT | 29 | scopa_aut26at_constipation_med_text | SCOPA-AUT item 26a med | string | nullable | | | | |
| 0 | SCOPA-AUT | 30 | scopa_aut26b_urine_med | SCOPA-AUT item 26b | string | nullable | ['no', 'yes', 'unknown'] | | | |
| 0 | SCOPA-AUT | 31 | scopa_aut26bt_urine_med_text | SCOPA-AUT item 26b med | string | nullable | | | | |
| 0 | SCOPA-AUT | 32 | scopa_aut26c_blood_pressure_med | SCOPA-AUT item 26c | string | nullable | ['no', 'yes', 'unknown'] | | | |
| 0 | SCOPA-AUT | 33 | scopa_aut26ct_blood_pressure_med_text | SCOPA-AUT item 26c med | string | nullable | | | | |
| 0 | SCOPA-AUT | 34 | scopa_aut26d_other_med | SCOPA-AUT item 26d | string | nullable | ['no', 'yes', 'unknown'] | | | |
| 0 | SCOPA-AUT | 35 | scopa_aut26dt_other_med_text | SCOPA-AUT item 26d med | string | nullable | | | | |
| 0 | SCOPA- | 1 | scopa_cog1_verbal_recall | SCOPA-COG item 1 | integer | nullable | y.isin([0,1,2,3,4,5]) | | | |
| 0 | SCOPA- | 2 | scopa_cog2_digit_span_backward | SCOPA-COG item 2 | integer | nullable | y.isin([0,1,2,3,4,5,6,7]) | | | |
| 0 | SCOPA- | 3 | scopa_cog3_cube | SCOPA-COG item 3 | integer | nullable | y.isin([0,1,2,3,4,5]) | | | |
| 0 | SCOPA- | 4 | scopa_cog4_count_backwards | SCOPA-COG item 4 | integer | nullable | y.isin([0,1,2]) | | | |
| 0 | SCOPA- | 5 | scopa_cog5_months_backward | SCOPA-COG item 5 | integer | nullable | y.isin([0,1,2]) | | | |
| 0 | SCOPA- | 6 | scopa_cog6_fist_edge_palm | SCOPA-COG item 6 | integer | nullable | y.isin([0,1,2,3]) | | | |
| 0 | SCOPA- | 7 | scopa_cog7_semantic_fluency | SCOPA-COG item 7 | integer | nullable | y.isin([0,1,2,3,4,5,6]) | | | |
| 0 | SCOPA- | 8 | scopa_cog8_dice | SCOPA-COG item 8 | integer | nullable | y.isin([0,1,2,3]) | | | |
| 0 | SCOPA- | 9 | scopa_cog9_assembling_figures | SCOPA-COG item 9 | integer | nullable | y.isin([0,1,2,3,4,5]) | | | |
| 0 | SCOPA- | 10 | scopa_cog10_delayed_recall | SCOPA-COG item 10 | integer | nullable | y.isin([0,1,2,3,4,5]) | | | |
| 0 | SCOPA- | 15 | scopa_cog_total_score | SCOPA-COG Total Score | integer | nullable | (y>=0) & (y<=43) | | | |

Global Parkinson's Genetics Program

#### Supplementary Table 2. GP2 Core Data Dictionary (continued)

| Single measure | Modality | no. | Item | Description | ItemType | Required | Values | AMP_PD_item | Conversion | Comment |
|---|---|---|---|---|---|---|---|---|---|---|
| 0 | RBD Screening Questionnaire | 1 | rbd_info_source | RBD Source Of Information | string | nullable | | code_rbd_info_source | identical | |
| 0 | RBD Screening Questionnaire | 2 | code_rbd01_vivid_dreams | RBD 01. Vivid Dreams {1:yes, 0:no, null:unknown} | integer | nullable | y.isin([0,1]) | code_rbd01_vivid_dreams | identical | |
| 0 | RBD Screening Questionnaire | 3 | code_rbd02_aggressive_or_action_packed_dreams | RBD 02. Aggressive Or Action-Packed Dreams {1:yes, 0:no, null:unknown} | integer | nullable | y.isin([0,1]) | code_rbd02_aggressive_or_action_packed_dreams | identical | |
| 0 | RBD Screening Questionnaire | 4 | code_rbd03_nocturnal_behaviour | RBD 03. Nocturnal Behaviour {1:yes, 0:no, null:unknown} | integer | nullable | y.isin([0,1]) | code_rbd03_nocturnal_behaviour | identical | |
| 0 | RBD Screening Questionnaire | 5 | code_rbd04_move_arms_legs_during_sleep | RBD 04. Move Arms and Legs During Sleep {1:yes, 0:no, null:unknown} | integer | nullable | y.isin([0,1]) | code_rbd04_move_arms_legs_during_sleep | identical | |
| 0 | RBD Screening Questionnaire | 6 | code_rbd05_hurt_bed_partner | RBD 05. Hurt Bed Partner {1:yes, 0:no, null:unknown} | integer | nullable | y.isin([0,1]) | code_rbd05_hurt_bed_partner | identical | |
| 0 | RBD Screening Questionnaire | 7 | code_rbd06_1_speaking_in_sleep | RBD 06.1 Speaking In Sleep {1:yes, 0:no, null:unknown} | integer | nullable | y.isin([0,1]) | code_rbd06_1_speaking_in_sleep | identical | |
| 0 | RBD Screening Questionnaire | 8 | code_rbd06_2_sudden_limb_movements | RBD 06.2 Sudden Limb Movements {1:yes, 0:no, null:unknown} | integer | nullable | y.isin([0,1]) | code_rbd06_2_sudden_limb_movements | identical | |
| 0 | RBD Screening Questionnaire | 9 | code_rbd06_3_complex_movements | RBD 06.3 Complex Movements {1:yes, 0:no, null:unknown} | integer | nullable | y.isin([0,1]) | code_rbd06_3_complex_movements | identical | |
| 0 | RBD Screening Questionnaire | 10 | code_rbd06_4_things_fell_down | RBD 06.4 Things Fell Down {1:yes, 0:no, null:unknown} | integer | nullable | y.isin([0,1]) | code_rbd06_4_things_fell_down | identical | |
| 0 | RBD Screening Questionnaire | 11 | code_rbd07_my_movements_awake_me | RBD 07. My Movements Awake Me {1:yes, 0:no, null:unknown} | integer | nullable | y.isin([0,1]) | code_rbd07_my_movements_awake_me | identical | |
| 0 | RBD Screening Questionnaire | 12 | code_rbd08_remember_dreams | RBD 08. Remember Dreams {1:yes, 0:no, null:unknown} | integer | nullable | y.isin([0,1]) | code_rbd08_remember_dreams | identical | |
| 0 | RBD Screening Questionnaire | 13 | code_rbd09_sleep_is_disturbed | RBD 09. Sleep Is Disturbed {1:yes, 0:no, null:unknown} | integer | nullable | y.isin([0,1]) | code_rbd09_sleep_is_disturbed | identical | |
| 0 | RBD Screening Questionnaire | 14 | code_rbd10a_stroke | RBD 10a Stroke {1:yes, 0:no, null:unknown} | integer | nullable | y.isin([0,1]) | code_rbd10a_stroke | identical | |
| 0 | RBD Screening Questionnaire | 15 | code_rbd10b_head_trauma | RBD 10b Head Trauma {1:yes, 0:no, null:unknown} | integer | nullable | y.isin([0,1]) | code_rbd10b_head_trauma | identical | |
| 0 | RBD Screening Questionnaire | 16 | code_rbd10c_parkinsonism | RBD 10c Parkinsonism {1:yes, 0:no, null:unknown} | integer | nullable | y.isin([0,1]) | code_rbd10c_parkinsonism | identical | |
| 0 | RBD Screening Questionnaire | 17 | code_rbd10d_rls | RBD 10d RLS {1:yes, 0:no, null:unknown} | integer | nullable | y.isin([0,1]) | code_rbd10d_rls | identical | |
| 0 | RBD Screening Questionnaire | 18 | code_rbd10e_narcolepsy | RBD 10e Narcolepsy {1:yes, 0:no, null:unknown} | integer | nullable | y.isin([0,1]) | code_rbd10e_narcolepsy | identical | |
| 0 | RBD Screening Questionnaire | 19 | code_rbd10f_depression | RBD 10f Depression {1:yes, 0:no, null:unknown} | integer | nullable | y.isin([0,1]) | code_rbd10f_depression | identical | |
| 0 | RBD Screening Questionnaire | 20 | code_rbd10g_epilepsy | RBD 10g Epilepsy {1:yes, 0:no, null:unknown} | integer | nullable | y.isin([0,1]) | code_rbd10g_epilepsy | identical | |
| 0 | RBD Screening Questionnaire | 21 | code_rbd10h_brain_inflammatory_disease | RBD 10h Inflammatory Disease Of The Brain {1:yes, 0:no, null:unknown} | integer | nullable | y.isin([0,1]) | code_rbd10h_brain_inflammatory_disease | identical | |
| 0 | RBD Screening Questionnaire | 22 | code_rbd10i_other | RBD 10i Other {1:yes, 0:no, null:unknown} | integer | nullable | y.isin([0,1]) | code_rbd10i_other | identical | |
| 0 | RBD Screening Questionnaire | 23 | code_rbd10i_other_comment | RBD 10i Other (comment) | string | nullable | | | | |
| 0 | RBD Screening Questionnaire | 24 | code_rbd10_nervous_system_disease | RBD 10. Nervous system Disease {1:yes, 0:no, null:unknown} | integer | nullable | y.isin([0,1]) | code_rbd10_nervous_system_disease | identical | |
| 0 | RBD Screening Questionnaire | 25 | rbd_summary_score | RBD Summary Score | integer | nullable | (y>=0) & (y<=13) | rbd_summary_score | identical | |
| 0 | RBD Single-Question Screen | 1 | rbd1q | A single question measure screening for REM sleep behavior disorder | string | nullable | ['No', 'Yes', 'Unknown'] | | | |
| 0 | Epworth Sleepiness Scale | 1 | ess_info_source | ESS Source Of Information | string | nullable | | ess_info_source | identical | |
| 0 | Epworth Sleepiness Scale | 2 | code_ess0101_sitting_and_reading | ESS - Sitting And Reading (ESS0101) {0:"would never doze or sleep", 1:"slight chance of dozing or sleeping", 2:"moderate chance of dozing or sleeping", 3:"high chance of dozing or sleeping"} | integer | nullable | y.isin([0,1,2,3]) | code_ess0101_sitting_and_reading | identical | |
| 0 | Epworth Sleepiness Scale | 3 | code_ess0102_watching_tv | ESS - Watching TV (ESS0102) {0:"would never doze or sleep", 1:"slight chance of dozing or sleeping", 2:"moderate chance of dozing or sleeping", 3:"high chance of dozing or sleeping"} | integer | nullable | y.isin([0,1,2,3]) | code_ess0102_watching_tv | identical | |
| 0 | Epworth Sleepiness Scale | 4 | code_ess0103_sitting_inactive_in_public_place | ESS - Sitting, Inactive In A Public Place (ESS0103) {0:"would never doze or sleep", 1:"slight chance of dozing or sleeping", 2:"moderate chance of dozing or sleeping", 3:"high chance of dozing or sleeping"} | integer | nullable | y.isin([0,1,2,3]) | code_ess0103_sitting_inactive_in_public_place | identical | |
| 0 | Epworth Sleepiness Scale | 5 | code_ess0104_passenger_in_car_for_hour | ESS - As A Passenger In A Car For An Hour Without A Break (ESS0104) {0:"would never doze or sleep", 1:"slight chance of dozing or sleeping", 2:"moderate chance of dozing or sleeping", 3:"high chance of dozing or sleeping"} | integer | nullable | y.isin([0,1,2,3]) | code_ess0104_passenger_in_car_for_hour | identical | |
| 0 | Epworth Sleepiness Scale | 6 | code_ess0105_lying_down_to_rest_in_afternoon | ESS - Lying Down To Rest In The Afternoon When Circumstances Permit (ESS0105) {0:"would never doze or sleep", 1:"slight chance of dozing or sleeping", 2:"moderate chance of dozing or sleeping", 3:"high chance of dozing or sleeping"} | integer | nullable | y.isin([0,1,2,3]) | code_ess0105_lying_down_to_rest_in_afternoon | identical | |
| 0 | Epworth Sleepiness Scale | 7 | code_ess0106_sitting_and_talking_to_someone | ESS - Sitting And Talking To Someone (ESS0106) {0:"would never doze or sleep", 1:"slight chance of dozing or sleeping", 2:"moderate chance of dozing or sleeping", 3:"high chance of dozing or sleeping"} | integer | nullable | y.isin([0,1,2,3]) | code_ess0106_sitting_and_talking_to_someone | identical | |
| 0 | Epworth Sleepiness Scale | 8 | code_ess0107_sitting_after_lunch | ESS - Sitting Quietly After A Lunch Without Alcohol (ESS0107) {0:"would never doze or sleep", 1:"slight chance of dozing or sleeping", 2:"moderate chance of dozing or sleeping", 3:"high chance of dozing or sleeping"} | integer | nullable | y.isin([0,1,2,3]) | code_ess0107_sitting_after_lunch | identical | |
| 0 | Epworth Sleepiness Scale | 9 | code_ess0108_car_stopped_in_traffic | ESS - In A Car, While Stopped For A Few Minutes In The Traffic (ESS0108) {0:"would never doze or sleep", 1:"slight chance of dozing or sleeping", 2:"moderate chance of dozing or sleeping", 3:"high chance of dozing or sleeping"} | integer | nullable | y.isin([0,1,2,3]) | code_ess0108_car_stopped_in_traffic | identical | |
| 0 | Epworth Sleepiness Scale | | ess_total_score | ESS - Total score | integer | nullable | (y>=0) & (y<=24) | | | |
| 0 | Geriatric Depression Scale: Short Form | 1 | code_gds01_satisfied_with_life | GDS15 - Are you basically satisfied with your life? {"yes":0, "no":1} | integer | nullable | y.isin([0,1]) | | | |
| 0 | Geriatric Depression Scale: Short Form | 2 | code_gds02_drop_interests | GDS15 - Have you dropped many of your activities and interests? {"yes":1, "no":0} | integer | nullable | y.isin([0,1]) | | | |
| 0 | Geriatric Depression Scale: Short Form | 3 | code_gds03_empty | GDS15 - Do you feel that your life is empty? {"yes":1, "no":0} | integer | nullable | y.isin([0,1]) | | | |
| 0 | Geriatric Depression Scale: Short Form | 4 | code_gds04_bored | GDS15 - Do you often get bored? {"yes":1, "no":0} | integer | nullable | y.isin([0,1]) | | | |
| 0 | Geriatric Depression Scale: Short Form | 5 | code_gds05_good_spirits | GDS15 - Are you in good spirits most of the time? {"yes":0, "no":1} | integer | nullable | y.isin([0,1]) | | | |
| 0 | Geriatric Depression Scale: Short Form | 6 | code_gds06_afraid | GDS15 - Are you afraid that something bad is going to happen to you? {"yes":1, "no":0} | integer | nullable | y.isin([0,1]) | | | |
| 0 | Geriatric Depression Scale: Short Form | 7 | code_gds07_happy | GDS15 - Do you feel happy most of the time? {"yes":0, "no":1} | integer | nullable | y.isin([0,1]) | | | |
| 0 | Geriatric Depression Scale: Short Form | 8 | code_gds08_helpless | GDS15 - Do you often feel helpless? {"yes":1, "no":0} | integer | nullable | y.isin([0,1]) | | | |
| 0 | Geriatric Depression Scale: Short Form | 9 | code_gds09_home_than_going_out | GDS15 - Do you prefer to stay at home, rather than going out and doing new things? {"yes":1, "no":0} | integer | nullable | y.isin([0,1]) | | | |
| 0 | Geriatric Depression Scale: Short Form | 10 | code_gds10_memory_problems | GDS15 - Do you feel you have more problems with memory than most people? {"yes":1, "no":0} | integer | nullable | y.isin([0,1]) | | | |
| 0 | Geriatric Depression Scale: Short Form | 11 | code_gds11_wonderful_to_be_alive | GDS15 - Do you think it is wonderful to be alive? {"yes":0, "no":1} | integer | nullable | y.isin([0,1]) | | | |
| 0 | Geriatric Depression Scale: Short Form | 12 | code_gds12_worthless | GDS15 - Do you feel pretty worthless the way you are now? {"yes":1, "no":0} | integer | nullable | y.isin([0,1]) | | | |
| 0 | Geriatric Depression Scale: Short Form | 13 | code_gds13_full_of_energy | GDS15 - Do you feel full of energy? {"yes":0, "no":1} | integer | nullable | y.isin([0,1]) | | | |
| 0 | Geriatric Depression Scale: Short Form | 14 | code_gds14_hopless | GDS15 - Do you feel that your situation is hopeless? {"yes":1, "no":0} | integer | nullable | y.isin([0,1]) | | | |
| 0 | Geriatric Depression Scale: Short Form | 15 | code_gds15_not_better_off | GDS15 - Do you think that most people are better off than you? {"yes":1, "no":0} | integer | nullable | y.isin([0,1]) | | | |
| 0 | Geriatric Depression Scale: Short Form | 16 | gds15_total_score | GDS 15 - Total score | integer | nullable | (y>=0) & (y<=15) | | | |
| 0 | Orthostatic hypotension | 1 | sbp_standing | Systolic blood pressure at standing position | numeric | nullable | (y>=0) & (y<=300) | | | |
| 0 | Orthostatic hypotension | 2 | dbp_standing | Diastolic blood pressure at standing position | numeric | nullable | (y>=0) & (y<=300) | | | |
| 0 | Orthostatic hypotension | 3 | hr_standing | Heart rate at standing position | numeric | nullable | (y>=0) & (y<=200) | | | |
| 0 | Orthostatic hypotension | 4 | sbp_supine | Systolic blood pressure at supine position | numeric | nullable | (y>=0) & (y<=300) | | | |
| 0 | Orthostatic hypotension | 5 | dbp_supine | Diastolic blood pressure at supine position | numeric | nullable | (y>=0) & (y<=300) | | | |
| 0 | Orthostatic hypotension | 6 | hr_supine | Heart rate at supine position | numeric | nullable | (y>=0) & (y<=200) | | | |
| 0 | Orthostatic hypotension | 7 | sbp_sitting | Systolic blood pressure at sitting position | numeric | nullable | (y>=0) & (y<=300) | | | |
| 0 | Orthostatic hypotension | 8 | dbp_sitting | Diastolic blood pressure at sitting position | numeric | nullable | (y>=0) & (y<=300) | | | |
| 0 | Orthostatic hypotension | 9 | hr_sitting | Heart rate at sitting position | numeric | nullable | (y>=0) & (y<=200) | | | |
| 0 | Schwab England ADL | 1 | schwab_england_pct_adl_score | Schwab And England Percent ADL Score | numeric | nullable | (y>=0) & (y<=100) | | | |
| 0 | Olfactory test | 1 | smell_test_results | Anosmia, hyposmia, norsomnia based on the age and sex standardized cut-off | string | nullable | ["Anosmia", "Hyposmia", "Normosmia"] | | | |
| 0 | Olfactory test | 2 | smell_test_name | Name of the used smell test (e.g. sniffin-stick, bsit, upsit) | string | nullable | | | | |
| 0 | Olfactory test | 3 | smell_test_score | Score of the smell test | numeric | nullable | (y>=0) & (y<=9999) | | | |
| 0 | Misc Depression Scale | 1 | is_depressed | Does the patient have depression based on the cut-off score of the scale used? | string | nullable | ["Yes","No","Unknown"] | | | |
| 0 | Misc Depression Scale | 2 | depress_test_name | Test used for depression screening e.g. GDS15, HDRS, BDI, PHQ9 (DPUK),GHQ | string | nullable | | | | |
| 0 | Misc Depression Scale | 3 | depress_test_score | Total score on the depression scale used | numeric | nullable | (y>=0) & (y<=100) | | | |
| 0 | PDQ-39 | 1 | pdq39_01_doing_leisure_activity | PDQ-39-Q1: Had difficulty doing the leisure activities you would like to do? | string | nullable | ["Never", "Occasionally", "Sometimes", "Often", | pdq39_01_doing_leisure_activity | identical | |
| 0 | PDQ-39 | 2 | pdq39_02_looking_after_home | PDQ-39-Q2: Had difficulty looking after your home, for example, housework, cooking or yardwork? | string | nullable | ["Never", "Occasionally", "Sometimes", "Often", | pdq39_02_looking_after_home | identical | |
| 0 | PDQ-39 | 3 | pdq39_03_carrying_shopping_bags | PDQ-39-Q3: Had difficulty carrying grocery bags? | string | nullable | ["Never", "Occasionally", "Sometimes", "Often", | pdq39_03_carrying_shopping_bags | identical | |
| 0 | PDQ-39 | 4 | pdq39_04_walking_half_mile | PDQ-39-Q4: Had problems walking half a mile? | string | nullable | ["Never", "Occasionally", "Sometimes", "Often", | pdq39_04_walking_half_mile | identical | |
| 0 | PDQ-39 | 5 | pdq39_05_walking_100_yards | PDQ-39-Q5: Had problems walking 100 yards (approximately 1 block)? | string | nullable | ["Never", "Occasionally", "Sometimes", "Often", | pdq39_05_walking_100_yards | identical | |
| 0 | PDQ-39 | 6 | pdq39_06_getting_around_house | PDQ-39-Q6: Had problems getting around the house as easily as you would like? | string | nullable | ["Never", "Occasionally", "Sometimes", "Often", | pdq39_06_getting_around_house | identical | |
| 0 | PDQ-39 | 7 | pdq39_07_getting_around_in_public | PDQ-39-Q7: Had difficulty getting around in public places? | string | nullable | ["Never", "Occasionally", "Sometimes", "Often", | pdq39_07_getting_around_in_public | identical | |
| 0 | PDQ-39 | 8 | pdq39_08_need_someone_to_accompany | PDQ-39-Q8: Needed someone else to accompany you when you went out? | string | nullable | ["Never", "Occasionally", "Sometimes", "Often", | pdq39_08_need_someone_to_accompany | identical | |
| 0 | PDQ-39 | 9 | pdq39_09_worried_about_falling | PDQ-39-Q9: Felt frightened or worried about falling in public? | string | nullable | ["Never", "Occasionally", "Sometimes", "Often", | pdq39_09_worried_about_falling | identical | |
| 0 | PDQ-39 | 10 | pdq39_10_confined_to_house | PDQ-39-Q10: Been confined to the house more than you would like? | string | nullable | ["Never", "Occasionally", "Sometimes", "Often", | pdq39_10_confined_to_house | identical | |
| 0 | PDQ-39 | 11 | pdq39_11_showering | PDQ-39-Q11: Had difficulty showering and bathing? | string | nullable | ["Never", "Occasionally", "Sometimes", "Often", | pdq39_11_showering | identical | |
| 0 | PDQ-39 | 12 | pdq39_12_dressing | PDQ-39-Q12: Had difficulty dressing? | string | nullable | ["Never", "Occasionally", "Sometimes", "Often", | pdq39_12_dressing | identical | |

Global Parkinson's Genetics Program

#### Supplementary Table 2. GP2 Core Data Dictionary (continued)

| Single measure | Modality | no. | Item | Description | ItemType | Required | Values | AMP_PD_item | Conversion | Comment |
|---|---|---|---|---|---|---|---|---|---|---|
| 0 | PDQ-39 | 13 | pdq39_13_buttons_and_shoelaces | PDQ-39-Q13: Had difficulty with buttons or shoelaces? | string | nullable | ["Never", "Occasionally", "Sometimes", "Often", | pdq39_13_buttons_and_shoelaces | identical | |
| 0 | PDQ-39 | 14 | pdq39_14_writing | PDQ-39-Q14: Had problems writing clearly? | string | nullable | ["Never", "Occasionally", "Sometimes", "Often", | pdq39_14_writing | identical | |
| 0 | PDQ-39 | 15 | pdq39_15_cutting_food | PDQ-39-Q15: Had difficulty cutting up your food? | string | nullable | ["Never", "Occasionally", "Sometimes", "Often", | pdq39_15_cutting_food | identical | |
| 0 | PDQ-39 | 16 | pdq39_16_spill_drink | PDQ-39-Q16: Had difficulty holding a drink without spilling it? | string | nullable | ["Never", "Occasionally", "Sometimes", "Often", | pdq39_16_spill_drink | identical | |
| 0 | PDQ-39 | 17 | pdq39_17_depressed | PDQ-39-Q17: Felt depressed? | string | nullable | ["Never", "Occasionally", "Sometimes", "Often", | pdq39_17_depressed | identical | |
| 0 | PDQ-39 | 18 | pdq39_18_lonely | PDQ-39-Q18: Felt isolated and lonely? | string | nullable | ["Never", "Occasionally", "Sometimes", "Often", | pdq39_18_lonely | identical | |
| 0 | PDQ-39 | 19 | pdq39_19_weepy | PDQ-39-Q19: Felt weepy or tearful? | string | nullable | ["Never", "Occasionally", "Sometimes", "Often", | pdq39_19_weepy | identical | |
| 0 | PDQ-39 | 20 | pdq39_20_angry | PDQ-39-Q20: Felt angry or bitter? | string | nullable | ["Never", "Occasionally", "Sometimes", "Often", | pdq39_20_angry | identical | |
| 0 | PDQ-39 | 21 | pdq39_21_anxious | PDQ-39-Q21: Felt anxious? | string | nullable | ["Never", "Occasionally", "Sometimes", "Often", | pdq39_21_anxious | identical | |
| 0 | PDQ-39 | 22 | pdq39_22_worried_about_future | PDQ-39-Q22: Felt worried about your future? | string | nullable | ["Never", "Occasionally", "Sometimes", "Often", | pdq39_22_worried_about_future | identical | |
| 0 | PDQ-39 | 23 | pdq39_23_hide_pd_from_people | PDQ-39-Q23: Felt you had to hide your Parkinson's from people? | string | nullable | ["Never", "Occasionally", "Sometimes", "Often", | pdq39_23_hide_pd_from_people | identical | |
| 0 | PDQ-39 | 24 | pdq39_24_avoid_eat_drink_in_public | PDQ-39-Q24: Avoided situations which involve eating or drinking in public? | string | nullable | ["Never", "Occasionally", "Sometimes", "Often", | pdq39_24_avoid_eat_drink_in_public | identical | |
| 0 | PDQ-39 | 25 | pdq39_25_embarassed_in_public | PDQ-39-Q25: Felt embarrassed in public? | string | nullable | ["Never", "Occasionally", "Sometimes", "Often", | pdq39_25_embarassed_in_public | identical | |
| 0 | PDQ-39 | 26 | pdq39_26_worried_about_reactions | PDQ-39-Q26: Felt worried about other people's reaction to you? | string | nullable | ["Never", "Occasionally", "Sometimes", "Often", | pdq39_26_worried_about_reactions | identical | |
| 0 | PDQ-39 | 27 | pdq39_27_close_personal_relations | PDQ-39-Q27: Had problems with your close personal relationships? | string | nullable | ["Never", "Occasionally", "Sometimes", "Often", | pdq39_27_close_personal_relations | identical | |
| 0 | PDQ-39 | 28 | pdq39_28_support_from_spouse | PDQ-39-Q28: Lacked the support you needed from your spouse or partner? | string | nullable | ["Never", "Occasionally", "Sometimes", "Often", | pdq39_28_support_from_spouse | identical | |
| 0 | PDQ-39 | 29 | pdq39_29_support_from_family | PDQ-39-Q29: Lacked the support you needed from your family or close friends? | string | nullable | ["Never", "Occasionally", "Sometimes", "Often", | pdq39_29_support_from_family | identical | |
| 0 | PDQ-39 | 30 | pdq39_30_sleep_in_day | PDQ-39-Q30: Unexpectedly fallen asleep during the day? | string | nullable | ["Never", "Occasionally", "Sometimes", "Often", | pdq39_30_sleep_in_day | identical | |
| 0 | PDQ-39 | 31 | pdq39_31_problem_with_concentration | PDQ-39-Q31: Had problems with your concentration, for example when reading or watching TV? | string | nullable | ["Never", "Occasionally", "Sometimes", "Often", | pdq39_31_problem_with_concentration | identical | |
| 0 | PDQ-39 | 32 | pdq39_32_memory_is_failing | PDQ-39-Q32: Felt your memory was failing? | string | nullable | ["Never", "Occasionally", "Sometimes", "Often", | pdq39_32_memory_is_failing | identical | |
| 0 | PDQ-39 | 33 | pdq39_33_hallucinations | PDQ-39-Q33: Had distressing dreams or hallucinations? | string | nullable | ["Never", "Occasionally", "Sometimes", "Often", | pdq39_33_hallucinations | identical | |
| 0 | PDQ-39 | 34 | pdq39_34_speaking | PDQ-39-Q34: Had difficulty speaking? | string | nullable | ["Never", "Occasionally", "Sometimes", "Often", | pdq39_34_speaking | identical | |
| 0 | PDQ-39 | 35 | pdq39_35_unable_to_communicate | PDQ-39-Q35: Felt unable to communicate effectively? | string | nullable | ["Never", "Occasionally", "Sometimes", "Often", | pdq39_35_unable_to_communicate | identical | |
| 0 | PDQ-39 | 36 | pdq39_36_felt_ignored | PDQ-39-Q36: Felt ignored by people? | string | nullable | ["Never", "Occasionally", "Sometimes", "Often", | pdq39_36_felt_ignored | identical | |
| 0 | PDQ-39 | 37 | pdq39_37_muscle_cramps | PDQ-39-Q37: Had painful muscle cramps or spasms? | string | nullable | ["Never", "Occasionally", "Sometimes", "Often", | pdq39_37_muscle_cramps | identical | |
| 0 | PDQ-39 | 38 | pdq39_38_joint_pains | PDQ-39-Q38: Had aches and pains in your joints or body? | string | nullable | ["Never", "Occasionally", "Sometimes", "Often", | pdq39_38_joint_pains | identical | |
| 0 | PDQ-39 | 39 | pdq39_39_hot_or_cold | PDQ-39-Q39: Felt uncomfortably hot or cold? | string | nullable | ["Never", "Occasionally", "Sometimes", "Often", | pdq39_39_hot_or_cold | identical | |
| 0 | PDQ-39 | 40 | pdq39_mobility_score | PDQ-39-Total Score-Mobility (item 1-10) | integer | nullable | (y>=0) & (y<=40) | pdq39_mobility_score | identical | |
| 0 | PDQ-39 | 41 | pdq39_adl_score | PDQ-39-Total Score-Activities Of Daily Living (ADL, item 11-16) | integer | nullable | (y>=0) & (y<=24) | pdq39_adl_score | identical | |
| 0 | PDQ-39 | 42 | pdq39_emotional_score | PDQ-39-Total Score-Emotional Well Being (item 17-22, max ) | integer | nullable | (y>=0) & (y<=24) | pdq39_emotional_score | identical | |
| 0 | PDQ-39 | 43 | pdq39_stigma_score | PDQ-39-Total Score-Stigma (item 23-26) | integer | nullable | (y>=0) & (y<=16) | pdq39_stigma_score | identical | |
| 0 | PDQ-39 | 44 | pdq39_social_score | PDQ-39-Total Score-Social Support (item 27-29) | integer | nullable | (y>=0) & (y<=12) | pdq39_social_score | identical | |
| 0 | PDQ-39 | 45 | pdq39_cognition_score | PDQ-39-Total Score-Cognitive Impairment (Cognitions, item 30-33) | integer | nullable | (y>=0) & (y<=16) | pdq39_cognition_score | identical | |
| 0 | PDQ-39 | 46 | pdq39_communication_score | PDQ-39-Total Score-Communication (item 34-36) | integer | nullable | (y>=0) & (y<=12) | pdq39_communication_score | identical | |
| 0 | PDQ-39 | 47 | pdq39_discomfort_score | PDQ-39-Total Score-Bodily Discomfort (item 37-39) | integer | nullable | (y>=0) & (y<=12) | pdq39_discomfort_score | identical | |
| 0 | PDQ-8 | 1 | pdq8_total_score | PDQ-8 (Short form of PDQ-39) Total Score | integer | nullable | (y>=0) & (y<=32) | | | |
| 0 | WBC counts | 1 | neutrophil_count | /microL | numeric | nullable | (y>=0) & (y<=100000) | | | |
| 0 | WBC counts | 2 | lymphocyte_count | /microL | numeric | nullable | (y>=0) & (y<=100000) | | | |
| 0 | WBC counts | 3 | eosinophil_count | /microL | numeric | nullable | (y>=0) & (y<=100000) | | | |
| 0 | WBC counts | 4 | basophil_count | /microL | numeric | nullable | (y>=0) & (y<=100000) | | | |
| 0 | WBC counts | 5 | monocyte_count | /microL | numeric | nullable | (y>=0) & (y<=100000) | | | |
| 0 | King's PD pain scale | 1 | king_01_joints_severity | | integer | nullable | y.isin([0,1,2,3]) | | | |
| 0 | King's PD pain scale | 2 | king_01_joints_freq | | integer | nullable | y.isin([0,1,2,3,4]) | | | |
| 0 | King's PD pain scale | 3 | king_02_deep_body_severity | | integer | nullable | y.isin([0,1,2,3]) | | | |
| 0 | King's PD pain scale | 4 | king_02_deep_body_freq | | integer | nullable | y.isin([0,1,2,3,4]) | | | |
| 0 | King's PD pain scale | 5 | king_03_orgain_severity | | integer | nullable | y.isin([0,1,2,3]) | | | |
| 0 | King's PD pain scale | 6 | king_03_orgain_freq | | integer | nullable | y.isin([0,1,2,3,4]) | | | |
| 0 | King's PD pain scale | 7 | king_04_dyskinetic_severity | | integer | nullable | y.isin([0,1,2,3]) | | | |
| 0 | King's PD pain scale | 8 | king_04_dyskinetic_freq | | integer | nullable | y.isin([0,1,2,3,4]) | | | |
| 0 | King's PD pain scale | 9 | king_05_offdystonia_specific_severity | | integer | nullable | y.isin([0,1,2,3]) | | | |
| 0 | King's PD pain scale | 10 | king_05_offdystonia_specific_freq | | integer | nullable | y.isin([0,1,2,3,4]) | | | |
| 0 | King's PD pain scale | 11 | king_06_offdystonia_general_severity | | integer | nullable | y.isin([0,1,2,3]) | | | |
| 0 | King's PD pain scale | 12 | king_06_offdystonia_general_freq | | integer | nullable | y.isin([0,1,2,3,4]) | | | |
| 0 | King's PD pain scale | 13 | king_07_leg_movement_night_severity | | integer | nullable | y.isin([0,1,2,3]) | | | |
| 0 | King's PD pain scale | 14 | king_07_leg_movement_night_freq | | integer | nullable | y.isin([0,1,2,3,4]) | | | |
| 0 | King's PD pain scale | 15 | king_08_turing_in_bed_severity | | integer | nullable | y.isin([0,1,2,3]) | | | |
| 0 | King's PD pain scale | 16 | king_08_turing_in_bed_freq | | integer | nullable | y.isin([0,1,2,3,4]) | | | |
| 0 | King's PD pain scale | 17 | king_09_chewing_severity | | integer | nullable | y.isin([0,1,2,3]) | | | |
| 0 | King's PD pain scale | 18 | king_09_chewing_freq | | integer | nullable | y.isin([0,1,2,3,4]) | | | |
| 0 | King's PD pain scale | 19 | king_10_grinding_teeth_night_severity | | integer | nullable | y.isin([0,1,2,3]) | | | |
| 0 | King's PD pain scale | 20 | king_10_grinding_teeth_night_freq | | integer | nullable | y.isin([0,1,2,3,4]) | | | |
| 0 | King's PD pain scale | 21 | king_11_burning_mouth_severity | | integer | nullable | y.isin([0,1,2,3]) | | | |
| 0 | King's PD pain scale | 22 | king_11_burning_mouth_freq | | integer | nullable | y.isin([0,1,2,3,4]) | | | |
| 0 | King's PD pain scale | 23 | king_12_burning_limbs_severity | | integer | nullable | y.isin([0,1,2,3]) | | | |
| 0 | King's PD pain scale | 24 | king_12_burning_limbs_freq | | integer | nullable | y.isin([0,1,2,3,4]) | | | |
| 0 | King's PD pain scale | 25 | king_13_lower_abs_severity | | integer | nullable | y.isin([0,1,2,3]) | | | |
| 0 | King's PD pain scale | 26 | king_13_lower_abs_freq | | integer | nullable | y.isin([0,1,2,3,4]) | | | |
| 0 | King's PD pain scale | 27 | king_14_shooting_limbs_severity | | integer | nullable | y.isin([0,1,2,3]) | | | |
| 0 | King's PD pain scale | 28 | king_14_shooting_limbs_freq | | integer | nullable | y.isin([0,1,2,3,4]) | | | |
| 0 | King's PD pain scale | 29 | king_muscleskeletal | | integer | nullable | (y>=0) & (y<=12) | | | |
| 0 | King's PD pain scale | 30 | king_chronic | | integer | nullable | (y>=0) & (y<=24) | | | |
| 0 | King's PD pain scale | 31 | king_fluctuation_related | | integer | nullable | (y>=0) & (y<=36) | | | |
| 0 | King's PD pain scale | 32 | king_nocturnal | | integer | nullable | (y>=0) & (y<=24) | | | |
| 0 | King's PD pain scale | 33 | king_orofacial | | integer | nullable | (y>=0) & (y<=36) | | | |
| 0 | King's PD pain scale | 34 | king_discolouration | | integer | nullable | (y>=0) & (y<=24) | | | |
| 0 | King's PD pain scale | 35 | king_radicular | | integer | nullable | (y>=0) & (y<=12) | | | |
| 0 | MMSE | 1 | mmse_01_year | MMSE 1 | integer | nullable | y.isin([0,1]) | | | |
| 0 | MMSE | 2 | mmse_02_season | MMSE 2 | integer | nullable | y.isin([0,1]) | | | |
| 0 | MMSE | 3 | mmse_03_date | MMSE 3 | integer | nullable | y.isin([0,1]) | | | |
| 0 | MMSE | 4 | mmse_04_day | MMSE 4 | integer | nullable | y.isin([0,1]) | | | |

#### Supplementary Table 2. GP2 Core Data Dictionary (continued)

| Single measure | Modality | no. | Item | Description | ItemType | Required | Values | AMP_PD_item | Conversion | Comment |
|---|---|---|---|---|---|---|---|---|---|---|
| 0 | MMSE | 5 | mmse_05_month | MMSE 5 | integer | nullable | y.isin([0,1]) | | x[[mmse_01 | |
| 0 | MMSE | 31 | mmse_orientation_time | x[["mmse_01_year","mmse_02_season","mmse_03_date","mmse_04_day","mmse_05_month"]].sum(axis=1) | integer | nullable | (y>=0) & (y<=5) | | | |
| 0 | MMSE | 6 | mmse_06_state | MMSE 6 | integer | nullable | y.isin([0,1]) | | | |
| 0 | MMSE | 7 | mmse_07_county | MMSE 7 | integer | nullable | y.isin([0,1]) | | | |
| 0 | MMSE | 8 | mmse_08_town | MMSE 8 | integer | nullable | y.isin([0,1]) | | | |
| 0 | MMSE | 9 | mmse_09_hospital | MMSE 9 | integer | nullable | y.isin([0,1]) | | | |
| 0 | MMSE | 10 | mmse_10_floor | MMSE 10 | integer | nullable | y.isin([0,1]) | | | |
| 0 | MMSE | 32 | mmse_orientation_place | sum MMSE 6-10 | integer | nullable | (y>=0) & (y<=5) | | | |
| 0 | MMSE | 11 | mmse_11_immediate_recall_1 | MMSE 11 | integer | nullable | y.isin([0,1]) | | | |
| 0 | MMSE | 12 | mmse_12_immediate_recall_2 | MMSE 12 | integer | nullable | y.isin([0,1]) | | | |
| 0 | MMSE | 13 | mmse_13_immediate_recall_3 | MMSE 13 | integer | nullable | y.isin([0,1]) | | | |
| 0 | MMSE | 33 | mmse_immediate_recall | sum MMSE 11-13 | integer | nullable | (y>=0) & (y<=3) | | | |
| 0 | MMSE | 14 | mmse_14_serial7_1 | MMSE 14 | integer | nullable | y.isin([0,1]) | | | |
| 0 | MMSE | 15 | mmse_15_serial7_2 | MMSE 15 | integer | nullable | y.isin([0,1]) | | | |
| 0 | MMSE | 16 | mmse_16_serial7_3 | MMSE 16 | integer | nullable | y.isin([0,1]) | | | |
| 0 | MMSE | 17 | mmse_17_serial7_4 | MMSE 17 | integer | nullable | y.isin([0,1]) | | | |
| 0 | MMSE | 18 | mmse_18_serial7_5 | MMSE 18 | integer | nullable | y.isin([0,1]) | | | |
| 0 | MMSE | 34 | mmse_serial7 | sum MMSE 14-18 | integer | nullable | (y>=0) & (y<=5) | | | |
| 0 | MMSE | 19 | mmse_19_delayed_recall_1 | MMSE 19 | integer | nullable | y.isin([0,1]) | | | |
| 0 | MMSE | 20 | mmse_20_delayed_recall_2 | MMSE 20 | integer | nullable | y.isin([0,1]) | | | |
| 0 | MMSE | 21 | mmse_21_delayed_recall_3 | MMSE 21 | integer | nullable | y.isin([0,1]) | | | |
| 0 | MMSE | 35 | mmse_delayed_recall | sum MMSE 19-21 | integer | nullable | (y>=0) & (y<=3) | | | |
| 0 | MMSE | 22 | mmse_22_naming_1 | MMSE 22 | integer | nullable | y.isin([0,1]) | | | |
| 0 | MMSE | 23 | mmse_23_naming_2 | MMSE 23 | integer | nullable | y.isin([0,1]) | | | |
| 0 | MMSE | 36 | mmse_naming | sum MMSE 22-23 | integer | nullable | y.isin([0,1, 2]) | | | |
| 0 | MMSE | 24 | mmse_24_repeating | MMSE 24 | integer | nullable | y.isin([0,1]) | | | |
| 0 | MMSE | 25 | mmse_25_oral_command_1 | MMSE 25 | integer | nullable | y.isin([0,1]) | | | |
| 0 | MMSE | 26 | mmse_26_oral_command_2 | MMSE 26 | integer | nullable | y.isin([0,1]) | | | |
| 0 | MMSE | 27 | mmse_27_oral_command_3 | MMSE 27 | integer | nullable | y.isin([0,1]) | | | |
| 0 | MMSE | 37 | mmse_comprehension | sum MMSE 25-27 | integer | nullable | (y>=0) & (y<=3) | | | |
| 0 | MMSE | 28 | mmse_28_reading | MMSE 28 | integer | nullable | y.isin([0,1]) | | | |
| 0 | MMSE | 29 | mmse_29_writing | MMSE 29 | integer | nullable | y.isin([0,1]) | | | |
| 0 | MMSE | 30 | mmse_30_drawing | MMSE 30 | integer | nullable | y.isin([0,1]) | | | |
| 0 | MMSE | 38 | mmse_total_score | Total | integer | nullable | (y>=0) & (y<=30) | | | |
| 0 | UPDRS Part I | 1 | code_upd101_intellectual_impairment | Mentation:Intellectual Impairment | integer | nullable | y.isin([0,1,2,3,4]) | | | |
| 0 | UPDRS Part I | 2 | code_upd102_thought_disorder | Mentation:Thought Disorder | integer | nullable | y.isin([0,1,2,3,4]) | | | |
| 0 | UPDRS Part I | 3 | code_upd103_depression | Mentation:Depression | integer | nullable | y.isin([0,1,2,3,4]) | | | |
| 0 | UPDRS Part I | 4 | code_upd104_motivation | Mentation:Motivation/Initiative | integer | nullable | y.isin([0,1,2,3,4]) | | | |
| 0 | UPDRS Part II | 5 | code_upd105_speech | Activities:Speech | integer | nullable | y.isin([0,1,2,3,4]) | | | |
| 0 | UPDRS Part II | 6 | code_upd106_salivation | Activities:Salivation | integer | nullable | y.isin([0,1,2,3,4]) | | | |
| 0 | UPDRS Part II | 7 | code_upd107_swallowing | Activities:Swallowing | integer | nullable | y.isin([0,1,2,3,4]) | | | |
| 0 | UPDRS Part II | 8 | code_upd108_handwriting | Activities:Handwriting | integer | nullable | y.isin([0,1,2,3,4]) | | | |
| 0 | UPDRS Part II | 9 | code_upd109_eating_tasks | Activities:Cut Food/Handle Utensil | integer | nullable | y.isin([0,1,2,3,4]) | | | |
| 0 | UPDRS Part II | 10 | code_upd110_dressing | Activities:Dressing | integer | nullable | y.isin([0,1,2,3,4]) | | | |
| 0 | UPDRS Part II | 11 | code_upd111_hygiene | Activities:Hygiene | integer | nullable | y.isin([0,1,2,3,4]) | | | |
| 0 | UPDRS Part II | 12 | code_upd112_bed | Activities:Turn Bed/Adj Clothes | integer | nullable | y.isin([0,1,2,3,4]) | | | |
| 0 | UPDRS Part II | 13 | code_upd113_falling | Activities:Falling | integer | nullable | y.isin([0,1,2,3,4]) | | | |
| 0 | UPDRS Part II | 14 | code_upd114_freezing_of_gait | Activities:Freezing When Walking | integer | nullable | y.isin([0,1,2,3,4]) | | | |
| 0 | UPDRS Part II | 15 | code_upd115_walking | Activities:Walking | integer | nullable | y.isin([0,1,2,3,4]) | | | |
| 0 | UPDRS Part II | 16 | code_upd116_tremor | Activities:Tremor | integer | nullable | y.isin([0,1,2,3,4]) | | | |
| 0 | UPDRS Part II | 17 | code_upd117_sensory_complaints | Activities:Sensory Complaints | integer | nullable | y.isin([0,1,2,3,4]) | | | |
| 0 | UPDRS Part III | 18 | code_upd118_speech | Motor:Speech | integer | nullable | y.isin([0,1,2,3,4]) | | | |
| 0 | UPDRS Part III | 19 | code_upd119_facial_expression | Motor:Facial Expression | integer | nullable | y.isin([0,1,2,3,4]) | | | |
| 0 | UPDRS Part III | 20 | code_upd120_rest_tremor | Motor:Tremor at Rest | integer | nullable | y.isin([0,1,2,3,4]) | | | |
| 0 | UPDRS Part III | 21 | code_upd120a_rest_tremor_right_upper_extremity | Motor:Action/Postural Hand Tremor | integer | nullable | y.isin([0,1,2,3,4]) | | | |
| 0 | UPDRS Part III | 22 | code_upd120b_rest_tremor_left_upper_extremity | | integer | nullable | y.isin([0,1,2,3,4]) | | | |
| 0 | UPDRS Part III | 23 | code_upd120c_rest_tremor_right_lower_extremity | | integer | nullable | y.isin([0,1,2,3,4]) | | | |
| 0 | UPDRS Part III | 24 | code_upd120d_rest_tremor_left_lower_extremity | | integer | nullable | y.isin([0,1,2,3,4]) | | | |
| 0 | UPDRS Part III | 25 | code_upd120e_rest_tremor_lip_or_jaw | | integer | nullable | y.isin([0,1,2,3,4]) | | | |
| 0 | UPDRS Part III | 26 | code_upd121_action_or_postural_tremor | Motor:Action/Postural Hand Tremor | integer | nullable | y.isin([0,1,2,3,4]) | | | |
| 0 | UPDRS Part III | 27 | code_upd121a_action_or_postural_tremor_right | | integer | nullable | y.isin([0,1,2,3,4]) | | | |
| 0 | UPDRS Part III | 28 | code_upd121b_action_or_postural_tremor_left | | integer | nullable | y.isin([0,1,2,3,4]) | | | |
| 0 | UPDRS Part III | 29 | code_upd122_rigidity | Motor:Rigidity | integer | nullable | y.isin([0,1,2,3,4]) | | | |
| 0 | UPDRS Part III | 30 | code_upd122a_rigidity_neck | | integer | nullable | y.isin([0,1,2,3,4]) | | | |
| 0 | UPDRS Part III | 31 | code_upd122b_rigidity_rt_upper_extremity | | integer | nullable | y.isin([0,1,2,3,4]) | | | |
| 0 | UPDRS Part III | 32 | code_upd122c_rigidity_left_upper_extremity | | integer | nullable | y.isin([0,1,2,3,4]) | | | |
| 0 | UPDRS Part III | 33 | code_upd122d_rigidity_rt_upper_extremity | | integer | nullable | y.isin([0,1,2,3,4]) | | | |
| 0 | UPDRS Part III | 34 | code_upd122e_rigidity_left_upper_extremity | | integer | nullable | y.isin([0,1,2,3,4]) | | | |
| 0 | UPDRS Part III | 35 | code_upd123_finger_taps | Motor:Finger Taps | integer | nullable | y.isin([0,1,2,3,4]) | | | |
| 0 | UPDRS Part III | 36 | code_upd123a_right_finger_taps | | integer | nullable | y.isin([0,1,2,3,4]) | | | |
| 0 | UPDRS Part III | 37 | code_upd123b_left_finger_taps | | integer | nullable | y.isin([0,1,2,3,4]) | | | |
| 0 | UPDRS Part III | 38 | code_upd124_hand_movements | Motor:Hand Movements | integer | nullable | y.isin([0,1,2,3,4]) | | | |
| 0 | UPDRS Part III | 39 | code_upd124a_right_hand_movements | | integer | nullable | y.isin([0,1,2,3,4]) | | | |
| 0 | UPDRS Part III | 40 | code_upd124b_left_hand_movements | | integer | nullable | y.isin([0,1,2,3,4]) | | | |
| 0 | UPDRS Part III | 41 | code_upd125_pron_sup_movement | Motor:Rapid Alternating Hand Moves | integer | nullable | y.isin([0,1,2,3,4]) | | | |
| 0 | UPDRS Part III | 42 | code_upd125a_pron_sup_movement_right_hand | | integer | nullable | y.isin([0,1,2,3,4]) | | | |
| 0 | UPDRS Part III | 43 | code_upd125b_pron_sup_movement_left_hand | | integer | nullable | y.isin([0,1,2,3,4]) | | | |
| 0 | UPDRS Part III | 44 | code_upd126_leg_agility | Motor:Leg Agility | integer | nullable | y.isin([0,1,2,3,4]) | | | |
| 0 | UPDRS Part III | 45 | code_upd126a_right_leg_agility | | integer | nullable | y.isin([0,1,2,3,4]) | | | |
| 0 | UPDRS Part III | 46 | code_upd126b_left_leg_agility | | integer | nullable | y.isin([0,1,2,3,4]) | | | |

Global Parkinson's Genetics Program

### Supplementary Table 2. GP2 Core Data Dictionary (continued)

| Single measure | Modality | no. | Item | Description | ItemType | Required | Values | AMP_PD_item | Conversion | Comment |
|---|---|---|---|---|---|---|---|---|---|---|
| 0 | UPDRS Part III | 47 | code_upd127_arising_from_chair | Motor:Arising from Chair | integer | nullable | y.isin([0,1,2,3,4]) | | | |
| 0 | UPDRS Part III | 48 | code_upd128_posture | Motor:Posture | integer | nullable | y.isin([0,1,2,3,4]) | | | |
| 0 | UPDRS Part III | 49 | code_upd129_gait | Motor:Gait | integer | nullable | y.isin([0,1,2,3,4]) | | | |
| 0 | UPDRS Part III | 50 | code_upd130_postural_stability | Motor:Postural Stability | integer | nullable | y.isin([0,1,2,3,4]) | | | |
| 0 | UPDRS Part III | 51 | code_upd131_body_bradykinesia | Motor:Bradykinesia and Hypokinesia | integer | nullable | y.isin([0,1,2,3,4]) | | | |
| 0 | UPDRS Part IV | 52 | code_upd132_time_spent_with_dyskinesias | Complications:Dyskinesias Duration | integer | nullable | y.isin([0,1,2,3,4]) | | | |
| 0 | UPDRS Part IV | 53 | code_upd133_functional_impact_of_dyskinesias | Complications:Dyskinesias Disable | integer | nullable | y.isin([0,1,2,3,4]) | | | |
| 0 | UPDRS Part IV | 54 | code_upd134_painful_dyskinesias | Complications:Dyskinesias Painful | integer | nullable | y.isin([0,1,2,3,4]) | | | |
| 0 | UPDRS Part IV | 55 | code_upd135_early_morning_dystonia | Complications:Dyskinesias Dystonia | integer | nullable | y.isin([0,1]) | | | |
| 0 | UPDRS Part IV | 56 | code_upd136_predictable_off | Complications:Fluct Predictable | integer | nullable | y.isin([0,1]) | | | |
| 0 | UPDRS Part IV | 57 | code_upd137_unpredictable_off | Complications:Fluct Unpredictable | integer | nullable | y.isin([0,1]) | | | |
| 0 | UPDRS Part IV | 58 | code_upd138_sudden_off | Complications:Fluct Suddenly | integer | nullable | y.isin([0,1]) | | | |
| 0 | UPDRS Part IV | 59 | code_upd139_time_spent_in_the_off_state | Complications:Fluct Average | integer | nullable | y.isin([0,1,2,3,4]) | | | |
| 0 | UPDRS Part IV | 60 | code_upd140_anorexia | Complications:Anorex Nausea Vomit | integer | nullable | y.isin([0,1]) | | | |
| 0 | UPDRS Part IV | 61 | code_upd141_sleep_disturbances | Complications:Insomnia Hypersomnol | integer | nullable | y.isin([0,1]) | | | |
| 0 | UPDRS Part IV | 62 | code_upd142_symptomatic_orthostasis | Complications:Symptom Orthostasis | integer | nullable | y.isin([0,1]) | | | |
| 0 | UPDRS | 63 | updrs_part_i_summary_score | | integer | nullable | (y>=0) & (y<=16) | | | |
| 0 | UPDRS | 64 | updrs_part_ii_summary_score | | integer | nullable | (y>=0) & (y<=52) | | | |
| 0 | UPDRS | 65 | updrs_part_iii_summary_score | | integer | nullable | (y>=0) & (y<=136) | | | |
| 0 | UPDRS | 66 | updrs_part_iv_summary_score | | integer | nullable | (y>=0) & (y<=23) | | | |
| 0 | PD RFQ-U | 1 | pdrfuq_coffee | In your lifetime, have you ever regularly drunk caffeinated coffee, that is, at least once per week for 6 months or longer? {0:No, 1:Yes, 2:Possibly, -9:Don't Konw, -7:Refused} | string | nullable | ["No", "Yes", "Don't Know", "Refused"] | | | |
| 0 | PD RFQ-U | 2 | pdrfuq_black_tea | In your lifetime, have you ever regularly drunk hot or iced caffeinated black tea, that is, at least once per week for 6 months or longer? {0:No, 1:Yes, 2:Possibly, -9:Don't Konw, -7:Refused} | string | nullable | ["No", "Yes", "Don't Know", "Refused"] | | | |
| 0 | PD RFQ-U | 3 | pdrfuq_green_tea | In your lifetime, have you ever regularly drunk caffeinated green tea, that is, at least once per week for 6 months or longer? {0:No, 1:Yes, 2:Possibly, -9:Don't Konw, -7:Refused} | string | nullable | ["No", "Yes", "Don't Know", "Refused"] | | | |
| 0 | PD RFQ-U | 4 | pdrfuq_caffeinated_soda | In your lifetime, have you ever regularly drunk caffeinated soda, that is, at least once per week for 6 months or longer? {0:No, 1:Yes, 2:Possibly, -9:Don't Konw, -7:Refused} | string | nullable | ["No", "Yes", "Don't Know", "Refused"] | | | |
| 0 | PD RFQ-U | 5 | pdrfuq_concussion | Have you ever had a head injury or concussion? {0:No, 1:Yes, 2:Possibly, -9:Don't Konw, -7:Refused} | string | nullable | ["No", "Yes", "Possibly", "Don't Know", | | | |
| 0 | PD RFQ-U | 6 | pdrfuq_ibuprofen | Have you ever regularly taken ibuprofen-based non-aspirin medications, that is, at least two pills per week for 6 months or longer? {0:No, 1:Yes, 2:Possibly, -9:Don't Konw, -7:Refused} | string | nullable | ["No", "Yes", "Don't Know", "Refused"] | | | |
| 0 | PD RFQ-U | 7 | pdrfuq_aspirin | Have you ever regularly taken aspirin, that is, at least two pills per week for 6 months or longer? {0:No, 1:Yes, 2:Possibly, -9:Don't Konw, -7:Refused} | string | nullable | ["No", "Yes", "Don't Know", "Refused"] | | | |
| 0 | PD RFQ-U | 8 | pdrfuq_other_anti_inflmmatory_med | Have you ever regularly taken other anti-inflammatory medications for pain, inflammation, or swelling, that is, at least two pills per week for 6 months or longer? {0:No, 1:Yes, 2:Possibly, -9:Don't Konw, -7:Refused} | string | nullable | ["No", "Yes", "Don't Know", "Refused"] | | | |
| 0 | PD RFQ-U | 9 | pdrfuq_smoke_100 | In your lifetime, have you smoked 100 or more cigarettes (5 packs)? {0:No, 1:Yes, 2:Possibly, -9:Don't Know, -7:Refused} | string | nullable | ["No", "Yes", "Don't Know", "Refused"] | | | |
| 0 | PD RFQ-U | 10 | pdrfuq_smoke_regularly | In your lifetime, have you ever regularly smoked cigarettes, | string | nullable | ["No", "Yes", "Don't Know", "Refused"] | | | |
| 0 | PD RFQ-U | 11 | pdrfuq_smokeless_tabacco | Have you ever used smokeless tobacco such as chewing tobacco or snuff regularly, that is, at least once per day for 6 months or longer? {0:No, 1:Yes, 2:Possibly, -9:Don't Konw, -7:Refused} | string | nullable | ["No", "Yes", "Don't Know", "Refused"] | | | |
| 0 | PD RFQ-U | 12 | pdrfuq_alcohol_100 | In your lifetime, have you drunk 100 or more alcoholic drinks (beer, wine, liquor, spirits)? {0:No, 1:Yes, 2:Possibly, -9:Don't Konw, -7:Refused} | string | nullable | ["No", "Yes", "Don't Know", "Refused"] | | | |
| 0 | PD RFQ-U | 13 | pdrfuq_alcohol_regularly | In your lifetime, have you ever regularly drunk alcohol, that is, at least one drink per week for 6 months or longer? {0:No, 1:Yes, 2:Possibly, -9:Don't Konw, -7:Refused} | string | nullable | ["No", "Yes", "Don't Know", "Refused"] | | | |
| 0 | DAT_imaging | 0 | dat_low_dopmaine_reuptake | DAT dopamine re-uptake negative. DAT dopamine transporter binding normal = SWEDD = 1. DAT binding abnormal = PD = 0 | string | nullable | y.isin([0,1]) | | | |
| 0 | DAT_imaging | 1 | dat_sbr_caudate_right | SBR for right caudate | numeric | nullable | (y>0)&(y<10) | | | |
| 0 | DAT_imaging | 2 | dat_sbr_caudate_left | SBR for left caudate | numeric | nullable | (y>0)&(y<10) | | | |
| 0 | DAT_imaging | 3 | dat_sbr_caudate_mean | SBR mean for caudate | numeric | nullable | (y>0)&(y<10) | | | |
| 0 | DAT_imaging | 4 | dat_sbr_putamen_right | SBR for right putamen | numeric | nullable | (y>0)&(y<10) | | | |
| 0 | DAT_imaging | 5 | dat_sbr_putamen_left | SBR for left putamen | numeric | nullable | (y>0)&(y<10) | | | |
| 0 | DAT_imaging | 6 | dat_sbr_putamen_mean | SBR mean for putamine | numeric | nullable | (y>0)&(y<10) | | | |
| 0 | DAT_imaging | 7 | dat_sbr_striatum_mean_right | Mean SBR for right caudate and putamen | numeric | nullable | (y>0)&(y<10) | | | |
| 0 | DAT_imaging | 8 | dat_sbr_striatum_mean_left | Mean SBR for left caudate and putamen | numeric | nullable | (y>0)&(y<10) | | | |
| 0 | DAT_imaging | 9 | dat_sbr_striatum_mean | Mean SBR | numeric | nullable | (y>0)&(y<10) | | | |
| 0 | MIBG_imaging | 1 | HM_ratio_early | | numeric | nullable | (y>0)&(y<10) | | | |
| 0 | MIBG_imaging | 2 | HM_ratio_delay | | numeric | nullable | (y>0)&(y<10) | | | |
