## Supplementary material for "Defining the causes of sporadic Parkinson’s disease in the global Parkinson’s genetics program (GP2)": Global Parkinson's Genetics Program (GP2) - Banner Author List

This project was supported by the Global Parkinson's Genetics Program (GP2). GP2 is funded by the Aligning Science Across Parkinson's (ASAP) initiative and implemented by The Michael J. Fox Foundation for Parkinson's Research. For a complete list of GP2 members see https://gp2.org.

Members: 211
Version date: 22 March 2023

| Country | Name | Institution | Funders and Disclosures |
|---|---|---|---|
| Argentina | Emilia M Gatto | Sanatorio de la Trinidad Mitre - INEBA | Nothing to declare |
| | Marcelo Kauffman | Hospital JM Ramos Mejia | Nothing to declare |
| Armenia | Samson Khachatryan | Somnus Neurology Clinic | Nothing to declare |
| | Zaruhi Tavadyan | Somnus Neurology Clinic | Nothing to declare |
| Australia | Claire E Shepherd | Neuroscience Research Australia | The Sydney Brain Bank is located at and supported by Neuroscience Research Australia |
| | Julie Hunter | ANZAC Research Institute | Nothing to declare |
| | Kishore Kumar | Garvan Institute of Medical Research and Concord Repatriation General Hospital | Paul Ainsworth Family Foundation |
| | Melina Ellis | Concord Hospital | Nothing to declare |
| | Miguel E. Rentería | QIMR Berghofer Medical Research Institute | The Australian Parkinson's Genetics Study is supported by the Shake It Up Australia Foundation and The Michael J. Fox Foundation for Parkinson's Research |
| | Sulev Koks | Murdoch University | Nothing to declare |
| Austria | Alexander Zimprich | Medical University Vienna Austria | Nothing to declare |
| Brazil | Artur F. Schumacher-Schuh | Universidade Federal do Rio Grande do Sul | Nothing to declare |
| | Carlos Rieder | Federal University of Health Sciences of Porto Alegre | Nothing to declare |

| | | | |
|---|---|---|---|
| | Vitor Tumas | University of São Paulo | Nothing to declare |
| Canada | Edward A. Fon | Montreal Neurological Institute | Nothing to declare |
| | Oury Monchi | Institut universitaire de gériatrie de Montréal | CIHR, Brain Canada, Parkinson Canada |
| | Ted Fon | McGill University | Nothing to declare |
| Chile | Benjamin Pizarro Galleguillos | Universidad de Chile | Nothing to declare |
| | Marcelo Miranda | Fundación Diagnosis | Nothing to declare |
| | Maria Leonor Bustamante | Universidad de Chile | Nothing to declare |
| | Patricio Olguin | Universidad de Chile | Nothing to declare |
| | Paula Saffie Awad | Universidade Federal do Rio Grande do Sul | Nothing to declare |
| | Pedro Chana | CETRAM | Nothing to declare |
| China | Beisha Tang | Central South University | Nothing to declare |
| | Huifang Shang | West China Hospital Sichuan University | Nothing to declare |
| | Jifeng Guo | Xiangya Hospital | Nothing to declare |
| | Piu Chan | Capital Medical University | Nothing to declare |
| | Wei Luo | Zhejiang University | Nothing to declare |
| | Xiaopu Zhou | The Hong Kong University of Science and Technology | Nothing to declare |
| Colombia | Gonzalo Arboleda | Universidad Nacional de Colombia | Nothing to declare |
| | Jorge Orozco | Fundación Valle del Lili | Nothing to declare |
| | Marlene Jimenez del Rio | University of Antioquia | Nothing to declare |
| Costa Rica | Alvaro Hernandez | University of Costa Rica | Nothing to declare |
| Egypt | Mohamed Salama | The American University in Cairo | The AUC/ ASRT/ DAAD |
| | Walaa A. Kamel | Beni-Suef University | Nothing to declare |
| Ethiopia | Yared Z. Zewde | Addis Ababa University | Nothing to declare |
| France | Alexis Brice | Paris Brain Institute | Nothing to declare |
| | Jean-Christophe Corvol | Sorbonne Université | Nothing to declare |
| Germany | Brit Mollenhauer | University Medical Center Göttingen | Nothing to declare |

| | | | |
|---|---|---|---|
| | Christine Klein | University of Lübeck | CK serves as a medical Advisor to Centogene on genetic testing reports in the field of movement disorders, except Parkinson's disease, and is a member of the Scientific Advisory Board of Retromer Therapeutics |
| | Eva-Juliane Vollstedt | University of Lübeck | Nothing to declare |
| | Franziska Hopfner | University Hospital, LMU Munich | Nothing to declare |
| | Günter Höglinger | University Hospital, LMU Munich | Nothing to declare |
| | Harutyun Madoev | University of Lübeck | Nothing to declare |
| | Johanna Junker | University of Lübeck | Nothing to declare |
| | Katja Lohmann | University of Lübeck | Nothing to declare |
| | Lara M. Lange | University of Lübeck | Nothing to declare |
| | Manu Sharma | University of Tübingen | Dr. Sharma is further funded by the Michael J. Fox Foundation for Parkinson's Research, USA Genetic Diversity in PD Program: GAP-India Grant ID: 009411. |
| | Sergio Groppa | University of Mainz | Nothing to declare |
| | Thomas Gasser | University of Tübingen | Nothing to declare |
| | Zih-Hua Fang | The German Center for Neurodegenerative Diseases | Nothing to declare |
| Ghana | Albert Akpalu | University of Ghana | Nothing to declare |
| | Vida Obese | University College London | Nothing to declare |
| Greece | Georgia Xiromerisiou | University of Thessaly | Nothing to declare |
| | Georgios Hadjigorgiou | University of Thessaly | Nothing to declare |
| | Ioannis Dagklis | Aristotle University of Thessaloniki | Nothing to declare |
| | Ioannis Tarnanas | Ionian University | Nothing to declare |
| | Leonidas Stefanis | Biomedical Research Foundation of the Academy of Athens | PPMI2 (funded by MJFF), ALAMEDA (H2020 grant), funding by HFRI |

| | | | |
|---|---|---|---|
| | Maria Stamelou | Diagnostic and Therapeutic Centre HYGEIA Hospital | Nothing to declare |
| | Efthymios Dadiotis | University of Thessaly | Nothing to declare |
| Honduras | Alex Medina | Hospital San Felipe | Nothing to declare |
| Hong Kong | Germaine Hiu-Fai Chan | Queen Elizabeth Hospital | Nothing to declare |
| | Nancy Ip | Hong Kong University of Science and Technology | Nothing to declare |
| | Nelson Yuk-Fai Cheung | Queen Elizabeth Hospital | Nothing to declare |
| | Phillip Chan | Hong Kong University of Science and Technology | Nothing to declare |
| India | Asha Kishore | Aster Medcity | Michael J Fox Foundation |
| | Divya KP | Sree Chitra Tirunal Institute for Medical Sciences and Technology | Nothing to declare |
| | Pramod Pal | National Institute of Mental Health & Neurosciences | Nothing to declare |
| | Prashanth Lingappa Kukkle | Manipal Hospital | Nothing to declare |
| | Roopa Rajan | All India Institute of Medical Sciences | Nothing to declare |
| | Rupam Borgohain | Nizam's Institute Of Medical Sciences | Nothing to declare |
| Italy | Andrea Quattrone | Magna Græcia University of Catanzaro | Nothing to declare |
| | Enza Maria Valente | University of Pavia | Nothing to declare |
| | Lucilla Parnetti | University of Perugia | Nothing to declare |
| | Micol Avenali | University of Pavia | Nothing to declare |
| | Tommaso Schirinzi | University of Rome Tor Vergata | Nothing to declare |
| Japan | Manabu Funayama | Juntendo University | Nothing to declare |
| | Nobutaka Hattori | Juntendo University | Nothing to declare |
| | Tomotaka Shiraishi | Jikei University | Nothing to declare |
| Kazakhstan | Altynay Karimova | Institute of Neurology and Neurorehabilitation | Nothing to declare |

| | | | |
|---|---|---|---|
| | Rauan Kaiyrzhanov | University College London | Nothing to declare |
| | Gulnaz Kaishibayeva | Institute of Neurology and Neurorehabilitation | Nothing to declare |
| Luxembourg | Rejko Krüger | University of Luxembourg | Nothing to declare |
| Malaysia | Ai Huey Tan | University of Malaya | Nothing to declare |
| | Azlina Ahmad-Annuar | University of Malaya | Nothing to declare |
| | Mohamed Ibrahim Norlinah | Universiti Kebangsaan Malaysia | Nothing to declare |
| | Nor Azian Abdul Murad | UKM Medical Molecular Biology Institute | Nothing to declare |
| | Norlinah Mohamed Ibrahim | Universiti Kebangsaan Malaysia Medical Centre | Nothing to declare |
| | Shahrul Azmin | Universiti Kebangsaan Malaysia Medical Centre | Nothing to declare |
| | Shen-Yang Lim | University of Malaya | Nothing to declare |
| | Wael Mohamed | International Islamic University Malaysia | Nothing to declare |
| | Yi Wen | University of Malaya | Nothing to declare |
| Mexico | Daniel Martinez-Ramirez | Tecnologico de Monterrey | Nothing to declare |
| | Mayela Rodriguez-Violante | Instituto Nacional de Neurologia y Neurocirugia | Nothing to declare |
| | Paula Reyes-Pérez | Universidad Nacional Autónoma de México | Nothing to declare |
| Nepal | Rajeev Ojha | Tribhuvan University | Nothing to declare |
| New Zealand | Tim J. Anderson | University of Otago | Health Research Council of New Zealand; Ministry of Business Innovation and Employment, New Zealand, Neurological Foundation of New Zealand |
| | Toni L. Pitcher | University of Otago | Health Research Council of New Zealand |
| Nigeria | Arinola Sanyaolu | University of Lagos | Nothing to declare |

| | | | |
|---|---|---|---|
| | Njideka Okubadejo | University of Lagos | Michael J Fox Foundation; Tertiary Education Trust Fund (TETFUND) National Research Fund |
| | Olaitan Okunoye | University College London | Nothing to declare |
| | Oluwadamilola Ojo | University of Lagos | Nothing to declare |
| Norway | Jan O. Aasly | Norwegian University of Science and Technology | Nothing to declare |
| | Lasse Pihlstrøm | Oslo University Hospital | Southeastern Regional Health Authority, Norway |
| | Manuela Tan | Oslo University Hospital | Southeastern Regional Health Authority, Norway and Michael J. Fox Foundation |
| Pakistan | Shoaib Ur-Rehman | University of Science and Technology Bannu | Nothing to declare |
| Peru | Miguel Inca-Martinez | Cleveland Clinic | Nothing to declare |
| | Mario Cornejo-Olivas | Instituto Nacional de Ciencias Neurologicas | Michael J. Fox Foundation for Parkinson's Research and Aligning Science Across Parkinson's Initiative |
| Puerto Rico | Angel Vinuela | University of Puerto Rico | Nothing to declare |
| Russia | Elena Iakovenko | Research Center of Neurology | Nothing to declare |
| Saudi Arabia | Bashayer Al Mubarak | King Faisal Specialist Hospital and Research Center | Nothing to declare |
| | Muhammad Umair | King Abdullah International Medical Research Center | Nothing to declare |
| Singapore | Jia Nee Foo | Nanyang Technological University | Singapore National Medical Research Council (MOH-000559) |
| | Eng-King Tan | National Neuroscience Institute | Singapore National Medical Research Council (MOH-OFLCG-000207) |
| South Africa | Jonathan Carr | University of Stellenbosch | Nothing to declare |
| | Soraya Bardien | Stellenbosch University | Funded by the National Research Foundation of South Africa [Grant Number 129249] |

| | | | |
|---|---|---|---|
| South Korea | Beomseok Jeon | Seoul National University Hospital | Nothing to declare |
| | Yun Joong Kim | Yongin Severance Hospital | Nothing to declare |
| Spain | Janet Hoenicka | Institut de Recerca Sant Joan de Deu | Fondo de Investigación Sanitaria, Instituto Salud Carlos III, Grant PI019/00126 |
| | Maria Teresa Periñan Tocino | Instituto de Biomedicina de Sevilla | Nothing to declare |
| | Pau Pastor | University Hospital Germans Trias i Pujol | Nothing to declare |
| | Katrin Beyer | Germans Trias i Pujol Research Institute | Nothing to declare |
| | Ignacio Alvarez | Hospital Universitari Mutua de Terrassa | Nothing to declare |
| Sudan | Sarah El-Sadig | University of Khartoum | Nothing to declare |
| Taiwan | Chin-Hsien Lin | National Taiwan University Hospital | Nothing to declare |
| | Hsiu-Chuan Wu | Chang Gung Memorial Hospital | Nothing to declare |
| | Pin-Jui Kung | National Taiwan University | Nothing to declare |
| | Ruey-Meei Wu | National Taiwan University Hospital | I have funding from 1. Minister of Science and Technology, Taiwan Government; 2. National Taiwan University, 3 Parkinson foundation, USA, 4. Michael J Fox Foundation. |
| | Serena Wu | Chang Gung University | Nothing to declare |
| | Yihru Wu | Chang Gung Memorial Hospital | Nothing to declare |
| Tunisia | Rim Amouri | National Institute Mongi Ben Hamida of Neurology | Nothing to declare |
| | Samia Ben Sassi | Mongi Ben Hmida National Institute of Neurology | Nothing to declare |
| Turkey | A. Nazlı Başak | Koç University | Kirac Foundation and Koc Univ. |
| | Gencer Genc | Şişli Etfal Training and Research Hospital | Nothing to declare |
| | Özgür Öztop Çakmak | Koç University | Nothing to declare |

| | | | |
|---|---|---|---|
| | Sibel Ertan | Koç University | Nothing to declare |
| United Kingdom | Alastair Noyce | Queen Mary University of London | Prof. Noyce reports grants from Parkinson's UK, Barts Charity, Cure Parkinson's, NIHR, Innovate UK, Virginia Keiley benefaction, Alchemab, Aligning Science Across Parkinson's and Michael J Fox Foundation. Consultancy and personal fees from Astra Zeneca, AbbVie, Profile, Roche, Biogen, UCB, Bial, Charco Neurotech, uMedeor and Britannia. |
| | Alejandro Martínez-Carrasco | University College London | Global Parkinson's Genetics Program |
| | Camille Carroll | University of Plymouth | C Carroll receives salary from University of Plymouth, University Hospitals Plymouth NHS Trust and National Institute of Health Research; she has received advisory, consulting, and/or lecture fees from AbbVie, Bial, Lundbeck, Global Kinetics, Britannia and Medscape, and research funding from Parkinson's UK, Edmond J Safra Foundation, National Institute of Health Research and Cure Parkinson's |
| | Claire Bale | Parkinson's UK | Nothing to declare |
| | Eleanor J. Stafford | University College London | Nothing to declare |
| | Henry Houlden | University College London | Nothing to declare |
| | Huw R Morris | University College London | Dr Morris is employed by UCL. In the last 12 months he reports paid consultancy from Roche and Amylyx ; lecture fees/honoraria - BMJ, Kyowa Kirin, Movement Disorders Society. Research Grants from Parkinson's UK, Cure Parkinson's Trust, PSP Association, CBD Solutions, Drake Foundation, Medical Research Council, Michael J Fox Foundation. Dr Morris is a co-applicant on a patent application related to C9ORF72 - |

| | | | |
|---|---|---|---|
| | | | Method for diagnosing a neurodegenerative disease (PCT/GB2012/052140) |
| | John Hardy | University College London | Nothing to declare |
| | Kin Ying Mok | University College London | Nothing to declare |
| | Mie Rizig | University College London | Nothing to declare |
| | Nicholas Wood | University College London | ASAP-CRN |
| | Nigel Williams | Cardiff University | Parkinson's UK |
| | Patrick Alfryn Lewis | Royal Veterinary College University of London | MJFF, UKRI (BBSRC, EPSRC), Parkinson’s UK, ASAP research network network |
| | Rimona Weil | University College London | Nothing to declare |
| | Simona Jasaitye | University College London | Nothing to declare |
| | Simon Stott | Cure Parkinson's | Employee of Cure Parkinson's |
| | Sumit Dey | Queen Mary University of London | Nothing to declare |
| USA | Alberto Espay | University of Cincinnati | Nothing to declare |
| | Alyssa O'Grady | The Michael J. Fox Foundation for Parkinson's Research | Nothing to declare |
| | Andrew B Singleton | National Institute on Aging | Michael J. Fox Foundation for Parkinson's Disease Research and Aligning Science Across Parkinson's Initiative |
| | Andrew K. Sobering | Augusta University/ University of Georgia Medical Partnership | Nothing to declare |
| | Bernadette Siddiqi | The Michael J. Fox Foundation for Parkinson's Research | Nothing to declare |
| | Bradford Casey | The Michael J. Fox Foundation for Parkinson's Research | Nothing to declare |
| | Brian Fiske | The Michael J. Fox Foundation for Parkinson's Research | Nothing to declare |
| | Cabell Jonas | Mid-Atlantic Permanente Medical Group | Nothing to declare |

| | | | |
|---|---|---|---|
| | Caroline B. Pantazis | National Institutes of Health | Nothing to declare |
| | Claire Wegel | Indiana University | Nothing to declare |
| | Cornelis Blauwendraat | National Institutes of Health | Nothing to declare |
| | Dan Vitale | National Institutes of Health | Nothing to declare |
| | Deborah Hall | Rush University | Nothing to declare |
| | Dena Hernandez | National Institutes of Health | Nothing to declare |
| | Ejaz Shiamim | Kaiser Permanente | Nothing to declare |
| | Ekemini Riley | Coalition for Aligning Science | Nothing to declare |
| | Faraz Faghri | National Institutes of Health | F.F.'s participation in this research was supported in part by the Intramural Research Program of the NIH, National Institute on Aging (NIA), National Institutes of Health, Department of Health and Human Services; project number ZO1 AG000535, as well as the National Institute of Neurological Disorders and Stroke. F.F.'s participation in this project was part of a competitive contract awarded to Data Tecnica International LLC by the National Institutes of Health to support open science research. |
| | Geidy E. Serrano | Banner Sun Health Research Institute | Banner Sun Health Research Institute Brain and Body Donation Program of Sun City, Arizona for the provision of human biological materials. The Brain and Body Donation Program has been supported by the National Institute of Neurological Disorders and Stroke (U24 NS072026 National Brain and Tissue Resource for Parkinson’s Disease and Related Disorders), the National Institute on Aging (P30 AG19610 and P30AG072980, Arizona Alzheimer’s Disease Center), the Arizona Department of Health Services (contract 211002, Arizona Alzheimer’s Research Center), the Arizona Biomedical |

| | | | |
|---|---|---|---|
| | | | Research Commission (contracts 4001, 0011, 05-901 and 1001 to the Arizona Parkinson's Disease Consortium) and the Michael J. Fox Foundation for Parkinson's Research. |
| | Hampton Leonard | National Institutes of Health | H.L.L is supported by a competitive contract awarded to Data Tecnica International LLC by the National Institutes of Health to support open science research |
| | Hirotaka Iwaki | Data Tecnica International | Nothing to declare |
| | Honglei Chen | Michigan State University | NIH/DoD/Parkinson Foundation/MSU Foundation/Gibby vs. Parky Foundation - No COI to disclose |
| | Ignacio F. Mata | Cleveland Clinic | Funding from MJFF and NIH |
| | Ignacio Juan Keller Sarmiento | Northwestern University | Nothing to declare |
| | Jared Williamson | Kaiser Permanente | Nothing to declare |
| | Jonggeol Jeff Kim | National Institutes of Health | Nothing to declare |
| | Joseph Jankovic | Baylor College of Medicine | Nothing to declare |
| | Joshua Shulman | Baylor College of Medicine | Collection of samples and data at Baylor College of Medicine was supported by the Huffington Foundation. |
| | Justin C. Solle | The Michael J. Fox Foundation for Parkinson's Research | Nothing to declare |
| | Kaileigh Murphy | The Michael J. Fox Foundation for Parkinson's Research | Nothing to declare |
| | Karen Nuytemans | University of Miami | This work has been supported by the American Parkinson Disease Association and the Margaret Q. Landenberger Research Foundation. |
| | Karl Kieburtz | Beth Israel Deaconess Medical Center | Nothing to declare |

| | | | |
|---|---|---|---|
| | Kenneth Marek | Institute for Neurodegenerative Disorders | Consultant for Michael J Fox Foundation, GE Healthcare, Roche, UCB, BIAL, Denali, Takeda, , Cerapsir, UCB, Biohaven, Neuron23, Aprinoia, Astellas, Calico, Genentech, Invicro |
| | Kristin S. Levine | Data Tecnica International | K.S.L. is supported by a competitive contract awarded to Data Tecnica International LLC by the National Institutes of Health to support open science research |
| | Lana M. Chahine | University of Pittsburgh | Nothing to declare |
| | Laurel Screven | National Institute on Aging | Nothing to declare |
| | Lisa Shulman | University of Maryland | Nothing to declare |
| | Luca Marsili | University of Cincinnati | Nothing to declare |
| | Maggie Kuhl | The Michael J. Fox Foundation for Parkinson's Research | Nothing to declare |
| | Marissa Dean | University of Alabama at Birmingham | Dr. Dean is an investigator in studies funded by Abbvie, Inc., Hoffmann-La Roche, CHDI Foundation, Inc., Annexon, Inc., Retrophin, Inc, Neurocrine Biosciences, UniQure Biopharma B.V., Praxis Precision Medicines, Neuraly, Inc., Michael J. Fox Foundation for Parkinson's Research, and US Army Medical Research and Materiel Command (grant#W81XWH-18-1-0508). In addition, Dr. Dean receives support through the Huntington's Disease Society of American Centers of Excellence program. |
| | Mary B Makarious | National Institutes of Health | Nothing to declare |
| | Mathew Koretsky | National Institutes of Health | Nothing to declare |

| | | | |
|---|---|---|---|
| | Mike A. Nalls | National Institutes of Health | M.A.N.'s participation in this project was part of a competitive contract awarded to Data Tecnica International LLC by the National Institutes of Health to support open science research. M.A.N. also currently serves as an advisor for Clover Therapeutics and Neuron23 Inc. |
| | Naomi Louie | The Michael J. Fox Foundation for Parkinson's Research | Nothing to declare |
| | Niccolò Emanuele Mencacci | Northwestern University | Nothing to declare |
| | Roy Alcalay | Columbia University | Dr. Alcalay is funded by the Michael J. Fox Foundation for Parkinson's Research and the Parkinson's Foundation. He received consultation fees from Avrobio, Caraway, GSK, Merck, Sanofi, Ono Therapeutics and Takeda |
| | Sara Bandres-Ciga | National Institutes of Health | Nothing to declare |
| | Sohini Chowdhury | The Michael J. Fox Foundation for Parkinson's Research | Nothing to declare |
| | Sonya Dumanis | Aligning Science Across Parkinson's | Nothing to declare |
| | Steven Lubbe | Northwestern University | Nothing to declare |
| | Tao Xie | University of Chicago | Nothing to declare |
| | Tatiana Foroud | Indiana University | The Michael J. Fox Foundation for Parkinson's Research |
| | Thomas Beach | Sun Health Research Institution | Banner Sun Health Research Institute Brain and Body Donation Program of Sun City, Arizona for the provision of human biological materials. The Brain and Body Donation Program has been supported by the National Institute of Neurological Disorders and Stroke (U24 |

| | | | |
|---|---|---|---|
| | | | NS072026 National Brain and Tissue Resource for Parkinson's Disease and Related Disorders), the National Institute on Aging (P30 AG19610 and P30AG072980, Arizona Alzheimer's Disease Center), the Arizona Department of Health Services (contract 211002, Arizona Alzheimer's Research Center), the Arizona Biomedical Research Commission (contracts 4001, 0011, 05-901 and 1001 to the Arizona Parkinson's Disease Consortium) and the Michael J. Fox Foundation for Parkinson's Research |
| | Todd Sherer | The Michael J Fox Foundation for Parkinson's Research | Nothing to declare |
| | Yeajin Song | National Institutes of Health | Nothing to declare |
| Vietnam | Duan Nguyen | Hue University | Nothing to declare |
| | Toan Nguyen | Hue University | Nothing to declare |
| Zambia | Masharip Atadzhanov | University of Zambia | Nothing to declare |
